# Increasing SARS-CoV2 cases, hospitalizations, and deaths among the vaccinated populations during the Omicron (B.1.1.529) variant surge in UK

**DOI:** 10.1101/2022.06.28.22276926

**Authors:** Venkata R. Emani, Vivek K. Pallipuram, Kartik K. Goswami, Kailash R. Maddula, Raghunath Reddy, Abirath S. Nakka, Sravya Panga, Nikhila K. Reddy, Nidhi K. Reddy, Dheeraj Nandanoor, Sanjeev Goswami

## Abstract

**BACKGROUND:** There were increased SARS-CoV2 hospitalizations and deaths noted during Omicron (B.1.1.529) variant surge in the UK despite decreased cases, and the reasons are unclear.

**METHODS:** In this retrospective observational study, we analyzed reported SARS-CoV2 cases, hospitalizations, and deaths during the COVID-19 pandemic in the UK. We also analyzed variables that affect the outcomes (including ethnicity, deprivation score, vaccination disparities, and pre-existing conditions). We also analyzed the vaccine effectiveness among those ≥18 years of age (from August 16, 2021 to March 27, 2022).

**RESULTS:** Of the total cases (n= 22,072,550), hospitalizations (n=848,911), and deaths (n=175,070) due to COVID-19 in the UK; 51.3% of cases (n=11,315,793), 28.8% of hospitalizations (n=244,708), and 16.4% of deaths (n=28,659) occurred during the Omicron variant surge as of May 1, 2022. During the latter part of the Omicron variant surge (February 28 - May 1, 2022 period), we observed a significant increase in the proportion of cases (23.7% vs 40.3%; RR1.70 [1.70-1.71]; p<0.001) and hospitalizations (39.3% vs 50.3%; RR1.28 [1.27-1.30]; p<0.001) among ≥50 years of age, and deaths (67.89% vs 80.07%; RR1.18 [1.16-1.20]; p<0.001) among ≥75 years of age compared to the earlier period (December 6, 2021-February 27, 2022) during the Omicron variant surge. Using the available data from vaccine surveillance reports, we compared the Omicron variant surge (December 27, 2021-March 20, 2022) with the Delta variant surge (August 16-December 5, 2021). Our comparative analysis shows a significant decline in case fatality rate (all ages [0.21% vs 0.39%; RR 0.54 (0.52-0.55); p<0.001], over 18 years of age [0.25% vs 0.58%; RR 0.44 (0.43-0.45); p<0.001], and over 50 years of age [0.72% vs 1.57%; RR 0.46 (0.45-0.47); P<0.001]) and the risk of hospitalizations (all ages [0.62% vs 0.99%; RR 0.63 (0.62-0.64); p<0.001], over 18 years of age [0.67% vs 1.38%; RR 0.484 (0.476-0.492); p<0.001], and over 50 years of age [1.45% vs 2.81%; RR 0.52 (0.51-0.53); p<0.001]). Both the unvaccinated (0.41% vs 0.77%; RR 0.54 (0.51-0.57); p<0.001) and vaccinated (0.25% vs 0.59%; RR 0.43 (0.42-0.44); p<0.001) populations of over 18 years of age showed a significant decline in the case fatality rate during the Omicron variant surge when compared to the Delta variant surge. In summary, a significant decline in the risk of hospitalizations was observed both among the unvaccinated (1.27% vs 2.92%; RR 0.44 (0.42-0.45); p<0.001) and vaccinated (0.65% vs 1.19%; RR 0.54 (0.53-0.55); p<0.001) populations of over 18 years of age during the same period. We observed negative vaccine effectiveness (VE) for the third dose since December 20, 2021, with a significantly increased proportion of SARS-CoV2 cases hospitalizations, and deaths among the vaccinated; and a decreased proportion of cases, hospitalizations, and deaths among the unvaccinated. The pre-existing conditions were present in 95.6% of all COVID-19 deaths. We also observed various ethnicity, deprivation score, and vaccination rate disparities that can adversely affect hospitalizations and deaths among the compared groups based on vaccination status.

**CONCLUSIONS:** There is no discernable optimal vaccine effectiveness among ≥18 years of age and vaccinated third dose population since the beginning (December 20, 2021) of the Omicron variant surge. Other data including pre-existing conditions, ethnicity, deprivation score, and vaccination rate disparities need to be adjusted by developing validated models for evaluating VE against hospitalizations and deaths. Both the vaccinated and unvaccinated populations showed favorable outcomes with a significant decline in case fatality rate and risk of hospitalizations during the Omicron variant surge. The suboptimal vaccine effectiveness with an increased proportion of cases among the vaccinated population was associated with a significantly increased proportion of hospitalizations and deaths during the Omicron variant surge. This underscores the need to prevent infections, especially in the elderly vaccinated population irrespective of vaccination status by employing uniform screening protocols and protective measures.

## INTRODUCTION

Following the World Health Organization (WHO), the UK Health Security Agency (UKHSA) designated the Omicron variant (B.1.1.529) of the SARS-CoV2, the virus that causes COVID-19, as a variant of concern on November 27, 2021^1–3^. The Omicron variant spread rapidly across the United Kingdom (UK) and became a predominant strain with 365,375 confirmed cases as of December 23, 2021^4, 5^. UK reported a total of 11.3 million SARS CoV2 cases during the Omicron variant surge in twenty-one weeks as of May 1, 2022 with a majority of the newly confirmed infections of Omicron variant lineages (B.1.1.529 and B.1.1.529.BA.2)^6, 7^.

The UK COVID-19 vaccination program started in December 2020, with two doses of either mRNA-based vaccines (BNT162b2 and mRNA-1273) or adenoviral vector-based vaccine (ChAdOx1 nCoV-19). Approximately 73.9% of the ≥50 years old population were vaccinated by May 2, 2021 during the beginning of the Delta variant (B.1.617.2) surge^8^.

After the initial approval of the third dose (booster) to only the immunocompromised population, the UK authorities approved the third dose to ≥50 years old general population on September 14, 2021 to be administered no sooner than within six months after the primary course^9, 10^. The third dose vaccination was subsequently expanded in the UK to ≥18 years old population on November 29, 2021 and changed the booster timing to no sooner than three months after the primary course due to changing risk posed by the Omicron variant^11^. The UK government later announced the fourth dose (booster) in March 2022 ^12, 13^.

While the vaccine immunity was reported waning during the Delta variant surge elsewhere, the initial Public Health England reported good vaccine effectiveness with two doses of BNT162b2 and ChAdOx1 nCoV-19 vaccines among those with the delta variant (B.1.617.2)^14–17^. A later study reported a waning of effectiveness for symptomatic infections with Delta variant, but limited waning in effectiveness against SARS-CoV2 related hospitalizations and deaths^18^. During this time, the UK technical briefings also showed a growing problem of breakthrough infections in confirmed Delta variant cases among the ≥50 years age group with 75.3% of breakthrough cases (63.5% of hospitalizations and 67.0% of deaths) as of September 12, 2021^19–21^.

Similarly, the weekly vaccine surveillance reports also show a trend of increased cases in the vaccinated third booster population than the unvaccinated population during the Omicron variant surge^22^. These reports highlight an alarming trend of increased hospitalizations and deaths among the vaccinated third dose population until they stopped reporting the underlying data after March 27, 2022^22^. The UK COVID-19 dashboard is also reporting a relatively higher number of daily hospitalizations and deaths during the later period of the Omicron variant surge in March-April 2022, despite substantially lower cases reported^6^.

In this retrospective observational study, the changing pattern of SARS-CoV2 cases, hospitalizations, and deaths among various age groups during the Omicron variant surge relative to earlier surges was analyzed. We also analyzed SARS-CoV2 cases, hospitalizations, and deaths among ≥18 years old population based on the vaccination status during the Omicron variant surge and compared it with the Delta variant surge.

## METHODS

### STUDY DESIGN

In this retrospective observational study, we analyzed the nationwide available data of the confirmed SARS-CoV2 cases, hospitalizations, and deaths in the UK from the beginning of the COVID-19 pandemic until May 1, 2022. The vaccine effectiveness (VE) was calculated based on the data from the UK vaccine surveillance reports and the UK technical briefings until March 27, 2022. Specifically, we use Equation 1 to calculate the vaccine effectiveness where *IRR* is the incidence rate ratio, *Fraction_unvaccinated_*denotes the fraction of cases among the unvaccinated population, and *Fraction_vaccinated_*represents the fraction of cases among the vaccinated population.

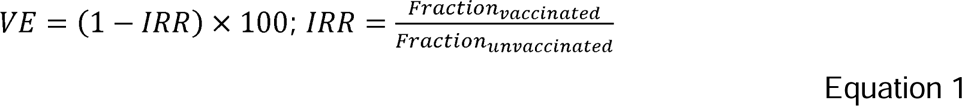

It is worth noting that VE (as per Equation 1) may achieve a maximum value of 100%, implying that the vaccinated population observes a significantly *lower* proportion of cases versus the unvaccinated. Conversely, VE can also achieve an extremely low negative value, implying that the vaccinated population observes a significantly *higher* proportion of cases versus the vaccinated population. We also analyzed the data on racial ethnicity, pre-existing conditions, Indices of Multiple Deprivation (IMD) score, and adaptation of SARS-CoV2 vaccination among racially ethnic groups.

### DATA SOURCES AND ANALYSIS

#### Total UK SARS-COV2 cases, hospitalizations and deaths

Analysis of the SARS-CoV2 cases (specimen date), SARS-CoV2 hospitalizations, and SARS-CoV2 deaths (deaths within 28 days of the positive test by the data of deaths) in the UK was performed from the beginning of the pandemic until May 1, 2022 using the data from the UK coronavirus dashboard^6^. The case fatality rate (CFR) was calculated as the percentage of SARS-CoV2 deaths of the reported cases, and the risk of hospitalizations (RH) was calculated as the percentage of SARS-CoV2 hospitalizations of the reported cases (Equation 2). The hospitalization death rate (HDR) was calculated as the percentage of SARS-CoV2 deaths of the reported hospitalizations (Equation 3). The various periods of the pandemic chosen for the analysis was based on the starting period of the surges and changing pattern of outcomes from February 28, 2022 during the latter part of the Omicron variant surge until May 1, 2022 study period.

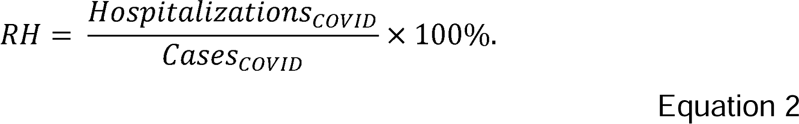

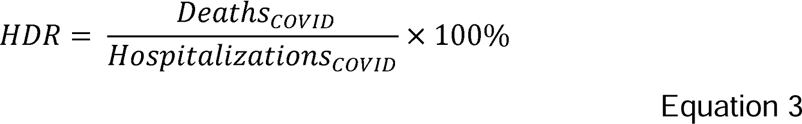

#### SARS-COV2 cases, hospitalizations among the age groups in England and SARS- CoV2 deaths in England and Wales

##### Cases

We analyzed SARS-CoV2 cases reported under pillar 1 (public health laboratories and hospitals) and pillar 2 (community testing) in England among various age groups (0-19, 20-29, 30-49, 50-69, ≥50, ≥70 and ≥80 years of age) from September 28, 2020 to May 1, 2022 using the data from National Flue and COVID-19 surveillance reports (2020-2021 season and 2021-2022 season)^8, 23^.

##### Hospitalizations

We examined the daily hospitalizations in England among various age groups (0-17, 18-34, 35-54, 55-64, ≥65-74 and ≥75 years of age) from October 12, 2020 to May 1, 2022 using the National Health Services (NHS) database of COVID-19 hospital activity^24^.

##### Deaths

We analyzed the weekly SAVRS-CoV2 deaths among various age groups (0- 19, 20-29, 30-49, 50-69, ≥70-74, ≥50 and ≥75 years of age) in England and Wales from the beginning of the pandemic in March 2020 to May 1, 2022 using the UK Office for National Statistics dataset^25^. We compared the weekly SARS-CoV2 deaths among various age groups during each surge with prior surges.

We tabulated and analyzed the changing pattern of cases, hospitalizations, and deaths during various surges.

#### Analysis of the factors known to impact the SARS-CoV2 outcomes

##### Pre-existing conditions among COVID-19 deaths and COVID-19 deaths among ethnic groups in England

The National Health Services weekly data of COVID-19 deaths archives was used to analyze the pre-existing conditions among COVID-19 deaths in various age groups^26^.

##### SARS-CoV2 cases, hospitalizations, and deaths among ethnic groups and deprived populations based on IMD score

SARS-CoV2 cases (pillar 1 and pillar 2) per 100, 000 population among various ethnic groups (from June 29, 2020 to May 1, 2022), SARS- COV-2 hospitalizations (excluding ICU/HDU), SARS-CoV2 admission to ICU/HDU units (March 16, 2020 to May 1, 2022), SARS-CoV2 pillar 1 and 2 cases and SARS-CoV2 deaths per 100, 000 population among population-based on Indices of Multiple Deprivation (from June 27, 2020 to April May 1, 2022) were tabulated with weekly data from the National Flu and COVID-19 surveillance reports^8^. The IMD quintile 1 and decile 1 are considered the most deprived and quintile 5 and decile 10 are the least deprived^8^.

##### Vaccine uptake among racial minorities

The weekly vaccine uptake data among various ethnic groups was analyzed for the period ending May 1, 2022^8^.

#### Vaccination data

The National Immunization Management System (NIMS) vaccination data since the beginning of the vaccination program in December 2020, which was published weekly in the National Flu and COVID-19 surveillance reports was used to calculate the vaccinated and unvaccinated population among age groups for a specified period of vaccine effectiveness^8^. The SARS-CoV2 cases, hospitalizations (presented to emergency care within 28 days of a positive test resulting in overnight inpatient admission), and deaths (within 28 days of a positive COVID-19 test) among various age groups (published weekly) with an aggregate of 4 weeks data from August 16, 2021 to March 27, 2022 (for the third dose since December 20, 2021) in the COVID-19 vaccine surveillance reports was used in our data analysis ^22^. The Public Health England technical briefings data of SARS-CoV2 cases, hospitalizations, and deaths among the confirmed cases of Delta variant was used for analysis during the period of June 21, 2021 to September 12, 2021 (included in supplemental appendix)^27^. The UK Health Security Agencies COVID-19 vaccine surveillance reports did not publish the third dose cases, hospitalizations, and deaths until December 19, 2021. We used ≥18 years, pillar 2 cases published by the UKHSA between November 27, 2021 to January 12, 2022 periods to calculate the vaccine effectiveness of the third dose^28^. We also analyzed the vaccine effectiveness among over 50 years population for infections. The SARS-CoV2 events (cases, hospitalizations, and deaths) tested positive with the specimen date ≥14 days post the second dose are considered vaccinated with two doses; the events ≥14 days after receiving the third dose are considered vaccinated with the third dose, and the events post one dose without additional doses of vaccination are considered one dose^22^.

### STATISTICAL ANALYSIS

We conducted the statistical analysis using the R software, version 4.1.3. We compared the cases, hospitalizations, and deaths among the various age groups between the surges to determine the changing pattern of cases, infections, and deaths during the pandemic. Among those experiencing the event (cases, hospitalizations, or deaths), the proportion of change (Δ) compared to the immediate prior period in a particular age group (increased or decreased) was calculated as the relative risk (RR) along with the 95% confidence interval and the p-value.

We used the weekly published National Immunization Management System (NIMS) vaccination data to calculate the population for each age group for vaccine effectiveness^8^. We obtained the number of cases, hospitalizations, and deaths from the vaccine surveillance reports, and the data of these outcomes linked to the NIMS database^22^. The incidence rate was calculated as the number of SARS-CoV2 infections, hospitalizations, or deaths per 100,000 population for each studied group.

The incidence rate ratio (IRR) was calculated among the compared groups using Equation 1. The partially vaccinated (received 1 dose), vaccinated with two doses, and the third dose populations were compared with the unvaccinated for the VE calculations. Vaccine effectiveness was calculated as (1-IRR) x 100 (Equation 1). We performed the two-proportions test with continuity correlation (henceforth two-proportions test) to determine whether the proportions of cases, hospitalizations, and deaths among the vaccinated population are significantly higher than the unvaccinated population. The case fatality rate (CFR), risk of hospitalizations (RH), and the hospitalization death rate (HDR) was calculated for the entire population (all ages), over 18 years and over 50 years of age vaccinated and unvaccinated populations during the Delta variant surge (August 16-December 5, 2021) and Omicron variant surge (December 27, 2021 to March 20, 2022). The proportion of change (Δ) in CFR, RH, and HDR among the age groups (all ages, over 18 years of age and over 50 years of age) during the Omicron variant surge versus the Delta variant surge was calculated as the relative risk (RR) along with the 95% confidence interval and the p-value. Similarly, of the total events (SARS-CoV2 cases, hospitalizations and deaths) among all ages, over 18 years and over 50 years of age, the percentage of the SARS-CoV2 cases, hospitalizations, and deaths among vaccinated and unvaccinated were calculated during the Delta variant surge (August 16-December 5, 2021) and Omicron variant surge (December 27, 2021 to March 20, 2022). The proportion of change (Δ) in events (SARS-CoV2 cases, hospitalizations, and deaths), calculated as the relative risk (RR), the 95% confidence interval, and the p-value, during the Omicron variant surge was compared with the Delta variant surge . We are unable to compile the continuous data for December 5-26, 2021 period because the SARS-CoV2 cases, hospitalizations, and deaths for Nov 29-Dec 26, 2021 (rolling four weeks data published weekly), and the weekly rolling four weeks SARS-CoV2 hospitalizations data for January 3-January 30, 2022 period were not published by UKHSA in their weekly vaccine surveillance reports. However, aggregate of the four weeks outcomes (SARS-CoV2 cases, hospitalizations and deaths, vaccine effectiveness, CFR and RH) as presented in the supplemental appendix (Tables S6a-c, Table S7a and Tables S7c) and figures 6(a-d) for the four weeks period ending December 12, 2021 and December 19, 2021, thereby limiting the impact of the missing data only to one week from December 20, 2021 to December 26, 2021.

## RESULTS

### United Kingdom SARS-CoV2 cases, SARS-CoV2 hospitalizations and SARS-CoV2 deaths (Figure 1)

There was a total of 22,072,550 SARS-CoV2 cases, 848,911 hospitalizations, and 175,070 deaths reported in the UK during the entire pandemic as of May 1, 2022. The majority of cases (51.3%; n=11,315,793), the majority of hospitalizations (28.8%; n=244,708), and the second-highest percentage of deaths (16.4%; n=28,659) occurred in the twenty-one weeks period during the Omicron variant surge as per our study (December 6, 2021 to May 1, 2022).

**Figure 1:**
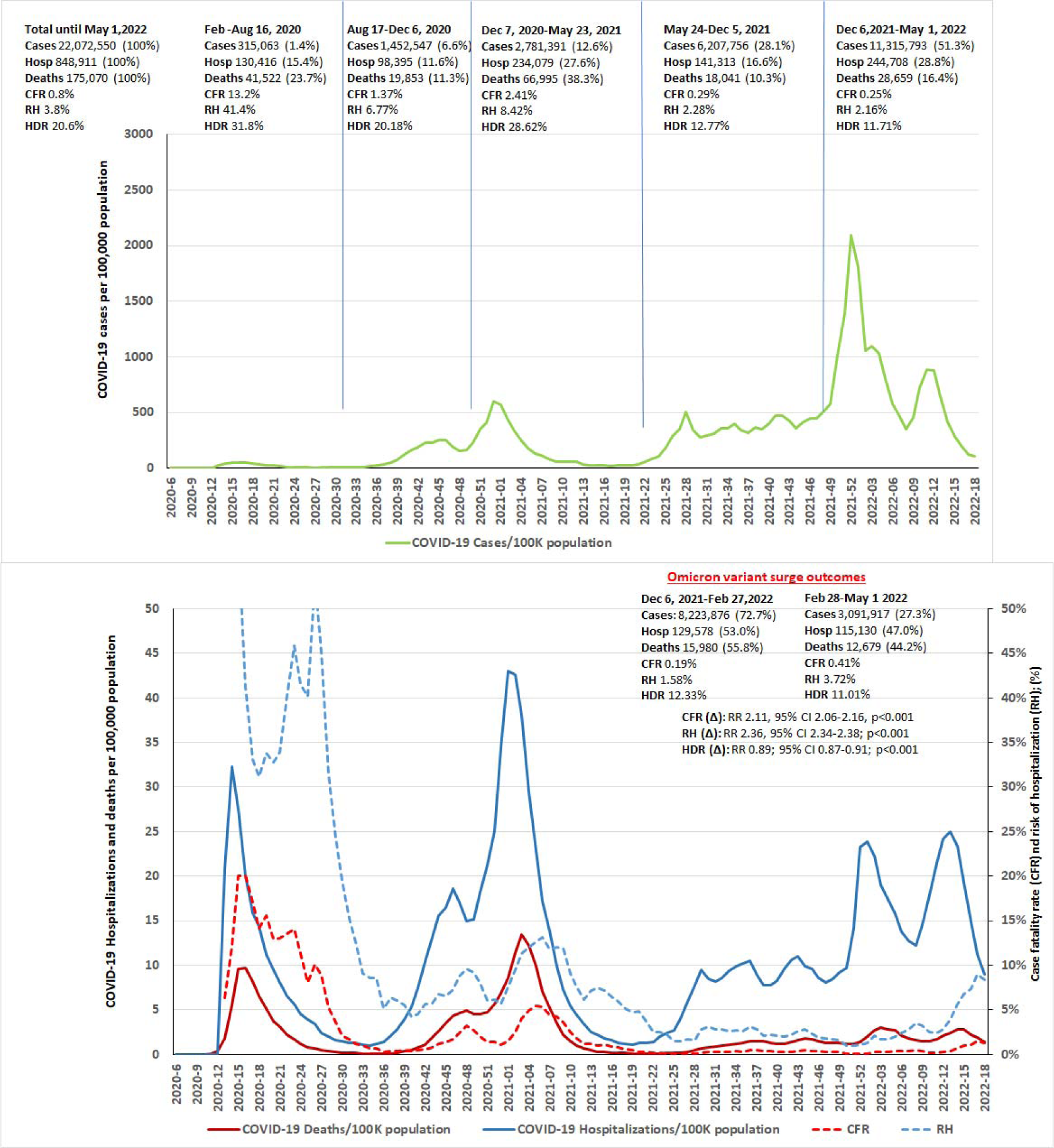
Total SARS-CoV2 cases, hospitalizations, and deaths in UK during the entire COVID-19 pandemic until May 1, 2022. The graph illustrates the increasing risk of the hospitalization (RH) and case fatality rate (CFR) during the latter part of the Omicron variant surge. The RH and CFR significantly increased during the latter part of the Omicron variant surge, (February 27-May 1, 2022) mainly driven by increased hospitalizations and deaths despite the lower number of cases. The changes in the hospitalization death rate (HDR) during the various time periods was also illustrated.

The case fatality rate (CFR) of 13.2% and risk of hospitalizations (RH) of 41.4% were highest during the first wave (February 24 - August 16, 2020). During the Alpha variant surge, the CFR was 2.41% with an RH of 8.42%. The CFR and RH decreased to 0.29% and 2.28%, respectively during the Delta variant surge and the lowest CFR of 0.25% and RH of 2.16% were observed during the Omicron variant surge. The CFR (0.19% vs. 0.41%; RR 2.11, 95% CI 2.06-2.16, p<0.001) and RH (1.58% vs 3.72%; RR 2.36, 95% CI 2.34-2.38; p<0.001) significantly increased during the 9 weeks period ending May 1, 2022 in the latter part of the Omicron variant surge compared to the earlier 12 weeks.

### SARS-CoV2 cases, SARS-CoV2 Hospitalizations (in England) and SARS-CoV2 deaths (England and Wales) among age groups (Figures 2a-c; Tables S1a-c)

**Figure 2a:**
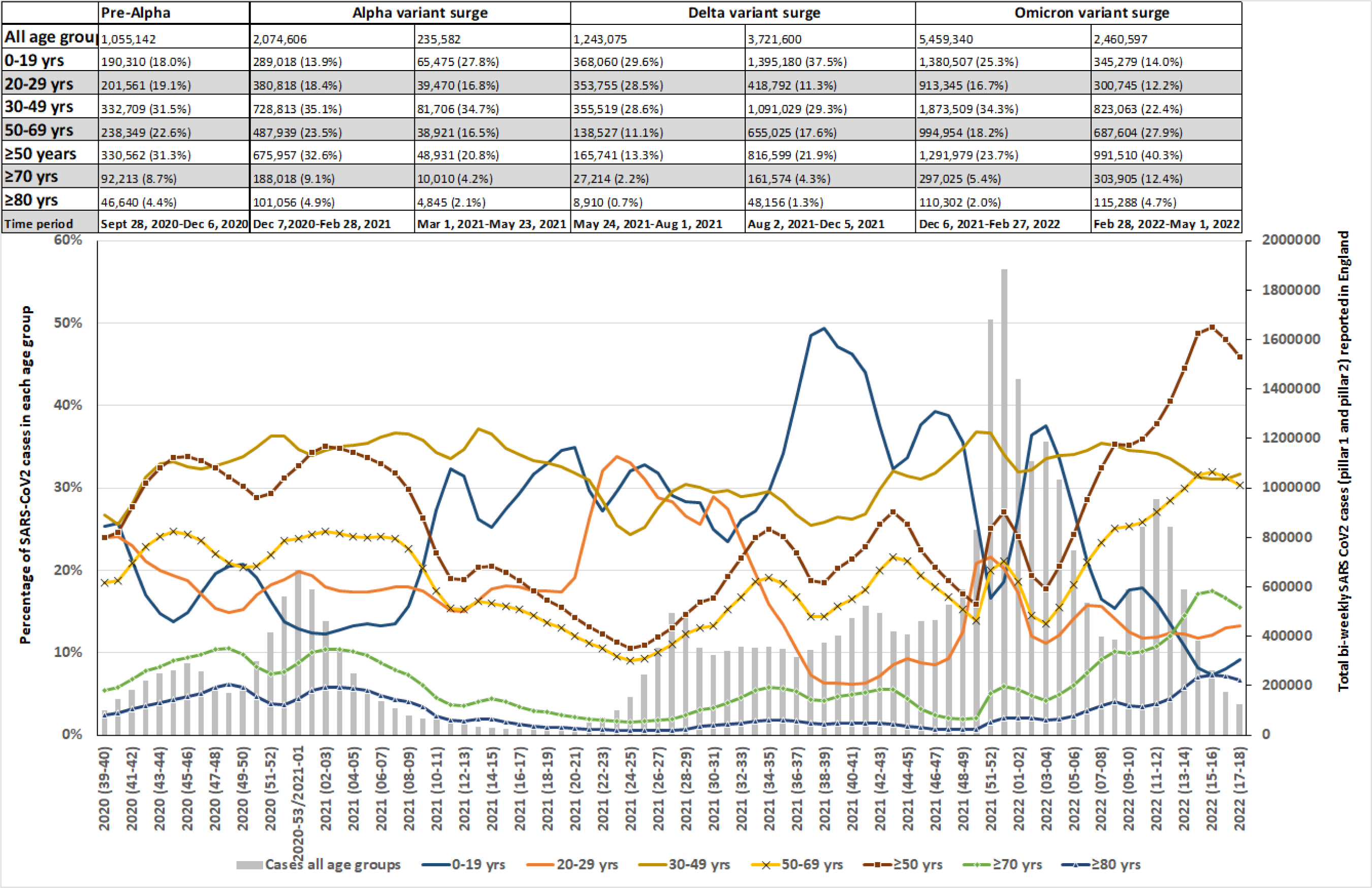
SARS-CoV2 cases (pillars 1 and 2) in England among various age groups in England from September 28, 2020 to May 1, 2022. The number of cases during the various surges is shown on the top panel for each age group (n=; %) and the percentage of two weekly cases among each age group is shown on the graph. As shown, there was an increased percentage of cases noted among ≥50 years of age groups of the total events compared to immediate prior period (that was noted to be statistically significant as shown in Table S1a) during the latter part of the Omicron variant surge during the February 28-May 1, 2022 period. The percentage of cases among the <50 years of age groups significantly decreased during the same period as shown in Table S1a.

**Figure 2b:**
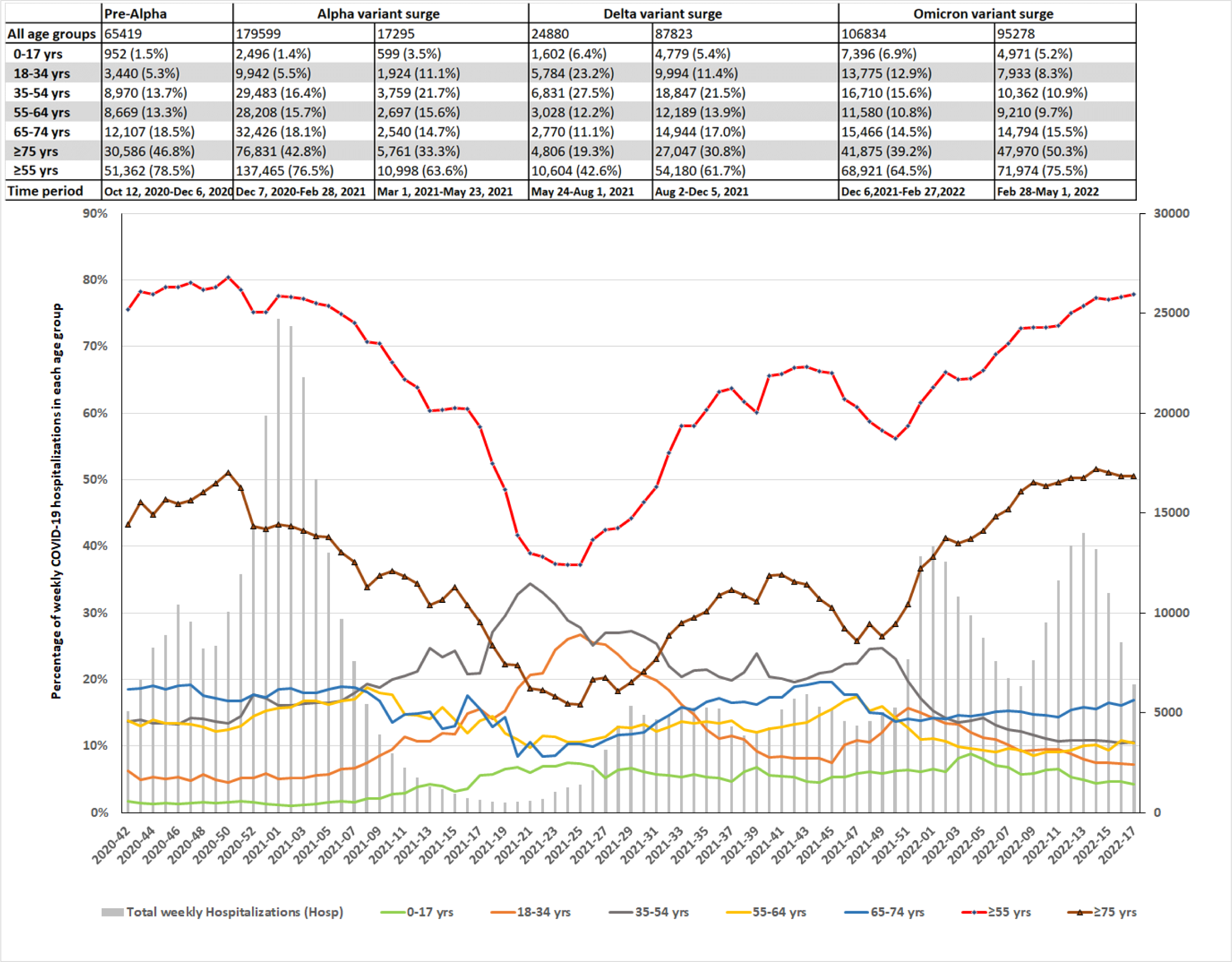
SARS-CoV2 hospitalization in England among various age groups in England from October 12, 2020 to May 1, 2022. The number of hospitalizations during the various surges is shown on the top panel for each age group (n=; %) and the percentage of weekly hospitalizations among each age group is shown on the graph. As shown, there was an increased percentage of hospitalizations noted among ≥75 years of age groups of the total events compared to immediate prior period (that was noted to be statistically significant as shown in Table S1b) during the latter part of the Omicron variant surge during the February 28-May 1, 2022 period. The percentage of hospitalizations among the <55 years of age groups significantly decreased during the same period as shown in Table S1b.

**Figure 2c:**
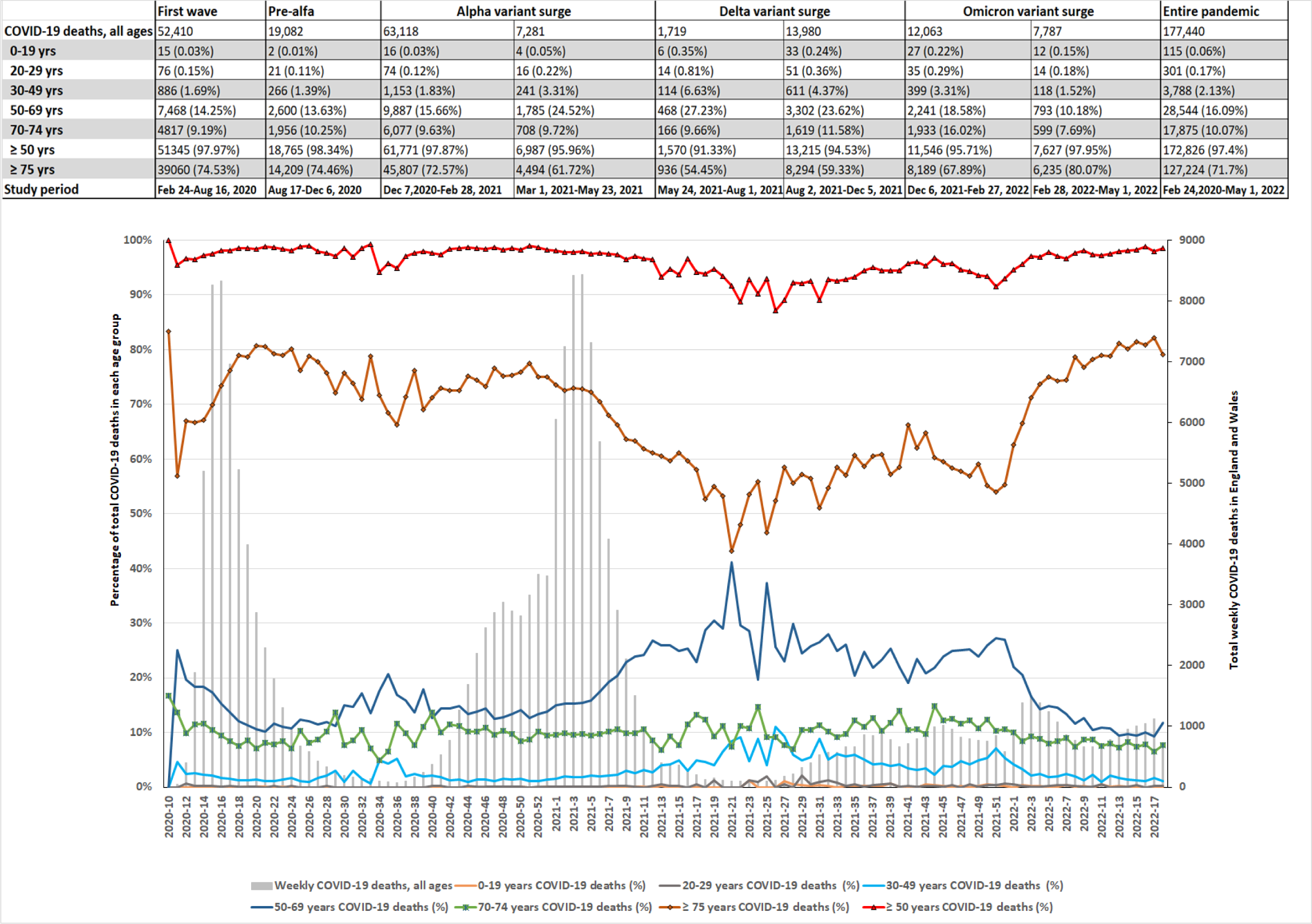
SARS-CoV2 deaths in England and Wales among various age groups from February 24, 2020 to May 1, 2022. The number of deaths during the various surges is shown on the top panel for each age group (n=; %) and the percentage of weekly deaths among each age group is shown on the graph. As shown, there was an increased percentage of deaths noted among ≥75 years of age groups of the total events compared to immediate prior period (that was noted to be statistically significant as shown in Table S1c) during the latter part of the Omicron variant surge during the February 28-May 1, 2022 period. The percentage of deaths among the <75 years of age groups significantly decreased during the same period as shown in Table S1c.

Of the reported total SARS-CoV2 cases in England, analysis of cases among various age groups showed a significantly increased percentage of cases among ≥50 years age group during the initial part of the Omicron variant surge compared to the prior period (21.9% vs 23.7%; RR 10.07; 95% CI 1.07-1.08);p<0.001) and during the latter part of Omicron variant surge (23.7% vs 40.3%; RR 1.70;95% CI 1.70-1.71;p<0.001) compared to the prior period as shown in Table S1a, particularly among ≥70 years of age. During the same periods of the reported total SARS-CoV2 hospitalizations in England, there were significantly increased hospitalizations among ≥75 years age group during the initial (30.8% vs 39.2%; RR 1.27;95% CI 1.25-1.29;p<0.001) and the latter period of Omicron variant surge (39.2% vs. 50.3%;RR 1.28;95% CI 1.27-1.30;p<0.001) compared to the immediate prior periods. The reported total SARS-CoV2 deaths in England and Wales during this time revealed significantly increased deaths during the initial part of the Omicron variant surge among 70-74 years (11.58% vs 16.02%; RR1.38;95%CI 1.30-1.47;p<0.001) and ≥75 years age group (59.33% vs 67.89%; RR1.14;95%CI 1.12-1.17;p<0.001) when compared to the immediate prior periods. During the latter part of the Omicron variant surge, ≥75 years age group have significantly increased deaths (67.89% vs 80.07%; RR1.18;95%CI 1.16-1.20;p<0.001) of the reported total SARS- CoV2 deaths. The 70-74 years age group showed significantly decreased deaths (16.02% vs 7.69%; RR 0.48;95% CI 0.44-0.52);p<0.001). The proportions of hospitalizations and deaths significantly decreased in <75 years age group with associated decreased cases in <50 years age group during the latter part of the Omicron variant surge of the reported total events.

### SARS-CoV2 cases, hospitalizations, ICU admissions and deaths among ethnic groups and deprived populations based on IMD score in England (Figures 3a-e)

**Figure 3a:**
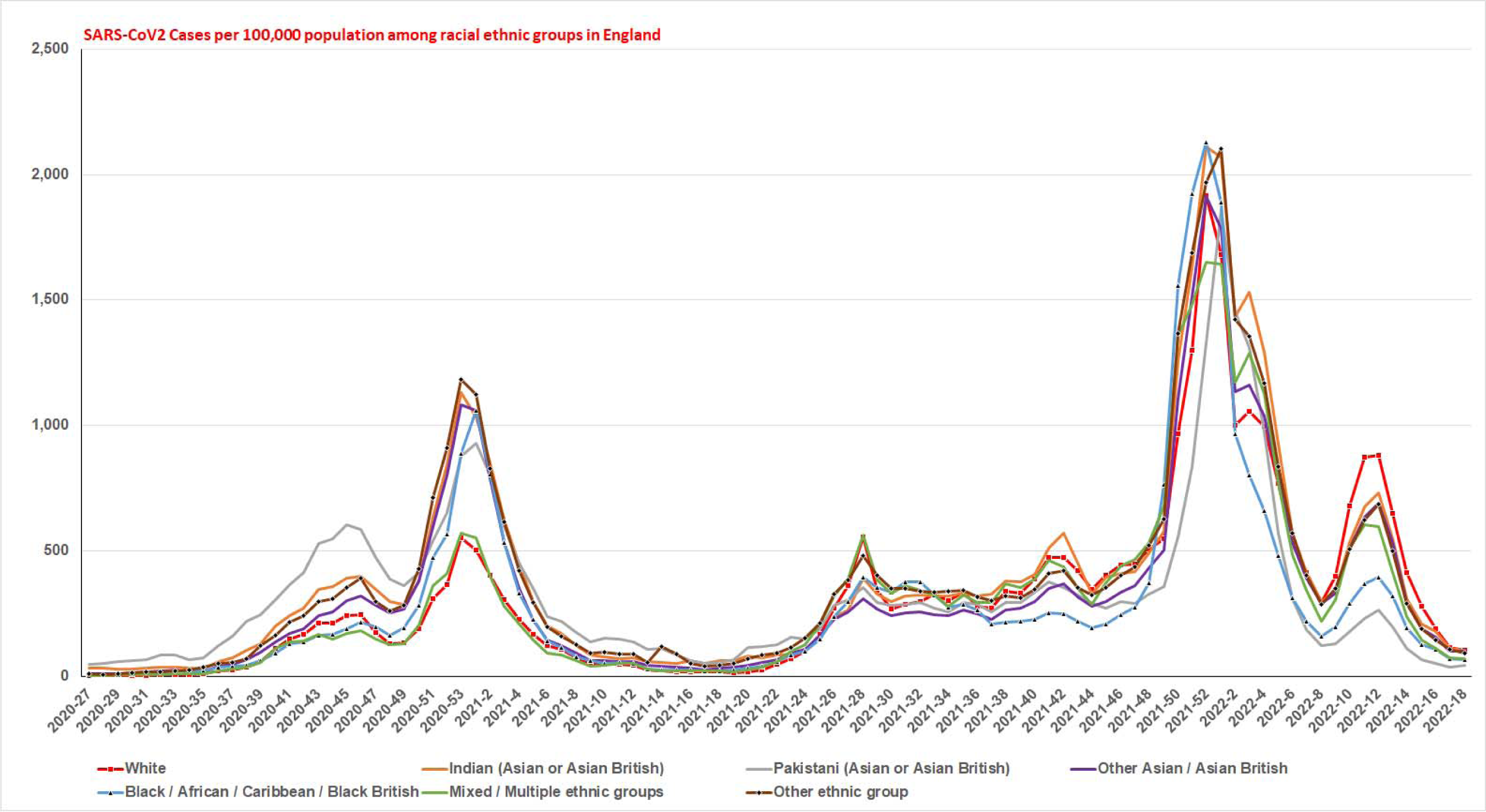
Weekly SARS-CoV2 cases (pillars 1 and 2) per 100,000 population among various ethnic groups in England from June 29, 2020 to May 1, 2022. As shown in the graph, racial minorities have higher infection rates period to and during the alpha variant surge, during the initial part of the Delta and Omicron variant surges than the white ethnic group. During the latter part of the Delta and Omicron variant surges, the white ethnic group have higher infection rates than other minorities.

**Figure 3b:**
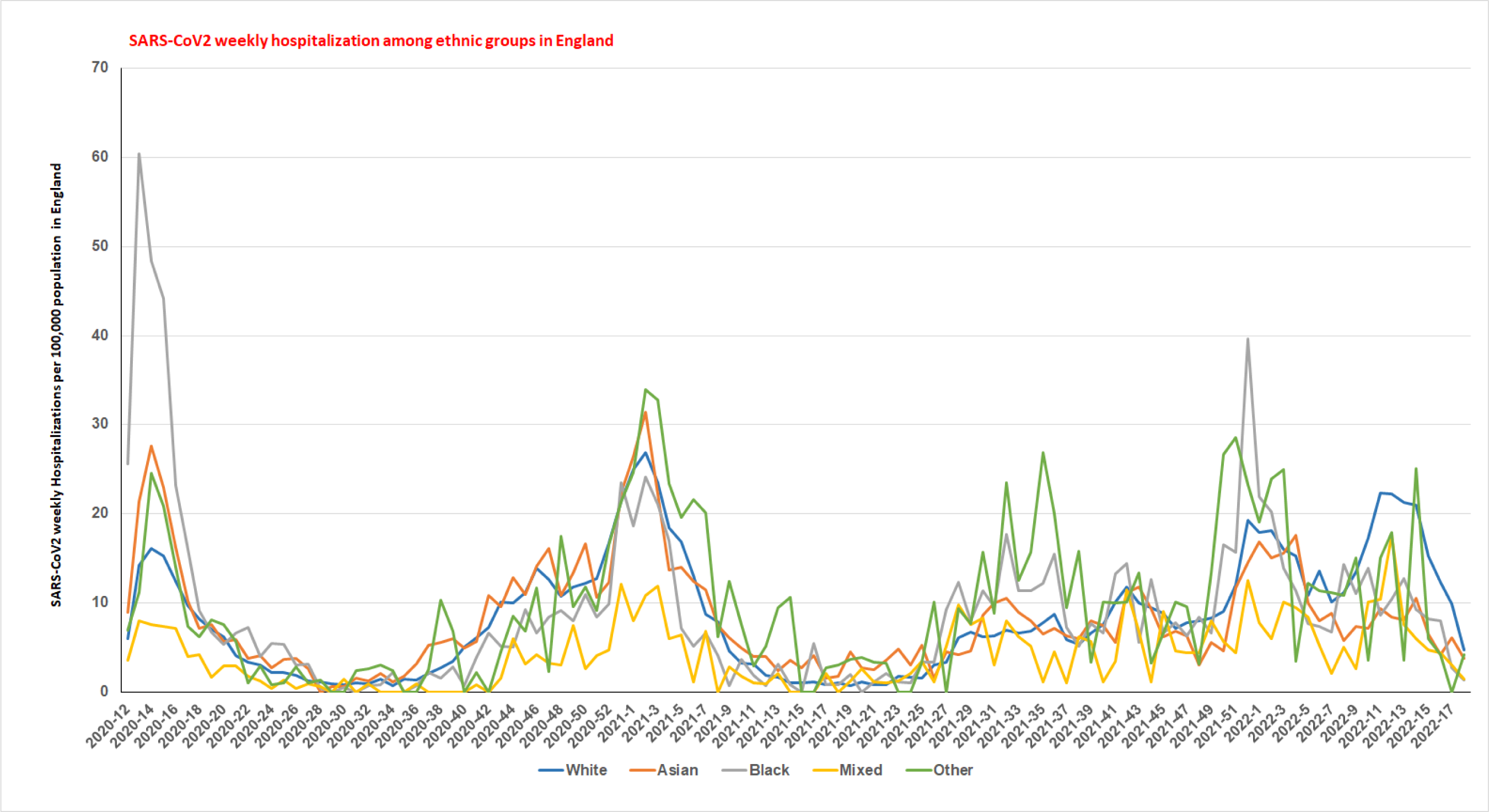
Weekly SARS-CoV2 hospitalizations per 100,000 population among various ethnic groups in England from March 16, 2020 to May 1, 2022. As shown the graph, racial minorities (black, Asian, mixed and other) have higher hospitalization rates during the First wave, the Alpha variant surge, and during the Delta variant surge. During the initial part of the Omicron variant surges racial minorities (black and other) have higher rates of hospitalization than the white ethnic group. During the latter part of the Omicron variant surges, the white ethnic group hospitalization rates increased.

**Figure 3c:**
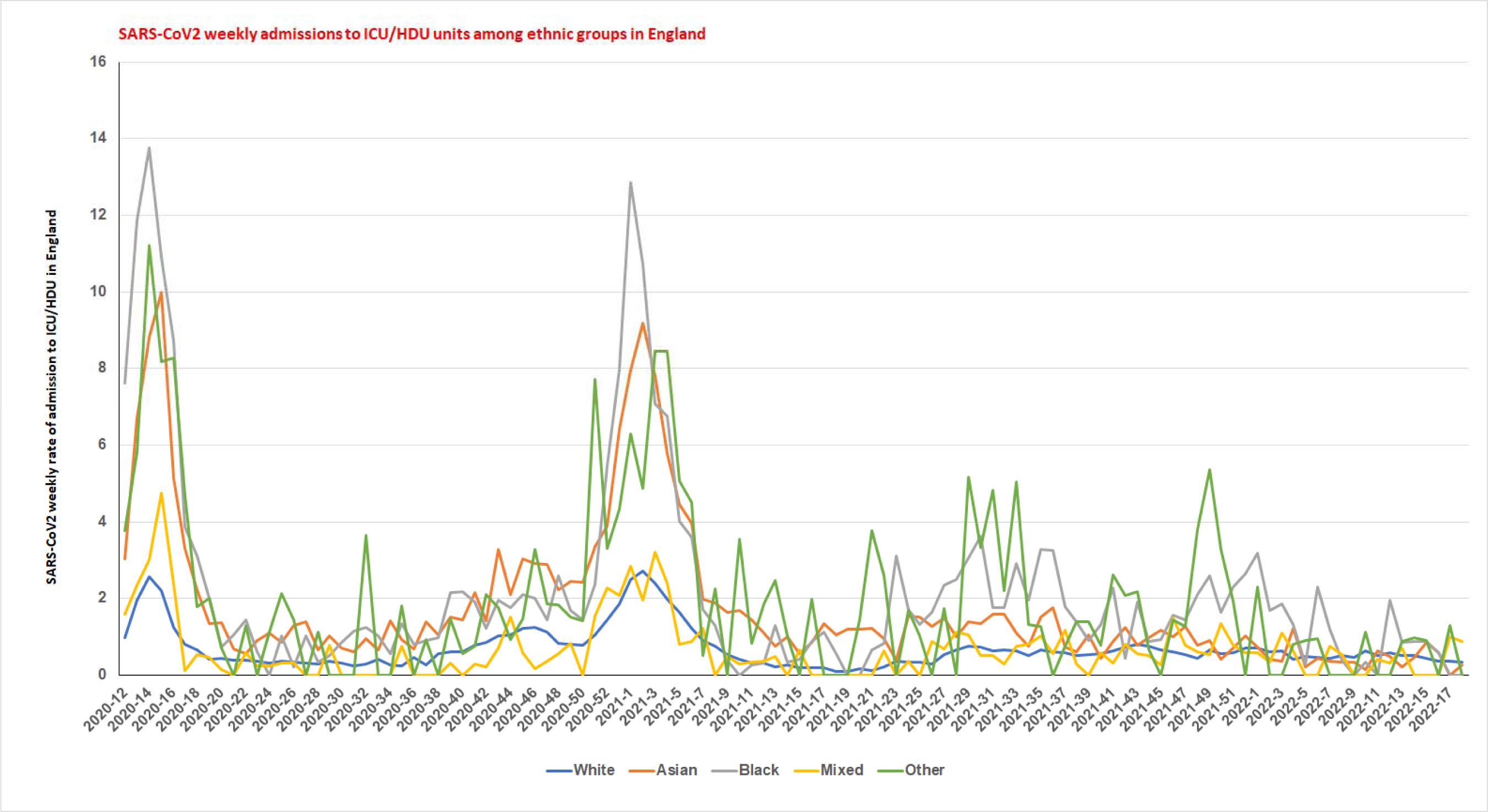
Weekly SARS-CoV2 admissions to ICU/HDU per 100,000 population among various ethnic groups in England during the March 16, 2020 to May 1, 2022. As shown, the racial minorities have higher rates of admission to ICU/HDU during the entire COVID-19 pandemic compared to the white ethnic group.

**Figure 3d:**
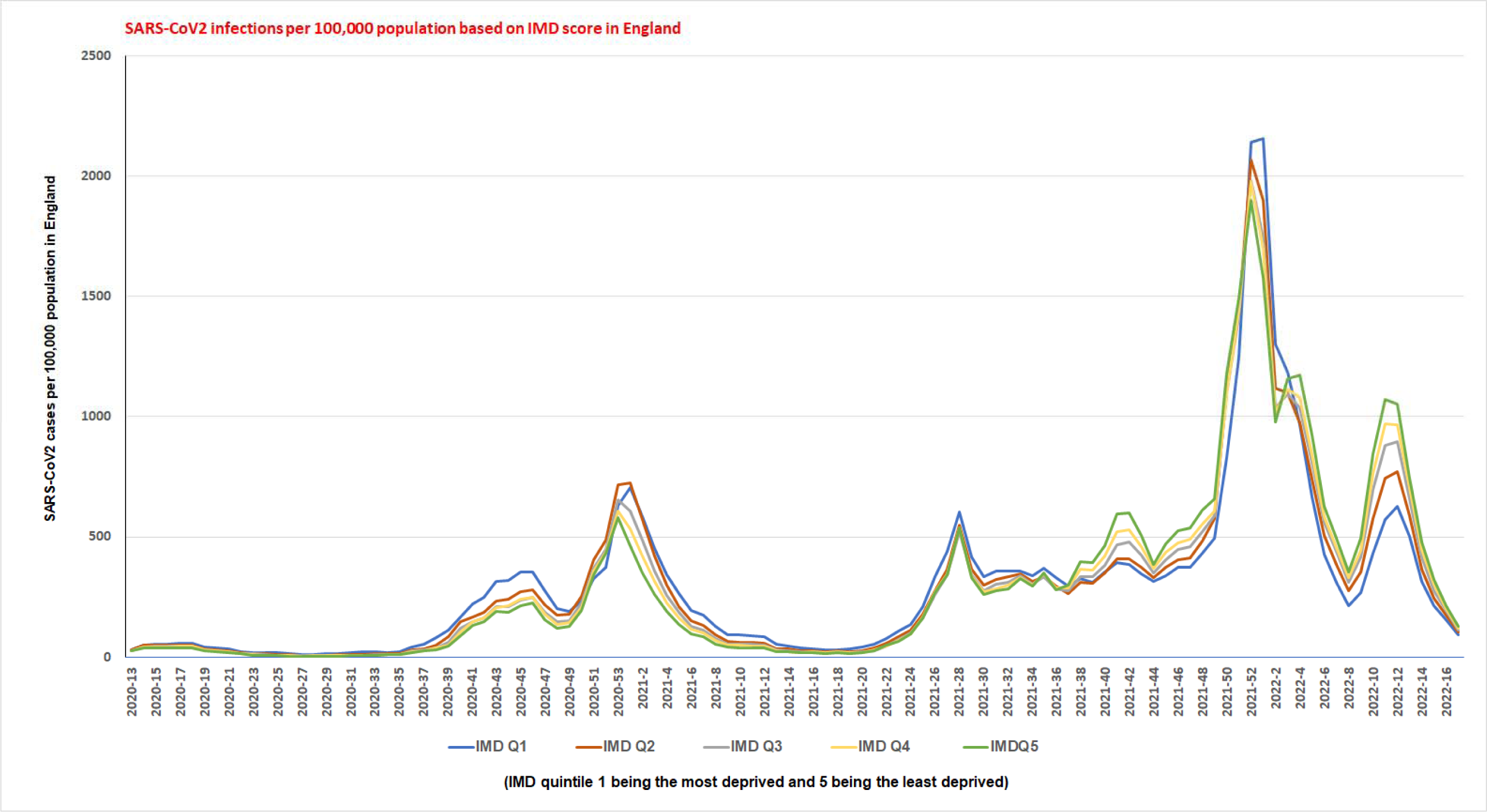
Weekly SARS-CoV2 cases (pillars 1 and 2) per 100,000 population based on the Indices of Multiple Deprivation (IMD) from March 16, 2020 to May 1, 2022. The infection rates were higher in most derived during the entire pandemic, except during the latter part of the Delta variant and particularly the latter part of the Omicron variant surge. During the latter part of the Omicron variant surge, the infection rates of the least deprived are higher than most deprived.

**Figure 3e:**
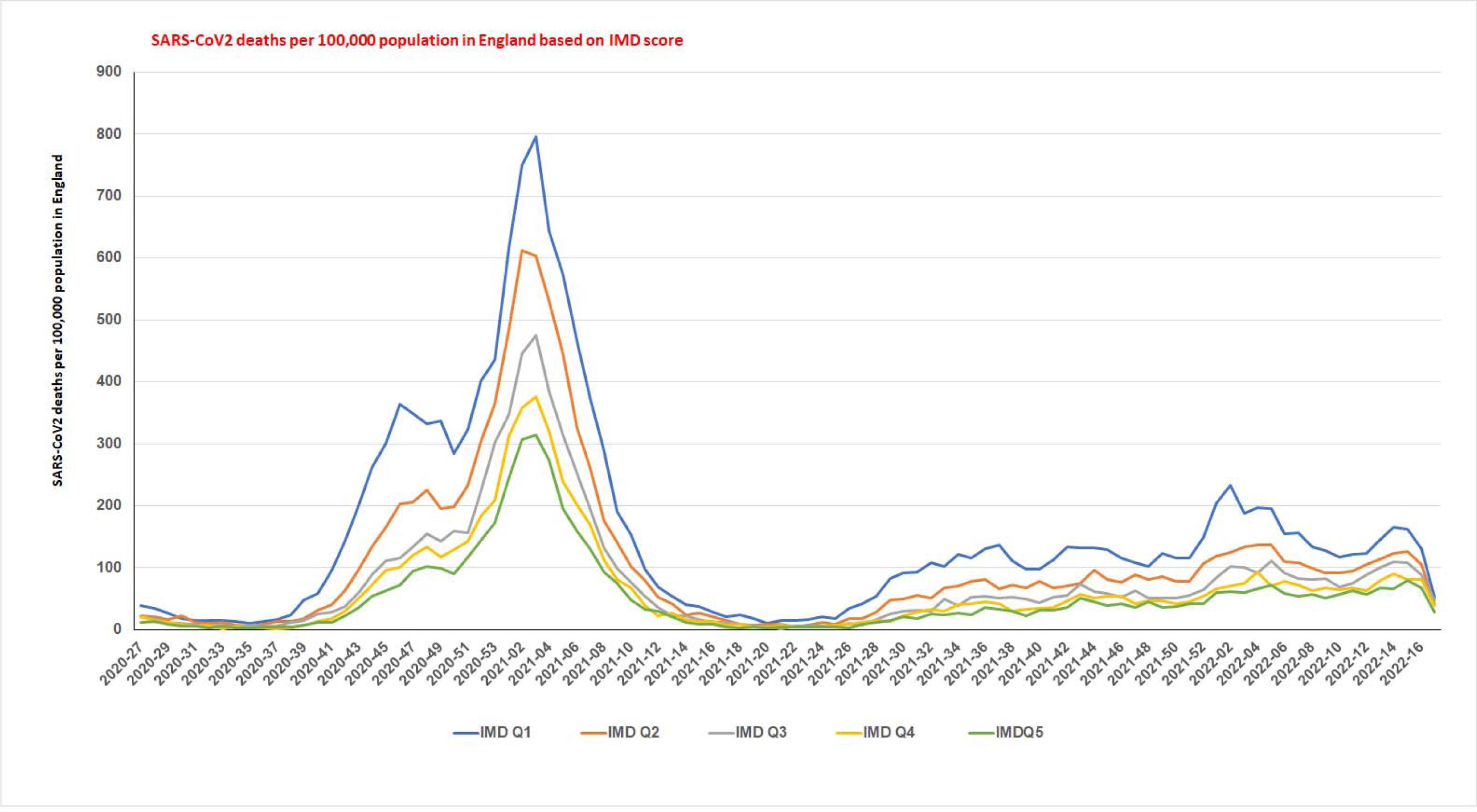
Weekly SARS-CoV2 deaths per 100,000 population based on the Indices of Multiple Deprivation (IMD) from March 16, 2020 to May 1, 2022. The death rates were higher in most derived during the entire pandemic including the latter part of the Omicron variant surge than the least deprived.

SARS-CoV2 cases and hospitalizations per 100,000 were highest in ethnic minorities than in the white ethnic group during and prior to the Alpha variant surge, the initial part of the Delta and Omicron variant surges. The case rates and hospitalization rates among the white ethnic group increased during the latter part of the Delta variant surge and the latter part of the Omicron variant surge from February 28-May 1, 2022.

However, the white ethnic group have lower ICU/HDU admission rates during the entire pandemic since March 2020. The cases per 100,000 population among the most deprived (IMD score) have higher case rates than the least deprived during the Alpha variant surge and the initial period of the Delta and Omicron variant surges. On the contrary, the least deprived have higher rates of cases during the latter part of the Delta variant surge and particularly during the latter part of the Omicron variant surge (February 28-May 1, 2022). However, the death rate among the most deprived is higher than the least deprived all throughout the pandemic, although the gap is narrowing during the latter part of the Omicron variant surge.

### SARS-CoV2 deaths among age groups in England and pre-existing conditions (Table S2)

The pre-existing conditions are present in 95.6% of COVID-19 deaths among all age groups (96.2% among those ≥60 years of age, 90.6% among 40-59 years, 82.8% among 20-39 years, and 69.2% of deaths among 0-19 years of age) during the Omicron variant surge.

### Vaccination rates among ethnic groups and population-based on deprivation score (IMD) as of May 1, 2022 **(Table S3b):**

According to the NIMS database of over 18 years, the British whites have the highest third dose vaccination rate of 76.4%, low rates of receiving two doses without a third dose (12.5%), and the lowest unvaccinated rate (8.9%). The population considered least deprived based on IMD decile score has the highest third dose vaccination rate (86.7%), low rates of two doses only without the third dose (10.2%), and the lowest unvaccinated rate (1.4%). Meanwhile, the most deprived have the lowest third dose vaccination rate (55.2%), higher rates of two doses without the third dose (25.2%), and non-vaccination rate (13.9%). The racial minorities have low rates of third dose vaccination (33-55%), high rates of not receiving the third dose after 2 doses (21.2%), and the highest rate of non-vaccination (28.8%).

### **Outcomes among** ≥**18 years of age NIMS vaccinated population in UK** (Figures 4-6, Table S3a, Tables S5a-c; Tables S6a-c, Tables S7a-c, Tables S8a-f)

**Figure 4:**
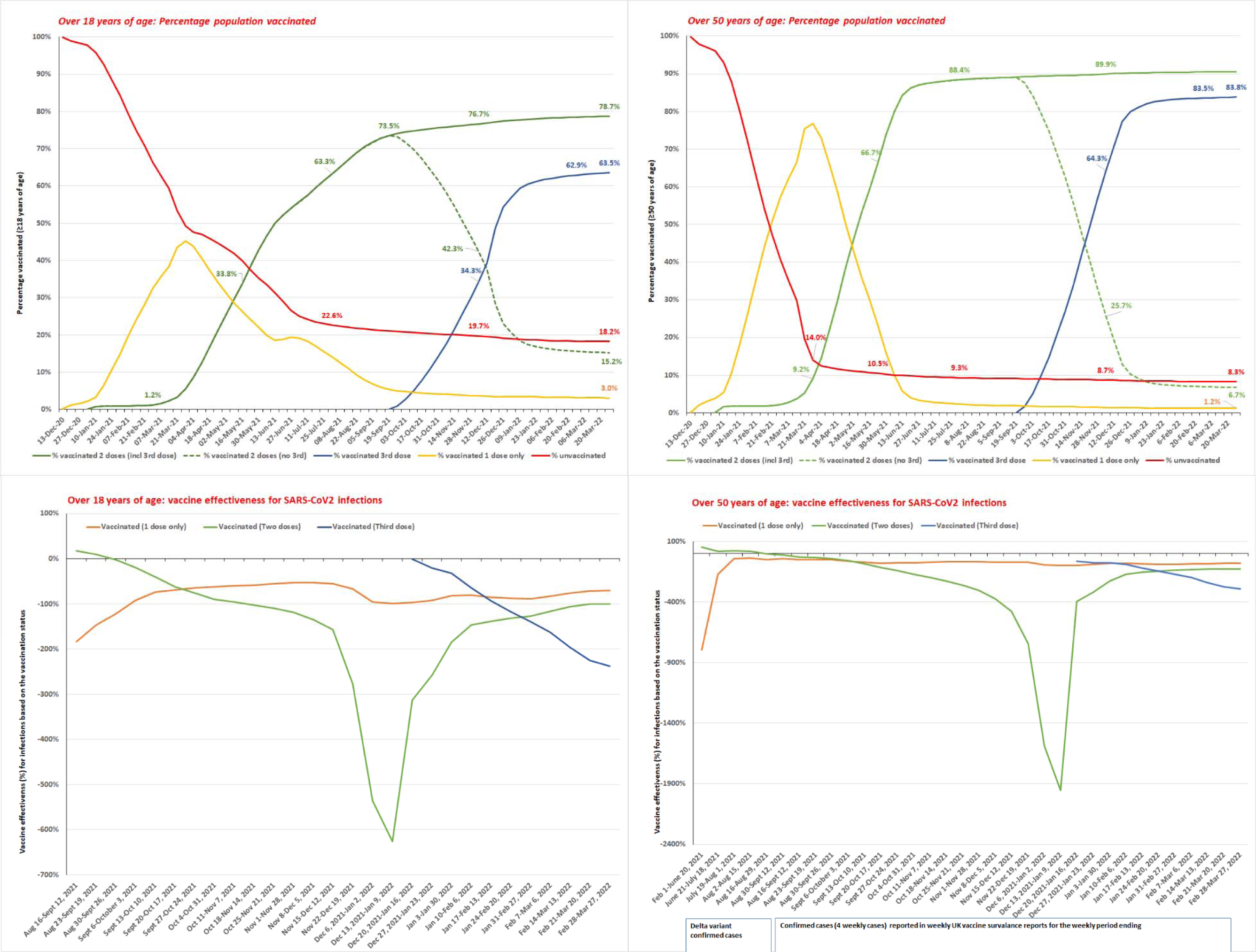
The vaccination rates of the over 18 years of age (top left), and over 50 years of age (top right); the vaccine effectiveness of over 18 years of age (bottom left) and over 50 years of age (bottom right) during the Delta variant sure and the Omicron variant surge until March 27, 2022. The top panel shows the percentage of the population vaccinated during the vaccination program weekly (for the week ending) until March 27, 2022. The bottom panel shows vaccine effectiveness for rolling four weekly cases from August 16, 2021 until March 27, 2022 (rolling 4 weeks ending date on x- axis). The ≥50 years of age Delta variant cases vaccine effectiveness (bottom right graph), the dates are listed on x-axis. Table S3a shows detailed vaccination rates of NIMS population (all ages, over 18 years and over 50 years of age groups) and Table S3b shows the vaccination rate disparities of the ethnic groups and vaccination disparities based on the IMD score. Table S7a-c shows the vaccine effectiveness (95% CI), of ≥18 years and ≥50 years of age NIMS population including the Delta variant cases vaccine effectiveness of ≥50 years of age. The SARS-CoV2 infections among the age groups based on the vaccination status, incidence rate per 100,000 population for the specified time period included the Table S7a (≥18 yrs. of age) and Tables S7b-c (≥50 yrs. of age). Th population denominator among the age groups based on the vaccination status for the specified time period included in the Table S3a.

**Figure 5a:**
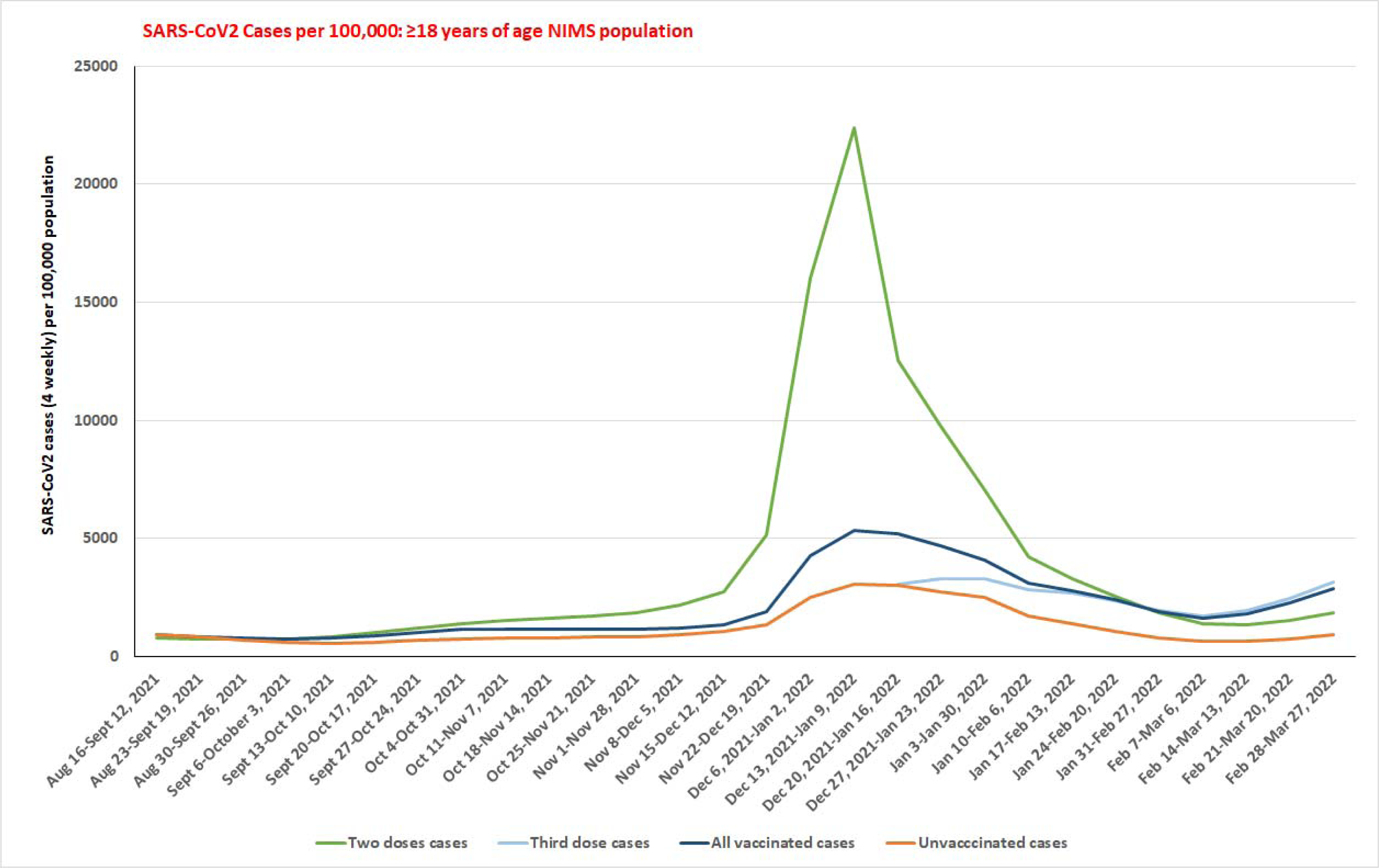
SARS-CoV2 cases per 100,000 population among the over 18 years of age group from August 16, 2021 to March 27, 2022. Table S8a shows that the vaccinated population (including two doses) have a significantly higher proportion of cases than the unvaccinated during the latter part of the Delta variant and initial part of Omicron variant surges. During the latter part of the Omicron variant surge, the vaccinated with the third dose have the highest proportion of infection than those vaccinated with two doses and unvaccinated. The SARS-CoV2 infections based on the vaccination status for the specified time period included the Table S6a. The population denominator for ≥18 years of age based on the vaccination status for the specified time period included in the Table S3a.

**Figure 5b:**
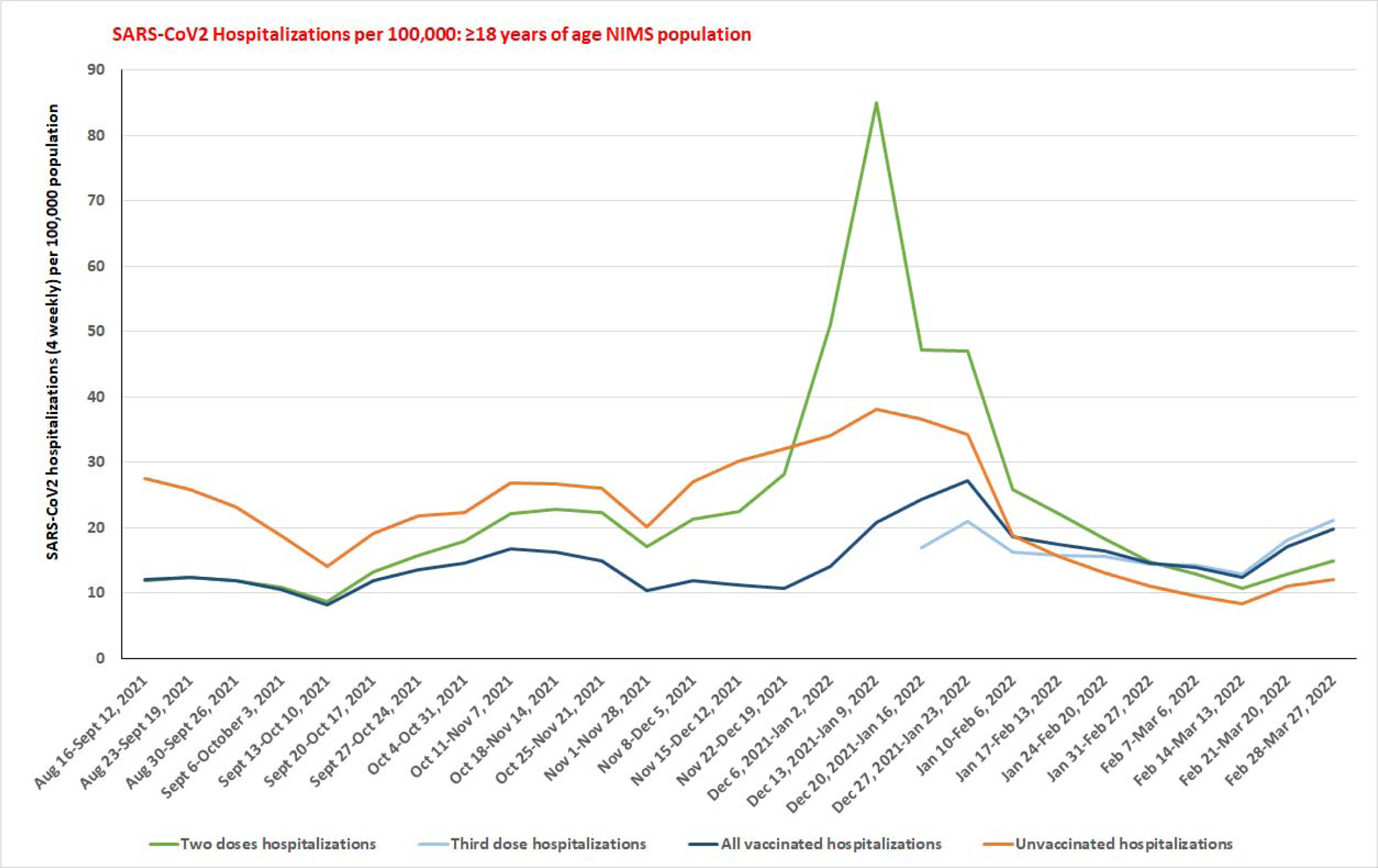
SARS-CoV2 hospitalizations per 100,000 population among the over 18 years of age group from August 16, 2021 to March 27, 2022. Table S8b shows that the vaccinated with two doses have a significantly higher proportion of hospitalizations than the unvaccinated during the initial part of the Omicron variant surge. During the latter part of the Omicron variant surge, the vaccinated with the third dose (including all vaccinated population) have the highest proportion of hospitalizations than those vaccinated with two doses and unvaccinated. The SARS-CoV2 hospitalizations based on the vaccination status for the specified time period included the Table S6b. The population denominator for ≥18 years of age based on the vaccination status for the specified time period included in the Table S3a.

**Figure 5c:**
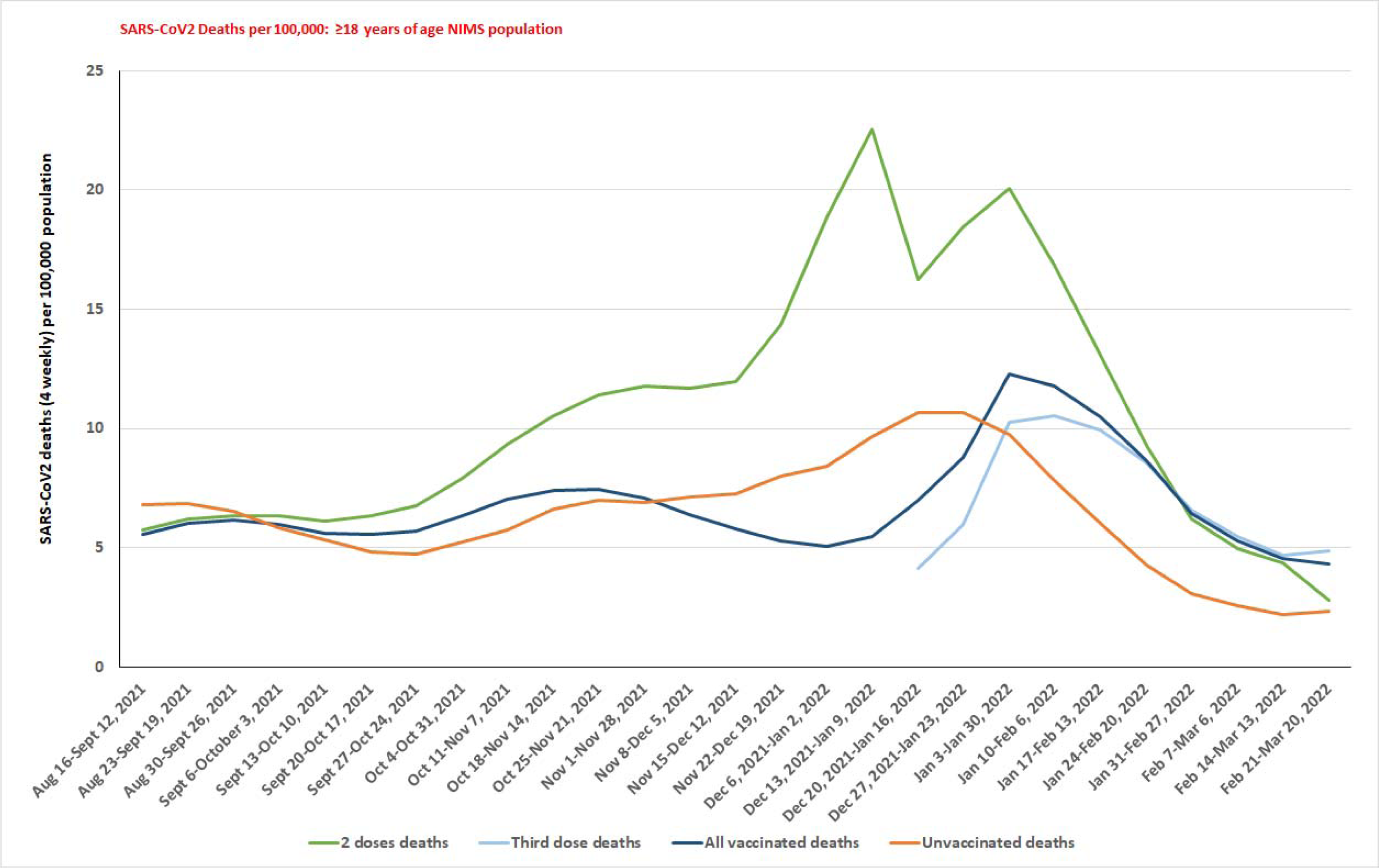
SARS-CoV2 deaths per 100,000 population among the over 18 years of age group from August 16, 2021 to March 27, 2022. Table S8c shows that the vaccinated population with two doses has a significantly higher proportion of hospitalizations than the unvaccinated during the latter part of the Delta variant surge and the initial part of the Omicron variant surge. During the latter part of the Omicron variant surge, the vaccinated with the third dose (including all vaccinated population) have the highest proportion of deaths than those vaccinated with two doses and unvaccinated. The SARS-CoV2 deaths based on the vaccination status for the specified time period included the Table S6c. The population denominator for ≥18 years of age based on the vaccination status for the specified time period included in the Table S3a.

**Figure 6a:**
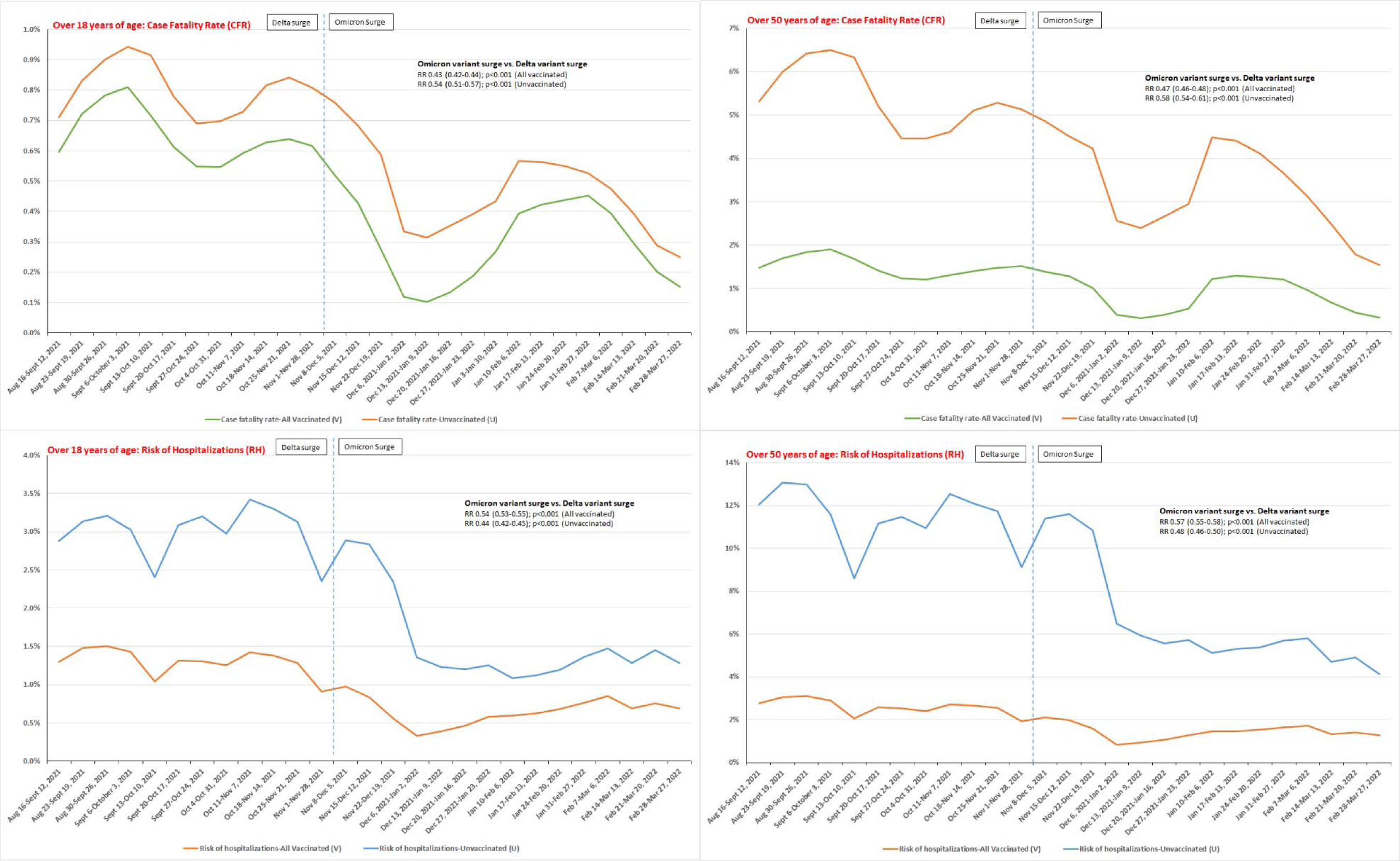
The case fatality rate (CFR) of over 18 years (top left), over 50 years (top right) of age, the Risk of Hospitalizations (RH) of over 18 years (bottom left), over 50 years (bottom right) of age populations. The CFR or RH presented on the y-axis and study period (rolling four weekly period from August 16, 2021 to March 27, 2022) on the x- axis of All vaccinated population (those who received at least one dose) and unvaccinated population of the respective age. The proportion of change (Δ) in events (SARS-CoV2 cases, hospitalizations, and deaths), calculated as the relative risk (RR), the 95% confidence interval, and the p-value, during the Omicron variant surge (December 27, 2021 to March 20, 2022) was compared with the Delta variant surge (August 16-December 5, 2021) as presented in the Table 1a-c.

**Figure 6b:**
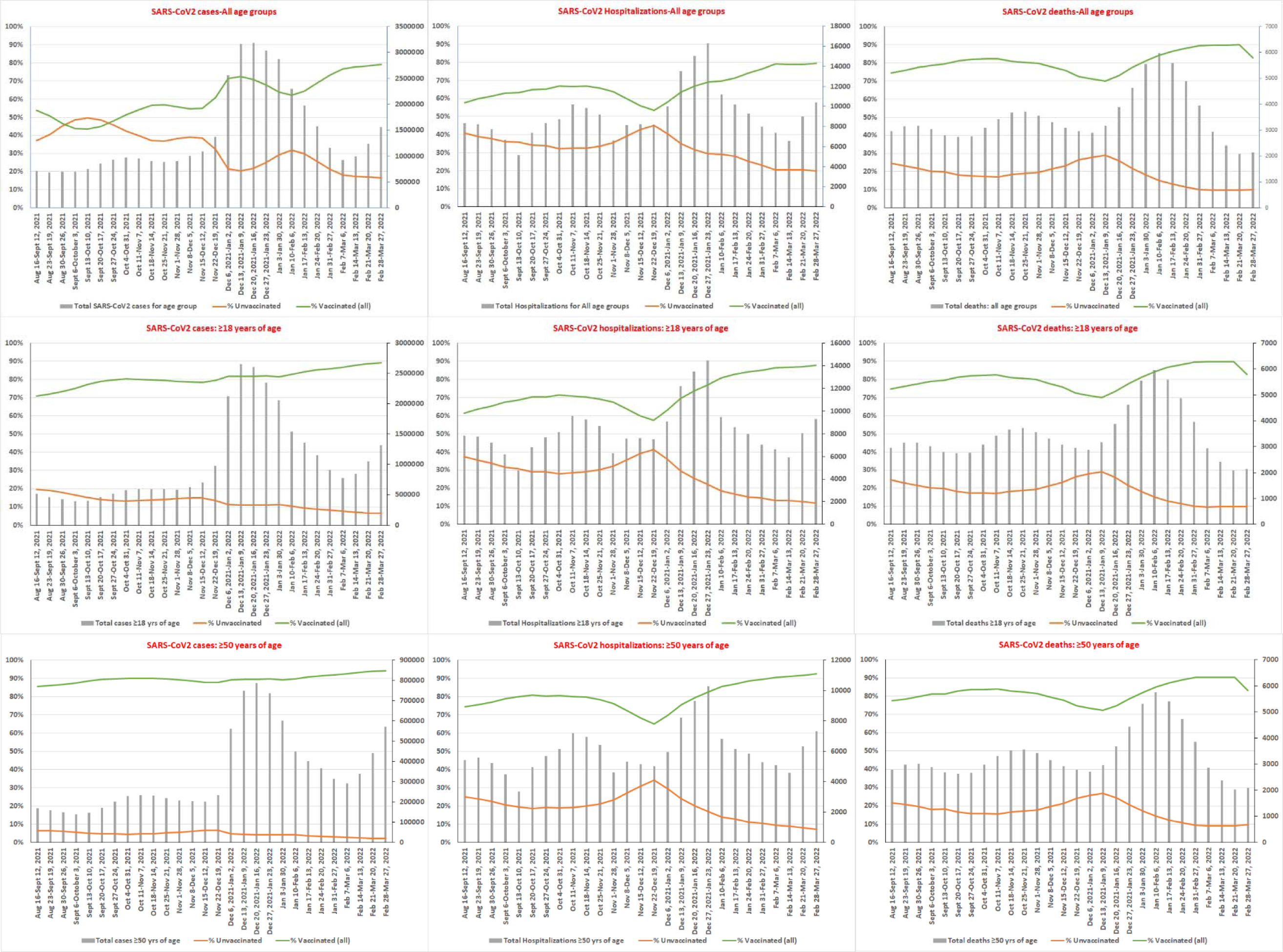
The percentage of SARS-CoV2 cases (left), hospitalizations (middle) and deaths (right) based on the vaccination status among all age groups (top row) over 18 years of age (middle row) and over 50 years of age (bottom row) reported from the NIMS database from August 16, 2021 to March 27, 2022. The detailed rolling four weekly changes the percentage of SARS-CoV2 cases (Table S6a), SARS-CoV2 hospitalizations (Table S6b) and SARS-CoV2 deaths (Table S6c) are shown the tables. The increased SARS-CoV2 cases among the vaccinated population in each age group during the Omicron variant surge is associated with increased SARS-CoV2 hospitalizations and SARS-CoV2 deaths in the vaccinated population. The decreased SARS-CoV2 cases among the unvaccinated population in each age group during the Omicron variant surge is associated with decreased SARS-CoV2 hospitalizations and SARS-CoV2 deaths in unvaccinated. The proportionality test results detailed in Tables S8(a-f); show that a significantly higher proportion of SARS-CoV2 cases in the vaccinated population (including the third dose) than in unvaccinated during the Omicron variant surge was associated with a significantly higher proportion of SARS-CoV2 hospitalizations in all vaccinated population (including the third dose) than the unvaccinated among ≥18 years of age and ≥50 years of age during the latter part of the Omicron variant surge. The proportionality test also shows a significantly higher proportion of deaths among the ≥18 years of age vaccinated population (including the third dose) than unvaccinated during the latter part of Omicron variant surge. The proportionality test also showed higher proportion of deaths among ≥50 years of age vaccinated with two doses than unvaccinated during the Omicron variant surge with the third dose population deaths lower than unvaccinated until March 27, 2022. The population denominator and the vaccination rates for the specified time period among the studied age groups included in the Table S3a.

**Figure 6c:**
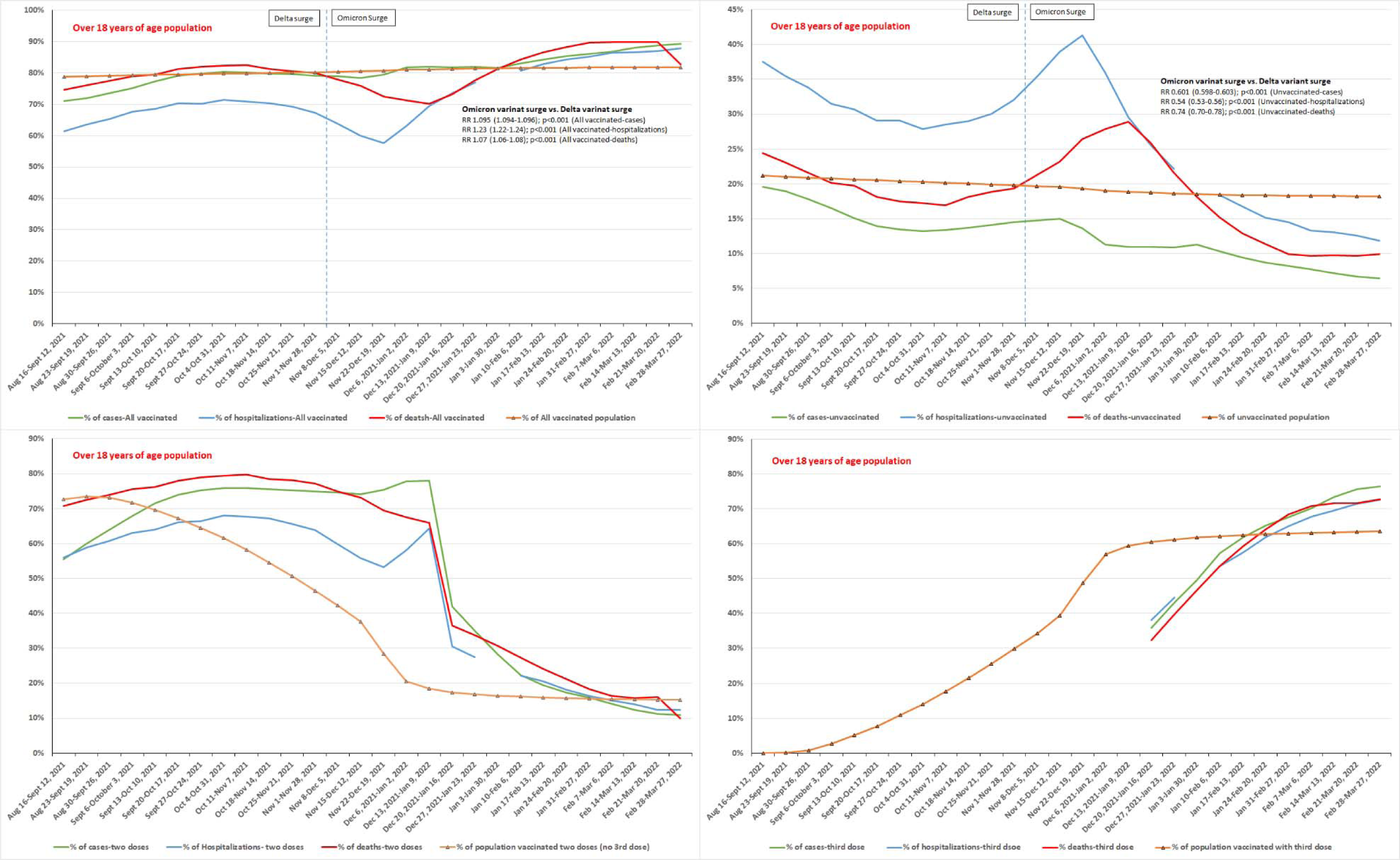
The real-world percentage of SARS-CoV2 cases, hospitalizations, and deaths among over 18 years of age population based on the vaccination status (All vaccinated-those vaccinated with at least one dose [upper left], unvaccinated [upper right], vaccinated with third dose [bottom left] and vaccinated with two doses only [bottom right] from August 16, 2021 to March 27, 2022. The percentage of SARS-CoV2 cases, hospitalizations, deaths and the percentage of population vaccinated (All vaccinated- those vaccinated with at last one dose, vaccinated with two doses or third dose) or unvaccinated population presented on the y-axis and study period (rolling four weekly period from August 16, 2021 to March 27, 2022) on the x-axis. The proportion of change (Δ) in events (SARS-CoV2 cases, hospitalizations, and deaths), calculated as the relative risk (RR), the 95% confidence interval, and the p-value, during the Omicron variant surge (December 27, 2021 to March 20, 2022) was compared with the Delta variant surge (August 16-December 5, 2021) as presented in the Table 1b.

**Figure 6d:**
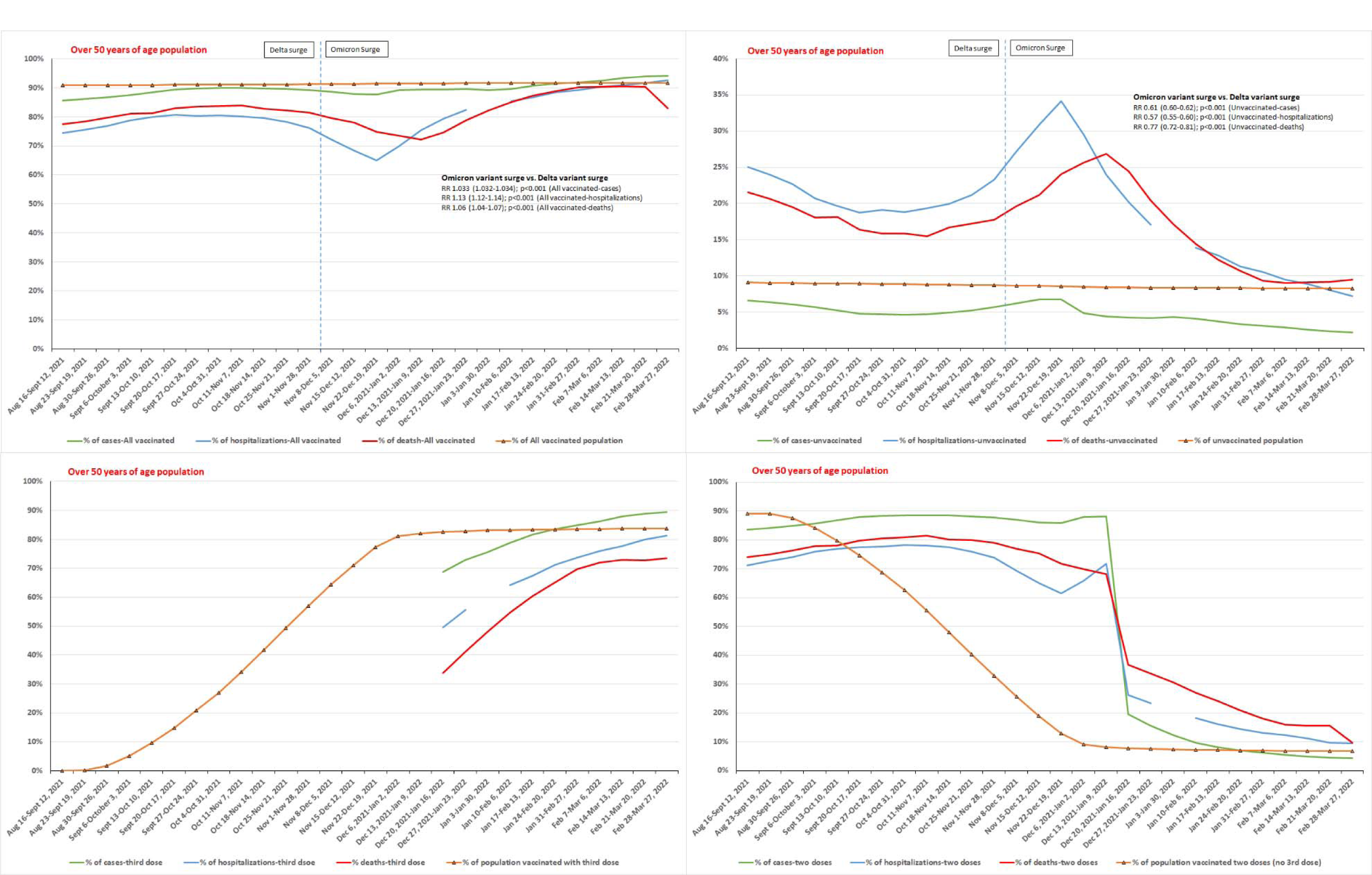
The real-world percentage of SARS-CoV2 cases, hospitalizations, and deaths among over 50 years of age population based on the vaccination status (All vaccinated-those vaccinated with at least one dose [upper left], unvaccinated [upper right], vaccinated with third dose [bottom left] and vaccinated with two doses only [bottom right] from August 16, 2021 to March 27, 2022. The percentage of SARS-CoV2 cases, hospitalizations, deaths and the percentage of population vaccinated (All vaccinated- those vaccinated with at last one dose, vaccinated with two doses or third dose) or unvaccinated population presented on the y-axis and study period (rolling four weekly period from August 16, 2021 to March 27, 2022) on the x-axis. The proportion of change (Δ) in events (SARS-CoV2 cases, hospitalizations, and deaths), calculated as the relative risk (RR), the 95% confidence interval, and the p-value, during the Omicron variant surge (December 27, 2021 to March 20, 2022) was compared with the Delta variant surge (August 16-December 5, 2021) as presented in the Table 1c.

The data published in UK vaccine surveillance reports (August 16, 2021-March 27, 2022) was used for the analysis. The ≥18 years of age, two-doses vaccinated population showed vaccine effectiveness of 17.5% (95% CI 16.5%-18.4%) for the four weeks period ending September 12, 2022. Ever since the vaccinated with two doses and the third dose (since December 20, 2021) have negative vaccine effectiveness, implying that the vaccinated population has a greater proportion of cases versus the unvaccinated population. The UKHSA did not report the third dose outcomes until December 20, 2021 in their weekly reports. In our study, we analyzed the 336,046 cases among the third booster pillar 2 population (November 27, 2021-January 12, 2022). Using the third dose weekly vaccinated population between December 26, 2021 to January 9, 2022, we observed that the best vaccine effectiveness values for the third dose were13.8%, 18.4%, and 22.2%, respectively. The two-proportions test showed that the all-vaccinated population have higher proportion of cases than the unvaccinated since August 23, 2021 (*X*^2^ = 11.686, df = 1, p = 0.0003149) and since the third dose reporting period of December 20, 2021 (*X*^2^= 79651, df = 1, p<0.001). Of the vaccinated populations, the vaccinated with two doses have a significantly higher proportion of cases since August 30, 2021. Since January 31, 2022, the third dose vaccinated population have a significantly higher proportion of cases than the population with two doses (*X*^2^= 368.53, df = 1, p<0.001) and the unvaccinated population (*X*^2^= 55147, df = 1, p<0.001). The two-proportions test also showed that the over 18 years old, two doses vaccinated population have a significantly higher proportion of hospitalizations than the unvaccinated (*X*^2^= 332.3, df = 1, p<0.001) since December 6, 2021. Since January 17, 2022, the vaccinated population has a significantly higher proportion of hospitalizations than the unvaccinated (*X*^2^= 14.103, df = 1, p<0.001). Since January 24, 2022, the third dose population also has a higher proportion of hospitalizations than the unvaccinated (*X*^2^= 33.446, df = 1, p<0.001). Since February 14, 2022 the third dose population has a higher proportion of hospitalizations than the two-doses (*X*^2^= 25.946, df = 1, p<0.001) and unvaccinated (*X*^2^= 121.11, df = 1, p<0.001) populations. The two-proportions test on deaths also showed a significantly higher proportion of deaths among the ≥18 years of age, two doses vaccinated versus the unvaccinated since September 13, 2021 (*X*^2^= 7.6872, df = 1, p = 0.0028). The all-vaccinated population has a higher proportion of deaths versus the unvaccinated since January 10, 2022 (*X*^2^= 41.017, df = 1, p<0.001), and the third dose population has a higher proportion of deaths (*X*^2^= 53.465, df = 1, p<0.001) than the unvaccinated since January 17, 2022.

Since, February 14, 2022, the vaccinated population with the third dose had a higher proportion of deaths versus the vaccinated with two doses (*X*^2^= 2.8687, df = 1, p p<0.001) and versus the unvaccinated (*X*^2^= 122.59, df = 1, p<0.001). The significantly increased proportion of cases in the vaccinated population during the Omicron variant was associated with significantly increased hospitalization and deaths among the vaccinated population of ≥18 years of age. On the contrary, the unvaccinated, ≥18 years of age population observed a significantly decreased cases, hospitalizations, and deaths. The analysis of the Case Fatality Rate (CFR) and the Risk of Hospitalizations (RH) as shown in Table 1a-c and Figure 6a showed a significantly decreased CFR among all persons (0.58% vs 0.25% [RR 0.44 (0.43-0.45); p<0.001]), unvaccinated (0.77% vs 0.41% [RR 0.54 (0.51-0.57]; p<0.001), all vaccinated (0.59% vs 0.25% [RR 0.43 (0.42-0.44); p<0.001]), and vaccinated with two doses (0.63% vs 0.26% [RR 0.41 (0.39-0.42); p<0.001]) populations of over 18 years of age during the Omicron variant surge (December 27, 2021 to March 20, 2022) compared to the Delta variant surge (August 16-December 5, 2021). Similarly, the RH was significantly decreased among all persons (1.38% vs 0.67%; [RR 0.484 (0.476-0.492); p<0.001]), unvaccinated (2.92% vs 1.27%; [RR 0.44 (0.42-0.45); p<0.001]), all-vaccinated (1.19% vs 0.65%; [RR 0.54 (0.53-0.55); p<0.001]), and vaccinated with two doses (1.23% vs 0.56%; [RR 0.46 (0.44-0.47); p<0.001]) populations of over 18 years of age during the Omicron variant surge compared to Delta variant surge. As shown in Table 1b, the significantly decreased SARS-CoV2 cases in unvaccinated [334,626 of 2,144,674 (15.6%) vs. 426,642 of 4,550,426 (9.4%); RR 0.601 (0.598-0.603); p<0.001] during the Omicron variant surge (December 27, 2021 to March 20, 2022) among over 18 years of age was associated with significantly decreased hospitalizations [9,785 of 29,680 (33.0%) vs 5,434 of 30,480 (17.8%); RR 0.54 (0.53-0.56); p<0.001] and significantly decreased SARS-CoV2 deaths [2,561 of 12,486 (20.5%) vs 1,757 of 11,584 (15.2%); RR 0.74 (0.70-0.78); p<0.001] versus the Delta variant surge (August 16-December 5, 2021). Similarly, a significantly increased SARS-CoV2 cases among the all vaccinated population [1,651,994 of 2,144,674 (77.0%) vs 3,839,525 of 4,550,426 (84.4%); RR 1.095 (1.094-1.096); p<0.001] during the Omicron variant surge was associated with significantly increased SARS-CoV2 hospitalizations [19,630 of 29,680 (66.1%) vs 24,843 of 30,480 (81.5%); RR 1.23 (1.22-1.24) p<0.001] and significantly increased SARS-CoV2 deaths [9,827 of 12,486 (78.7%) vs 9,760 of 11,584 (84.3%); RR 1.07 (1.06-1.08); p<0.001 compared with the Delta variant surge.

**Tables 1a:** SARS-CoV2 cases, SARS-CoV2 hospitalizations, SARS-CoV2 deaths, the case fatality rate, risk of the hospitalizations and the hospitalization deaths rate of the NIMS population of all ages (entire population) during the Omicron variant surge compared with the Delta variant surge.

| Outcomes/<br>Vaccination status | Delta variant surge<br>(August 16-December 5,<br>2021) | Omicron variant surge<br>(December 27, 2021 to March<br>20, 2022) | Statistical analysis ($\Delta$ from prior<br>period) [RR (95% CI); p value] |
| --- | --- | --- | --- |
| **Total SARS-CoV2 cases (All age groups-Entire NIMS population)** |  |  |  |
| All persons | 3,409,981 | 5,850,110 |  |
| Unvaccinated | 1,403,100 (41.1%) | 1,378,430 (23.6%) | RR 0.567 (0.566-0.568); p<0.001 |
| All Vaccinated | 1,765,718 (51.8%) | 4,115,729 (70.4%) | RR 1.344 (1.342-1.345); p<0.001 |
| 1 dose only | 266,661 (7.8%) | 348,899 (6.0%) |  |
| Two doses | 1,499,057 (44.0%) | 1,208,476 (20.7%) |  |
| Third dose | - | 2,558,354 (43.7%) |  |
| Vaccination status unknown | 241,163 (7.1%) | 355,951 (6.1%) |  |
| **Total SARS-CoV2 Hospitalizations (All age groups-Entire NIMS population)** |  |  |  |
| All persons | 31,839 | 34,593 |  |
| Unvaccinated | 11,767 (37.0%) | 8,996 (26.0%) | RR 0.70 (0.69-0.72); p<0.001 |
| All Vaccinated | 19,729 (62.0%) | 25,254 (73.0%) | RR 1.18 (1.16-1.19); p<0.001 |
| 1 dose only | 1,323 (4.2%) | 1,601 (4.6%) |  |
| Two doses | 18,406 (57.8%) | 6,514 (18.8%) |  |
| Third dose | - | 17,139 (49.5%) |  |
| Vaccination status unknown | 343 (1.1%) | 343 (1.0%) |  |
| **Total SARS-CoV2 Deaths (All age groups-Entire NIMS population)** |  |  |  |
| All persons | 12,506 | 11,604 |  |
| Unvaccinated | 2,578 (20.6%) | 1,771 (15.3%) | RR 0.74 (0.70-0.78); p<0.001 |
| All Vaccinated | 9,828 (78.6%) | 9,766 (84.2%) | RR 1.07 (1.06-1.08); p<0.001 |
| 1 dose only | 386 (3.1%) | 386 (3.3%) |  |
| Two doses | 9,442 (75.5%) | 2,929 (25.2%) |  |
| Third dose | - | 6,451 (55.6%) |  |
| Vaccination status unknown | 100 (0.8%) | 67 (0.6%) |  |
| **Case fatality rate (CFR): (All age groups-Entire NIMS population)** |  |  |  |
| All persons | 0.39% | 0.21% | RR 0.54 (0.52-0.55); p<0.001 |
| Unvaccinated | 0.18% | 0.13% | RR 0.70 (0.66-0.74); p<0.001 |
| All Vaccinated | 0.56% | 0.24% | RR 0.43 (0.41-0.44); p<0.001 |
| Two doses | 0.63% | 0.24% | RR 0.38 (0.37-0.40); p<0.001 |
| Third dose | - | 0.25% |  |
| **Risk of Hospitalizations (RH): (All age groups-Entire NIMS population)** |  |  |  |
| All persons | 0.99% | 0.62% | RR 0.63 (0.62-0.64); p<0.001 |
| Unvaccinated | 0.84% | 0.65% | RR 0.78 (0.76-0.80); p<0.001 |
| All Vaccinated | 1.12% | 0.61% | RR 0.55 (0.54-0.56); p<0.001 |
| Two doses | 1.23% | 0.54% | RR 0.44 (0.43-0.45); p<0.001 |
| Third dose | - | 0.67% |  |
| **Hospitalization death rate (HDR): (All age groups-Entire NIMS population)** |  |  |  |
| All persons | 39.4% | 33.7% | RR 0.86 (0.84-0.87); p<0.001 |
| Unvaccinated | 21.9% | 19.7% | RR 0.90 (0.85-0.95); P<0.001 |
| All Vaccinated | 49.8% | 38.7% | RR 0.78 (0.76-0.79); p<0.001 |
| Two doses | 51.3% | 45.0% | RR 0.88 (0.85-0.90); p<0.001 |
| Third dose | - | 37.6% |  |

**Tables 1b:**
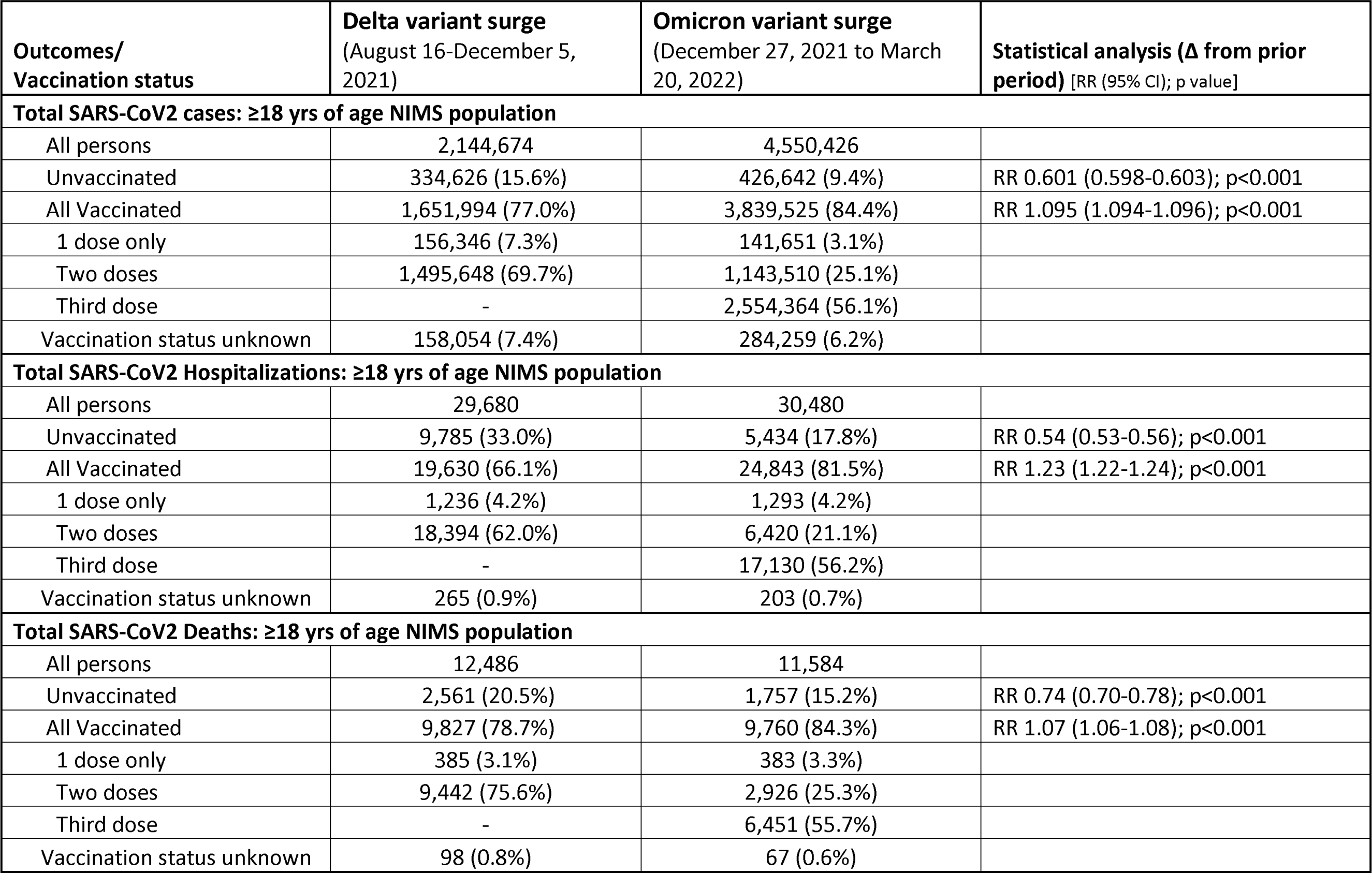
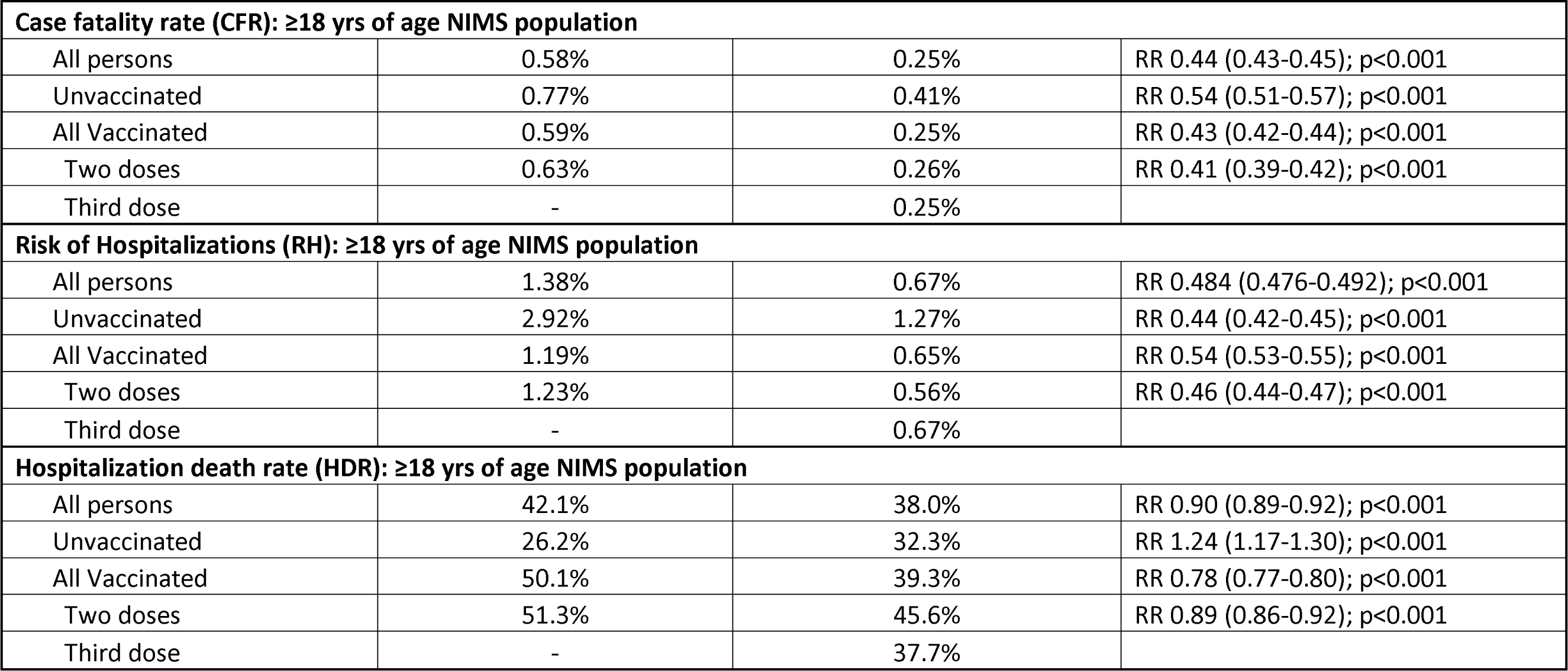
SARS-CoV2 cases, SARS-CoV2 hospitalizations, SARS-CoV2 deaths, the case fatality rate, risk of the hospitalizations and the hospitalization deaths rate among the over 18 years of age NIMS population during the Omicron variant surge compared with the Delta variant surge.

**Tables 1c:** SARS-CoV2 cases, SARS-CoV2 hospitalizations, SARS-CoV2 deaths, the case fatality rate, risk of the hospitalizations and the hospitalization deaths rate among the over 50 years of age NIMS population during the Omicron variant surge compared with the Delta variant surge.

| **Outcomes/**<br>**Vaccination status** | **Delta variant surge**<br>**(August 16-December 5, 2021)** | **Omicron variant surge**<br>**(December 27, 2021 to March**<br>**20, 2022)** | **Statistical analysis ($\Delta$ from prior**<br>**period) [RR (95% CI); p value]** |
| --- | --- | --- | --- |
| **SARS-CoV2 cases: $\geq 50$ yrs of age NIMS population** |  |  |  |
| All persons | 757,942 | 1,544,744 |  |
| Unvaccinated | 42,796 (5.6%) | 53,365 (3.5%) | RR 0.61 (0.60-0.62); p<0.001 |
| All Vaccinated | 669,847 (88.4%) | 1,410,754 (91.3%) | RR 1.033 (1.032-1.034); p<0.001 |
| 1 dose only | 12,949 (1.7%) | 15,175 (1.0%) |  |
| Two doses | 656,898 (86.7%) | 159,732 (10.3%) |  |
| Third dose | - | 1,235,847 (80.0%) |  |
| Vaccination status unknown | 45,299 (6.0%) | 80,625 (5.2%) |  |
| **SARS-CoV2 Hospitalizations: $\geq 50$ yrs of age NIMS population** |  |  |  |
| All persons | 21,300 | 22,456 |  |
| Unvaccinated | 4,859 (22.8%) | 2,924 (13.0%) | RR 0.57 (0.55-0.60); p<0.001 |
| All Vaccinated | 16,330 (76.7%) | 19,452 (86.6%) | RR 1.13 (1.12-1.14); p<0.001 |
| 1 dose only | 592 (2.8%) | 636 (2.8%) |  |
| Two doses | 15,738 (73.9%) | 3,862 (17.2%) |  |
| Third dose | - | 14,954 (66.6%) |  |
| Vaccination status unknown | 111 (0.5%) | 80 (0.4%) |  |
| **SARS-CoV2 Deaths: $\geq 50$ yrs of age NIMS population** |  |  |  |
| All persons | 11,912 | 11,184 |  |
| Unvaccinated | 2,216 (18.6%) | 1,595 (14.3%) | RR 0.77 (0.72-0.81); p<0.001 |
| All Vaccinated | 9,613 (80.7%) | 9,528 (85.2%) | RR 1.06 (1.04-1.07); p<0.001 |
| 1 dose only | 360 (3.0%) | 350 (3.1%) |  |
| Two doses | 9,253 (77.7%) | 2,795 (25.0%) |  |
| Third dose | - | 6,383 (57.1%) |  |
| Vaccination status unknown | 83 (0.7%) | 61 (0.5%) |  |
| **Case fatality rate (CFR): ≥50 yrs of age NIMS population** |  |  |  |
| All persons | 1.57% | 0.72% | RR 0.46 (0.45-0.47); p<0.001 |
| Unvaccinated | 5.18% | 2.99% | RR 0.58 (0.54-0.61); p<0.001 |
| All Vaccinated | 1.44% | 0.68% | RR 0.47 (0.46-0.48); p<0.001 |
| Two doses | 1.41% | 1.75% | RR 1.24 (1.19-1.30); p<0.001 |
| Third dose | - | 0.52% |  |
| **Risk of Hospitalizations (RH): ≥50 yrs of age NIMS population** |  |  |  |
| All persons | 2.81% | 1.45% | RR 0.52 (0.51-0.53); p<0.001 |
| Unvaccinated | 11.35% | 5.48% | RR 0.48 (0.46-0.50); p<0.001 |
| All Vaccinated | 2.44% | 1.38% | RR 0.57 (0.55-0.58); p<0.001 |
| Two doses | 2.40% | 2.42% | RR 1.01 (0.97-1.04); p=0.606 |
| Third dose | - | 1.21% |  |
| **Hospitalization death rate (HDR): ≥50 yrs of age NIMS population** |  |  |  |
| All persons | 55.9% | 49.8% | RR 0.89 (0.87-0.91) p<0.001 |
| Unvaccinated | 45.6% | 54.5% | RR 1.20 (1.14-1.25); p<0.001 |
| All Vaccinated | 58.9% | 49.0% | RR 0.83 (0.82-0.85); p<0.001 |
| Two doses | 58.8% | 72.4% | RR 1.23 (1.20-1.26); p<0.001 |
| Third dose | - | 42.7% |  |

The detailed analysis of ≥50 years of age vaccinated population and analysis of changing patterns of cases, hospitalizations, and deaths among the various age groups are tabulated in the supplemental appendix. We also performed a comparative analysis of the two vaccination databases (NIMS and population vaccinated in England from the UK coronal virus dashboard); the details appear in the supplemental appendix. For the entire population of England, there was 8.4-9.0% higher proportion of the unvaccinated population in NIMS database between August 16, 2021 to March 27, 2021, which can underestimate the vaccine effectiveness by about 7.5 to 12.5% (less variability in ∼4.0% range when the VE was good for two doses in February-June 2021 period). This disparity did not alter SARS-CoV2 cases per 100,000 population among the vaccinated groups (two doses, third dose, and one dose).

## DISCUSSION

There was a total of 22.07 million SARS-CoV2 cases, 848,911 hospitalizations, and 175,070 deaths reported in the UK during the pandemic as of May 1, 2022. The highest percentage (51.3%) of the cases, 28.8% of total hospitalizations, and 16.4% of total deaths occurred during the Omicron variant surge in 21 weeks until May 1, 2022. The Omicron variant surge was associated with a lower case fatality rate and lower risk of hospitalizations than the Delta variant surge during the first twelve weeks until February 27, 2022 in our study; a similar finding was reported in a recent study for the period ending January 9, 2022^29^. We observed significantly increased case fatality rates and risk of hospitalizations during the latter part of the Omicron variant surge, which was higher than what was observed during the Delta variant surge. We also noted an increased proportion of cases among ≥50 years age groups associated with a significantly increased proportion of hospitalizations and deaths particularly in the ≥75 years age group (80.1% of total deaths) after February 27, 2022. A similar pattern of a significantly increased proportion of SARS-CoV2 cases and hospitalizations in ≥50 years old population was associated with a significantly increased proportion of deaths among ≥ 70 years old age group during the latter part of the Delta variant surge in the UK^30^.

The ≥18 years of age NIMS population in our study showed a waning of vaccine effectiveness for the two doses during the Delta variant surge (since September, 2021) with negative vaccines effectiveness for the infections (higher proportion of infection among the vaccinated population than unvaccinated) during the Omicron variant surge in all the vaccinated groups including the third dose population since December 20, 2021. The two-proportions test showed a significantly higher proportion of infections in vaccinated with two doses (including all vaccinated) than the unvaccinated during the Delta variant surge (since September 2021). Additionally, the vaccinated with the third dose had significantly higher infection rates versus the vaccinated with two doses and unvaccinated during the Omicron variant surge (since February 2022). This was associated with a significantly higher proportion of hospitalizations in vaccinated with two doses versus the unvaccinated during the Omicron variant surge (since January 2022), a higher proportion of deaths in vaccinated with two doses versus the unvaccinated (since October 2021), and a higher proportion of hospitalizations and deaths in vaccinated with the third dose versus the vaccinated with two doses and the unvaccinated during the Omicron variant surge (since February 2022). In our study, the unvaccinated have a higher case fatality rate and risk of hospitalizations than the vaccinated during the Delta variant surge. However, both the vaccinated and unvaccinated populations showed a significant decline in the case fatality rate and risk of hospitalization during the Omicron variant surge when compared to the Delta variant surge among all ages, and over 18 years of age. Additionally, a significantly decreased SARS-CoV2 cases in the unvaccinated population during the Omicron variant surge (compared to the Delta variant surge) was associated with significantly decreased hospitalizations and deaths among over 18 years and 50 years age groups. Similarly, significantly increased SARS-CoV2 infection in the vaccinated population of over 18 and 50 years of age was associated with significantly increased hospitalization and deaths during the same period. The improved outcomes among the unvaccinated observed in our study are similar to the favorable health outcomes that were observed among the self-selected unvaccinated cohort^31^.

The NIMS database also clearly shows the disparities in the vaccination rates with British white and least deprived per IMD decile score (vaccinated with highest percentages of the third dose) have the lowest infection rates, hospitalizations and deaths due to COVID-19 even before the vaccination. The ethnic minorities and the most deprived based on IMD decile score (have lowest rates of third booster vaccinations, highest rates receiving only two doses without the booster and highest rates of unvaccinated compared to British white) also had higher infection rates, hospitalizations, and deaths due to COVID-19 from the beginning of the pandemic. Our study of changing case fatality rate among ≥50 years of age group vaccinated population showed heterogenicity. The all-vaccinated population of ≥50 years of age showed a significant decline in the case fatality rate during the Omicron variant surge compared to the Omicron variant surge. However, those vaccinated with two doses only (higher proportion of minorities and most deprived based on IMD) have a significantly higher case fatality rate during the Omicron variant surge versus the vaccinated with two doses during the Delta variant surge (majority of them are white and/or least deprived).

In our data analysis, the pre-existing conditions were associated with 95.8 to 96.8% of deaths among 60+ years old population in England during the Omicron variant surge until May 1, 2022. The pre-existing conditions were also reported in the majority of COVID-19 deaths during the prior surges including the Delta variant surge^32^. The racial disparities with increased deaths due to COVID-19 among ethnic minorities compared to white British population were reported in multiple UK Office for National Statistics publications from the beginning of the COVID-19 pandemic in 2020 until recent publication on April 7,2022^33–36^. Other studies have also shown that increased infections, hospitalizations, and deaths were noted among racial minorities in UK^37, 38^.

Based on the analysis, the vaccination groups (third dose, two doses only, and unvaccinated) have heterogenous populations that have different known infection rates, hospitalizations, and deaths due to COVID-19 dating back to the beginning of the pandemic way before the UK started the vaccination program in December 2020. In light of these known variables, SARS-CoV2 cases among the compared groups for vaccine effectiveness can be adversely affected by the behavior of the population and/ or opportunity for exposure either at home or at work based on the density of population^16, 39–41^. Additionally, all the scientific evidence suggests that in addition to increased cases, a vast majority of hospitalizations and deaths were associated with pre-existing conditions, especially in the elderly population^32, 42–44^. Our study demonstrates that changes in the proportion of cases among various age groups during surges were associated with similar changes in hospitalizations (England) and deaths (England and Wales), particularly in the elderly. We also noted that significantly increased SARS-CoV2 cases in the vaccinated population were associated with significantly increased hospitalizations and deaths; whereas significantly decreased cases among the unvaccinated population were associated with significantly decreased hospitalizations and deaths. Based on these known variables, the vaccine effectiveness for hospitalizations and deaths needs to be adjusted for increased cases due to behavior or exposure risk and other known variables including racial ethnicity and deprivation score. Additionally, VE must be adjusted for pre-existing conditions, especially in the elderly (97.95% of the deaths during the latter part of the Omicron variant surge occurred among ≥50 years of age and 80.07% of deaths ≥75 years of age).

The disparities among SARS-CoV2 hospitalizations and deaths among the vaccinated groups must be interpreted with great caution because the randomized controlled trials for the COVID-19 vaccines did not show demonstrable benefits in preventing hospitalization and/ or deaths and were not adequately powered to study these outcomes^45–49^. A recent randomized controlled study using the third dose of BNT162b2 vaccine did not report any benefit in preventing severe illness, hospitalizations or deaths^50^. Additionally, the six months data of the mRNA-1273 and BNT162b2 vaccines showed similar levels of vaccine efficacy for preventing symptomatic infections and severe illness, and it did not show any discordance with good vaccine efficacy for severe illness and suboptimal vaccine efficacy for preventing symptomatic infections^48, 51^. The observational studies reported good vaccine effectiveness for hospitalizations, severe illness and/or deaths by reported adjustment of the variables in their statistical models for the population based on the vaccination status, but they did not publish the detailed baseline characteristics (pre-existing conditions, ethnic disparities, deprivation score and their vaccination disparities) among those who were hospitalized or died due to SARS-CoV2 infection based on the vaccination status^17, 18, 52, 53^. A large US study (Dec 14, 2020-Aug 8, 2021) analyzed the electronic healthcare records of individuals in their health care system (n=3,436,957 of ≥12 years of age) reported 184,041 SARS-CoV2 infections and 12,130 SARS-CoV2 hospitalizations^43^. They reported significantly higher comorbidities among those with SARS-CoV2 hospitalizations; the uninfected and infected total populations have similar comorbidities ^43^. The study did not publish the comorbidities and vaccination disparities among those who were hospitalized due to COVID-19 based on vaccination status. The study, reported vaccine effectiveness for SARS-CoV2 infection and hospitalizations and did not report if there was any mortality benefit among the vaccinated population^43^. In the absence of vaccine effectiveness to prevent COVID-19 infections, it is of utmost importance to develop validated models with a separate independent, and comprehensive analysis of all factors (age, gender, ethnic/IMD score, and vaccination disparities and most importantly a comprehensive analysis of all pre-existing conditions) that can independently alter the outcomes (SARS-CoV2 hospitalizations and deaths).

This analysis should be independent of the risk factors in the population. After an independent analysis of these variables, a determination can be made if any intervention (vaccination or any drug therapies) was beneficial in reducing the risk of hospitalizations and/ or deaths due to SARS-COV2 infections. Difficulties in conducting clinical trials in the critical care settings during the COVID-19 pandemic were already outlined with higher chances for the occurrence of type I and type II errors due to pitfalls in appropriately matching the compared groups for all the variables that can independently affect the studies outcomes without bias^54–56^. Any errors in the analysis or interpretation of data in the current situation (lack of effectiveness to prevent infections) may overestimate the effectiveness (for hospitalizations and deaths) that can create a false sense of security and may harm the vaccinated population if they do not take adequate precautions to avoid infections. This is more relevant in the view of studies showing decreased vaccine effectiveness among the elderly population that are at the highest risk for increased hospitalizations and deaths^18, 57^. We propose urgent scrutiny while evaluating vaccine effectiveness for hospitalizations and deaths in order to maintain the scientific integrity of data and to promote public safety. Any SARS-CoV2 hospitalization and death has to be analyzed by considering all of the underlying individual risk factors. This is an absolute necessity in the setting of improved outcomes that we observed with a significantly decreased case fatality rate and risk of hospitalizations among both the unvaccinated and vaccinated populations during the Omicron variant surge. We also observed a disparity between the vaccinated and unvaccinated populations. While the unvaccinated population had a significantly decreased number of cases, hospitalizations, and deaths, the vaccinated population observed a significant increase in these three areas during the Omicron variant surge.

The decreased cases among the unvaccinated population during the Omicron variant surge is probably due to protection from their prior natural infections that were already shown to give lasting immunity, which needs further study^58–60^. The different vaccine effectiveness methodologies used by the Israeli (traditional vaccine effectiveness) and UK (test-negative case-control design) health officials are probably the main reasons for the discordance between the Israeli data that led to the early approvals of the third dose on July 30, 2021 (Delta variant surge) and fourth dose on December 21, 2021 (Omicron variant surge) while the UK data is still showing favorable vaccine effectiveness using test-negative case-control design especially for hospitalizations and deaths which needs to be further studied^14, 17, 18, 61, 62^.

One limitation of our study is that it is a retrospective observational study of publicly reported data, and the generalizability of the findings is limited to the population studied in the UK. We used NIMS data population as the denominator for comparing outcomes (SARS-CoV2 cases, hospitalizations, and deaths per 100,000 population) among various age groups based on vaccination status which was also published by UKHSA in their regular weekly vaccine surveillance reports from August 16, 2021 to March 27, 2022. Our use of the unvaccinated population from the NIMS databases for the vaccine effectiveness may have overestimated the unvaccinated population by ∼8.4% to 9.0% and may have underestimated the vaccine effectiveness by about 7.5% to 12.5% which is not a major issue during the Omicron variant surge with waned vaccine effectiveness as outlined in the supplemental appendix. Our findings of the suboptimal of vaccine effectiveness to prevent infections has validity with a recent UKHSA publication reported protection against the *mild disease only* with the third dose vaccination during the initial stages of the Omicron variant surge and the actions of the UK authorities to recommend the fourth dose later during the Omicron surge ^12, 13, 28^. The prior UKHSA publications reported vaccine effectiveness for the SARS-CoV2 hospitalizations and deaths during the Delta variant surge and the recent UKHSA third dose study during the Omicron variant surge did not report outcomes of 52,369 SARS-CoV2 hospitalizations and 4,101 COVD-19 deaths among over 18 years of age that occurred in England during the study period^18, 24, 28, 29, 63^. The various time periods that were used in our statistical analysis are based on the timing of the start of a surge and/or changing patterns of an increased proportion of cases, hospitalizations or deaths during the surge that were observed on the plotted data. The retrospective analysis of data based on the changing patterns of outcomes yields valuable epidemiological information that can be used for public safety during the emerging pandemic as we have shown in our data analysis. Another limitation of our analysis is that we are unable to adjust the data for behavior among the compared groups which can affect the chances of contracting the disease. The changing of community testing standards in the UK since April 1, 2022 may have underestimated SARS-CoV2 cases and can overestimate CFR and RH reported in our study for that four weeks period. However, this does not invalidate the increased proportion of SARS-CoV2 cases noted among the elderly population in our study during the latter part of the Omicron variant and Delta variant surges that were associated with a significantly increased proportion of hospitalizations and deaths – a matter of significant health importance^64^. Our analysis identified multiple variables including ethnic disparities, deprivation score disparities and pre-existing conditions among the vaccinated populations that can affect the outcomes (SARS-CoV2 cases, hospitalizations, ICU/HDU admissions and deaths). We are unable to adjust the data specifically for the hospitalizations and deaths since we do not have access to the data (among those who were hospitalized or died) and there are no validated models that can yield unbiased data comparable to the randomized controlled trials. Furthermore, based on the vaccination disparities (ethnic and IMD score) the unadjusted data of the third dose population is expected to have better outcomes, and unadjusted data of unvaccinated and two doses without the third dose population are expected to have the worse outcomes (SARS-CoV2 hospitalizations, ICU/HDU admissions and deaths).

However, our findings even with the unadjusted data have public health importance as there was an increased proportion of SARS-CoV2 cases among the vaccinated population (including the third dose) that was associated with a significantly increased proportion of SARS-CoV2 hospitalizations and deaths. On the other hand, the decreasing proportion of SARS-CoV2 cases during the Omicron variant surge in the unvaccinated population is associated with a decreased proportion of SARS-CoV2 hospitalizations and deaths.

The strength of our investigation is the comprehensive analysis of nationwide real-world data of the various variables associated with SARS-CoV2 infections, hospitalizations, and deaths in the UK during the entire pandemic. Our analysis also identified heterogenicity in the vaccinated (two doses or third dose) and unvaccinated populations that had variabilities in infection rates, hospitalizations, and death rates from the beginning of the pandemic. This factor needs to be adjusted while calculating the vaccine effectiveness. The findings in this study are similar to a recent study in Israel that showed increasing breakthrough infections with a waning of vaccine effectiveness in the elderly population who are considered immune (mostly third dose booster population). These breakthrough infections were associated with increased severe illness and deaths during the Omicron variant surge^40^. We used the ≥18 years age group for evaluating the vaccine effectiveness and outcomes among the vaccinated populations, which is the similar age group included in the randomized controlled trails for vaccine efficacy^46, 47, 49^. A recent UKHSA publication also used the same group for vaccine effectiveness during the Omicron variant surge^28^. We have also included a comprehensive data on over 50 years of age population in the supplemental appendix.

## CONCLUSIONS

There was no discernable optimal vaccine effectiveness to prevent infections among the third dose ≥18 years of age population since December 20, 2021 during the initial part of the Omicron variant surge in the UK. The increased SARS CoV2 cases in the vaccinated population (including the third dose) among those over 18 years of age during the Omicron variant surge were associated with a significant proportion of hospitalizations and deaths; whereas the decreasing cases in the unvaccinated population were associated with a decreased proportion of hospitalizations and deaths. In our study, we also noted favorable outcomes in both unvaccinated and vaccinated populations with significantly decreased case fatality rate and risk of hospitalizations during the Omicron variant surge. Our data analysis identified heterogenous vaccinated (third dose, received two doses without booster) and unvaccinated populations with well-known variables that can increase the risk for hospitalizations and deaths based on ethnicity, deprivation score, and pre-existing conditions. The vaccine effectiveness for hospitalizations and deaths should be adjusted for these variables by developing validated models to avoid bias. This underscores the importance of the public health measures directed at uniform screening protocols, and protective measures to prevent infections especially in the elderly population irrespective of the vaccination status.

## Supporting information

Suplementary appendix Emani VR at al

## Data Availability

All data produced in the present study are available upon reasonable request to the authors

https://coronavirus.data.gov.uk/

https://www.gov.uk/government/statistics/national-flu-and-covid-19-surveillance-reports-2021-to-2022-season

https://www.gov.uk/government/publications/covid-19-vaccine-weekly-surveillance-reports

https://www.gov.uk/government/statistics/national-flu-and-covid-19-surveillance-reports

https://www.england.nhs.uk/statistics/statistical-work-areas/covid-19-hospital-activity/

https://www.ons.gov.uk/peoplepopulationandcommunity/birthsdeathsandmarriages/deaths/datasets/weeklyprovisionalfiguresondeathsregisteredinenglandandwales

https://www.england.nhs.uk/statistics/statistical-work-areas/covid-19-daily-deaths/weekly-total-archive/

https://www.gov.uk/government/publications/investigation-of-novel-sars-cov-2-variant-variant-of-concern-20201201

https://www.ons.gov.uk/peoplepopulationandcommunity/birthsdeathsandmarriages/deaths/articles/coronaviruscovid19relateddeathsbyethnicgroupenglandandwales/2march2020to15may2020

https://www.ons.gov.uk/peoplepopulationandcommunity/birthsdeathsandmarriages/deaths/articles/updatingethniccontrastsindeathsinvolvingthecoronaviruscovid19englandandwales/24january2020to31march2021

