## Supplementary material for "Increasing SARS-CoV2 cases, hospitalizations, and deaths among the vaccinated populations during the Omicron (B.1.1.529) variant surge in UK": Suplementary appendix Emani VR at al

#### Supplementary appendix

#### METHODS:

The following are the additional details to methods that are described in in the manuscript.

**Missing data:** The UKHSA regularly publishing the SARS-CoV2 outcomes as an aggregate total for prior four weeks period (SARS-CoV2 cases, hospitalizations and deaths) among various vaccinated age groups weekly from August 16, 2021 to March 27, 2022. The UKHSA did not publish the outcomes for the SARS-CoV2 cases, hospital admissions and deaths among vaccinated population for the four weeks period; from November 23 to December 20, 2021 (week 48-51) and the hospital admissions among vaccinated groups for the four weeks for the period January 31, 2022 to February 27, 2022 (week 5-8). However, the aggregate four weeks outcomes for the prior and later weeks were available, thereby limiting the impact of missing data only to those missing week(s). The UKHSA stopped reporting the data of SARS-CoV2 cases, hospitalizations and deaths among NIMS population in the weekly vaccine surveillance reports. We are therefore unable to report vaccine effectiveness or the proportionality test results from March 28-May 1, 2022 period.

**Pillar 1 and 2 cases among various age groups in England for weeks 13-17 in 2022 (March 28-May 1, 2022):** The National Flue and COVID-19 surveillance reports were published rolling two weekly pillar 1 (public health laboratories and hospitals) and pillar 2 (community testing) SARS-CoV2 cases among age groups from September 28, 2020 to March 27, 2022. Since March 28, 2022 only weekly pillar 1 and 2 SARS-CoV2 cases per 100,000 were published in the weekly National Flue and COVID-19 surveillance reports. We computed weekly SARS-CoV2 cases (pillar 1 and 2) among various age groups in England for the period March 28-May 1, 2022 (week 13-17) using mid-year 2020 census estimate of England (Table S4) and case rates (pillar 1 and 2) per 100,000 population for each age group.

**Validation of the impact of unvaccinated population in England for vaccine effectiveness:**

We used the regularly published data (SARS-CoV2 cases, hospitalizations, and deaths among the age groups based on the vaccination status) by the UK Health Security Agency (UKHSA) in their weekly UK vaccine surveillance reports (in between August 16, 2021 to March 27, 2022) derived from the population linked to the National Immunization Management Service (NIMS)[1]. We used the National Immunization Management Service (NIMS) databases population denominator that was published weekly in the National Flue and COVID-19 surveillance reports to calculate the vaccination rates and to calculate the rates of SARS-CoV2 infections, hospitalizations and deaths among the age groups based on the vaccination status[2]. The NIMS data was also used by UKHSA to publish the rates of SARS-CoV2 infections, hospitalizations and deaths based on the vaccination status[1]. Of the two potential sources of population denominators, the NIMS can potentially overestimate the unvaccinated population in some age groups and the Office for National Statistics (ONS) denominator can underestimate the unvaccinated populations in some age groups[1].

We used the England vaccinated population database of all ages (age specific denominators other than over 12 years of age are not published) from the UK corona virus dashboard and compared it with the NIMS database population of all ages[2,3]. We calculated the denominator for vaccination groups (1 dose only, 2 doses only and third dose) based on the raw data for each week. We used the mid 2020 England census estimated population to derive the unvaccinated population for England vaccination database (UK corona virus dashboard)[4]. We used the NIMS total population to derive unvaccinated population for the NIMS database[2].

We used the weekly published total SARS-CoV2 cases (all ages) based on the vaccination status that was published by UKHSA between August 16, 2021 to March 27, 2022 in the vaccine surveillance reports and the Delta variant verified cases in England published by the Public Health England in between February 1 – September 12, 2021 to validate the two databases[1,5]. The incidence rate per 100,000 population among the vaccination groups and the vaccine effectiveness was calculated using both databases' denominators for the comparative analysis.

#### RESULTS:

**SARS-CoV2 cases, SARS-CoV2 Hospitalizations (in England) and SARS-CoV2 deaths (England and Wales) among age groups** (Figures 2a-c; Tables S1a-c)**:**

Analysis of SARS-CoV2 cases (Figure 2a and Table S1a) in England among various age group showed significantly increased percent of SARS-CoV2 cases among <30 years (0-19 years, 20-29 years) during the transition from the Alpha variant surge to the initial part of the Delta variant surge. During the latter part of the Delta variant surge, the over 50 years of age (particularly over 70 years of age) have significantly increased cases with decreased cases among 20-29 years of age.  The analysis of various age groups SARS-CoV2 hospitalizations (Figure 2b and Table S1b) showed significantly increased hospitalizations among under 55 years of age (particularly among 0-34 years of age) during the initial part of Delta variant surge. During the latter part of Delta variant surge, hospitalizations among over 55 years of age (particularly over 65 years) significantly increased with significantly decreased hospitalization among the other age groups. The same was observed during the latter part of Omicron variant surge (February 28-May 1, 2022) for the  ≥75 years of age group. Analysis of deaths among various age groups in England and Wales (Figure 2c and Table S1c) showed a significantly increased deaths among under 70 years of age (particularly among under 50 years) during the Delta variant surge until August 1, 2021,significantly increased deaths noted among over 70 years of age, and significantly decreased deaths among < 70 years of age during the latter part of Delta variant surge (August 2-December 5, 2021).  Similarly, during the latter part of the Omicron variant surge (February 28-May 1, 2022), significantly increased deaths were noted among over 75 years of age and significantly decreased deaths among under 75 years of age. The significantly increased SARS-CoV2 cases during the latter part of the Alpha variant surge and the initial part of Delta variant surge among <50 years of age groups were associated with significantly increased deaths during the same period.

**Vaccine effectiveness among over 50 years NIMS population in UK** (Table S7b-c and Figure 4)

The Public Health England reported the outcomes among the Delta variant cases (confirmed and provisional) in the UK technical briefings until September 12, 2022. The UK vaccine surveillance reports started publishing from August 16, 2021 until March 27, 2022. As shown in Table S3, about 66.7% of the over 50 years of age NIMS population was vaccinated with two doses of primary vaccination by May 23, 2021 during the beginning of the Delta variant surge. The vaccine effectiveness for ≥50 years of age NIMS population vaccinated with two doses was 53.1% (95%CI 48.4%-57.2%) for the period ending in June 20, 2021. Since June 21, 2021 period as shown in Table S7b, the vaccine effectiveness declined to 18.0% - 23.2% among ≥50 years of age vaccinated with two doses and was in the negative territory with incidence rate (IR) of cases higher among the vaccinated with two doses versus the unvaccinated since August 16, 2021 period. The vaccine effectiveness of the ≥50 years of age vaccinated with two doses was also in the negative territory since August 16, 2021 among the four weekly running cases that were reported in the weekly vaccine surveillance reports. In addition, the IR of cases among those vaccinated with two doses was higher than the unvaccinated. Since the rolling four weekly cases from December 20, 2021 to June 16, 2021, the vaccinated with the third dose among ≥50 years of age was also in the negative. The two-proportions test with continuity check (henceforth proportions test) also showed that the proportion of SARS-CoV2 cases were higher among the vaccinated population versus the unvaccinated since August 16, 2021 among the Delta variant cases (vaccinated with two doses; $X^2$= 2.9683, df = 1, p = 0.042 and all vaccinated $X^2$= 4.4235, df = 1, p-value = 0.01772). We observed the same with the NIMS population of ≥50 years of age (two doses cases; $X^2$= 708.94, df = 1, p<0.001; all vaccinated cases; $X^2$= 688.86, df = 1, p<0.001). Since the reporting period of December 20, 2021, those over 50 years of age vaccinated with third dose have significantly higher proportion of cases ($X^2$= 7987, df = 1, p<0.001) than the unvaccinated and since Jan 31, 2022, the third dose population has a significantly higher proportion of cases than the two doses population ($X^2$= 309.3, df = 1, p<0.001). The proportions test on the hospitalization among ≥50 years of age showed that vaccinated with two doses population ($X^2$= 678.34, df = 1, p<0.001) has a significantly higher proportion of hospitalizations than the unvaccinated since December 6. The third dose population of ≥50 years of age ($X^2$ = 5.523, df = 1, p = 0.009384) and all vaccinated population ($X^2$ = 10.468, df = 1, p<0.001) have a significantly higher proportion of hospitalizations than the unvaccinated since February 28, 2022. Similarly, the proportions test on the SARS-CoV2 deaths within 28 days of the positive test among those vaccinated with two doses were higher than the unvaccinated ($X^2$ = 39.061, df = 1, p<0.001) since November 8, 2021. On the contrary, the third dose population does not have a significantly higher proportion of deaths versus the unvaccinated as of March 27, 2022. Using the available data from vaccine surveillance reports, we compared the case fatality rate (CFR) and risk of hospitalizations(RH) during the Omicron variant surge (December 27, 2021-March 20, 2022) with the Delta variant surge (August 16-December 5, 2021). Our comparative analysis shows a significant decline in case fatality rate (all ages [0.21% vs 0.39%; RR 0.54 (0.52-0.55); p<0.001], over 18 years of age [0.25% vs 0.58%; RR 0.44 (0.43-0.45); p<0.001], and over 50 years of age [0.72% vs 1.57%; RR 0.46 (0.45-0.47); P<0.001]) and the risk of hospitalizations (all ages [0.62% vs 0.99%; RR 0.63 (0.62-0.64); p<0.001], over 18 years of age [0.67% vs 1.38%; RR 0.484 (0.476-0.492); p<0.001], and over 50 years of age [1.45% vs 2.81%; RR 0.52 (0.51-0.53); p<0.001]). There was a significantly decreased CFR among all persons (0.72% vs 1.57%; RR 0.46 (0.45-0.47); P<0.001), unvaccinated (2.99% vs 5.18%; RR 0.58 (0.54-0.61); p<0.001), and all vaccinated (0.68 vs 1.44%; RR 0.47 (0.46-0.48); p<0.001) populations of over 50 years of age during the Omicron variant surge (December 27, 2021 to March 20, 2022) compared to the Delta variant surge (August 16-December 5, 2021). Similarly, the RH was significantly decreased among all persons (1.45% vs 2.81%; RR 0.52 (0.51-0.53); p<0.001), unvaccinated (5.48% vs 11.35%; RR 0.48 (0.46-0.50); p<0.001), all-vaccinated (1.38% vs 2.44%; RR 0.57 (0.55-0.58); p<0.001) populations of over 50 years of age during the Omicron variant surge compared to Delta variant surge. Among those over 50 years of age vaccinated with two doses, the CFR significantly increased (1.75% vs 1.41%; RR 1.24 (1.19-1.30); p<0.001) with no significant change in the RH (2.42% vs 2.40%; RR 1.01 (0.97-1.04); p=0606) during the Omicron variant surge compared to the Delta variant surge. As shown in Table 1c, the significantly decreased SARS-CoV2 cases in unvaccinated [53,365 of 1,544,744 (3.5%) vs 42,796 of 757,942 (5.6%); RR 0.61 (0.60-0.62); p<0.001] during the Omicron variant surge (December 27, 2021 to March 20, 2022) among over 50 years of age was associated with significantly decreased hospitalizations [2,924 of 22,456 (13.0%) vs 4,859 of 21,300 (22.8%); RR 0.57 (0.55-0.60); p<0.001] and significantly decreased SARS-CoV2 deaths [1,595 of 11,184 (14.3% vs 2,216 of 11,912 (18.6%); RR 0.77 (0.72-0.81); p<0.001] versus the Delta variant surge (August 16-December 5, 2021). Similarly, a significantly increased SARS-CoV2 cases among the all vaccinated population [1,410,754 of 1,544,744 (91.3%) vs 669,847 of 757,942 (88.4%); RR 1.033 (1.032-1.034); p<0.001] during the Omicron variant surge was associated with significantly increased SARS-CoV2 hospitalizations [19,452 of 22,456 (86.6%) vs. 16,330 of 21,300 (76.7%); RR 1.13 (1.12-1.14); p<0.001] and significantly increased SARS-CoV2 deaths [9,528 of 11,184 (85.2%) vs. 9,613 of 11,912 (80.7%); RR 1.06 (1.04-1.07); p<0.001].

Analysis of the SARS-CoV2 deaths in England and Wales showed that 97.95% and 80.07% of the deaths during the latter part of the Omicron variant surge occurred among ≥50 years and ≥75 years of age groups, respectively. The pre-existing conditions were present in 95.6% of deaths among all ages and 96.2% of deaths ≥60 years of age during the Omicron variant surge in England. The deaths among the elderly vaccinated population have to be considered in the context of their advanced age along with the pre-existing conditions.

**Vaccine effectiveness of the third dose population prior to December 20, 2021:**

Since the third dose vaccination approval in September 2021, about 77.3% of the ≥50 years of age NIMS population were vaccinated with the third dose by December 19,

2021. The UKHSA did not report the third dose outcomes until December 20, 2021 in their weekly reports. In our study, we analyzed the 999,064 pillar 2 cases among over 18 years old reported from a UKHSA publication with 336,046 cases among the third dose (booster), 484,742 among the two doses, and 137,478 cases among the unvaccinated[6]. Using the third dose weekly vaccinated populations between December 26, 2021, January 2, 2022, and January 9,2022, the best vaccine effectiveness for the third dose was 13.8%, 18.4%, and 22.2%, respectively. During the same period (November 27, 2021-January 12, 2022) England reported 52,369 SARS-CoV2 hospitalizations and 4,101 COVD-19 deaths among over 18 years of age[7,8].

**Comparative analysis to study the impact of unvaccinated population on vaccine effectiveness** (NIMS database denominator versus England vaccination database denominator data from the UK corona virus dashboard)**:**

**The percentage of unvaccinated disparities among the two databases**

The comparison of the percent of the unvaccinated population in the NIMS database (Table S3a) and the England vaccination database data from the UK corona virus dashboard (Table S3c) show that the NIMS unvaccinated population is 8.4% (as of August 15, 2021) to 9.0% (as of March 27, 2022) higher than the England vaccination database.

**The SARS-CoV2 cases per 100,000 population among the vaccinated groups**

We studied the SARS-CoV2 cases incidence rate per 100,000 population among the confirmed Delta variant cases of all ages in England (February 1 – September 12, 2021) and SARS-CoV2 cases all ages from NIMS linked population published in the COVID-19 vaccine surveillance reports (August 16, 2021 – March 27, 2022) as shown in Table S7d. The incidence rate of the SARS-CoV2 cases per 100,000 did not vary much among the vaccinated population (1 dose, two doses only, or third dose) as shown in Table S7d. However, the incidence rate of SARS-CoV2 cases per 100,000 unvaccinated population was underestimated using the NIMS database as the denominator. The same was overestimated using the England vaccinated database as the denominator.

**Impact on vaccine effectiveness using comparative analysis of each of the two databases population as denominator**

*Vaccine effectiveness (VE) two doses among confirmed Delta variant cases in England by Public Health England (Table S7e):* The vaccine effectiveness for those vaccinated with two doses during Feb 1-June 20, 2021 was 84.4% (95% CI 83.9% to 84.8%) using the NIMS database and 88.8% (95% CI 88.0% to 88.7%) using the England database as the denominators with a difference of 4.4% of underestimation of VE with the NIMS database. During the subsequent four weeks period ending in July 18, 2021, the VE of those vaccinated with two doses was 70.3% (using NIMS database as the denominator) and 78.8% (England DB as the denominator) with a difference of 8.5%. During the subsequent two weeks period ending in August 1, 2021, the VE for those with two doses were 53.4% and 67.0%, respectively with a difference of 13.6%. During the subsequent six weeks, there was a decline in VE among those vaccinated with two doses using both the databases; for the two weeks period ending in September 12, 2021 the VE for those vaccinated with two doses was 27.8% (95% CI 26.1% to 29.3%) using the NIMS database as the denominator and 50.0% (95% CI 48.9% to 51.1%) using the England vaccination database as the denominator (NIMS underestimates VE versus the England database up to 22.3%).

*Vaccine effectiveness (VE) two doses and third dose among NIMS linked SARS-CoV2 cases reported in UKHSA vaccine surveillance reports (Table S7e):* Since the first reporting period for those vaccinated with two doses starting August 16 – Sept 12, 2021 four weeks period the VE was 31.4% (95% CI 30.9% to 32.0%) and 52.5% (95% CI 52.2% to 53.0%), respectively using the NIMS and England DB as the denominators with a 21.1% difference; since then the variation is steadily increasing. Since November 22, 2021 the VE values among those vaccinated with two doses using both the databases are in the negative. The third dose reporting period started from December

20, 2021 to January 16, 2022 four-week period. The third dose VE values (all ages) were 12.9% (95% CI 12.4% to 13.3%) and 44.0% (95% CI 43.7% to 44.3%), respectively using the NIMS and England DB as the denominators with a difference of 31.1%. Since then, there was a decline in the VE values in both the databases and by Feb 7- Mar 6, 2022, the VE for the third dose using both the database denominators was in the negative with a difference between the two databases as high as 86.9%.

In summary, when the VE for those vaccinated with two doses of the entire population of England (all ages) during the Feb 1-August 1, 2021 was good *when* the VE using both databases (NIMS vs England vaccination DB) was above the FDA recommended VE of >50%. The variation in VE among these databases was 4.0% to 13.6% with a lowest variation of 4.0% when the VE in both databases was greater than 80% and 13.6% variation when the VE was in the 53.4% to 67.0% range. The third dose VE since the December 20, 2021 was <50% in both databases (12.9% using NIMS as denominator and 44.0% using England DB as denominator), with the variation of 31.1%. The variation increased to 86.9% by March 27, 2022 and the VE for the third dose using both the databases as denominators was in negative since February 7, 2022.

**Public health restrictions, lockdowns and reopening in the United Kingdom (UK):** The UK instituted the first lockdown during the beginning of the pandemic on March 23, 2020. The second lockdown (October 30, 2020) and the third lockdown (December 22, 2020) were instituted during the second wave and the surge in cases during the Alpha variant surge.

The phased easing of restriction in UK started with reopening of the schools and colleges effective March 8, 2021 (week 10). The step 2 of the reopening became effective since April 12, 2021 and step 3 of the reopening since May 17, 2021 and by May 2021 the outdoor social contact were to be removed, two households or six people were allowed to meet indoors, and indoor hospitality services were provided and hotels were opened[9–12]. The UK government announced that most restrictions including face masks mandates, social distancing measures and capacity limitations in venues were lifted in England by July 19, 2021 and these were replaced by recommendations[11,12].

#### DISCUSSION

Based on the UKHSA reliance on the NIMS database linked outcomes (weekly reported number of SARS-CoV2 cases, hospitalizations, and deaths among various age groups based on the vaccination status and these outcomes per 100, 000 population based on the vaccination status) in the regular weekly vaccine surveillance reports, we also believe the NIMS database population is a better database that can be used as the denominator for an effective outcomes assessment[1].

The UKHSA describes the NIMS databases as “dynamic database of named individuals, where the numerator and the denominator come from the same source and there is a record of each individual’s vaccination status. Additionally, NIMS contains key sociodemographic variables for those who are targeted and then receive the vaccine, providing a rich and consistently coded data source for evaluation of the vaccine programme. Large scale efforts to contact people in the register will result in the identification of people who may be overcounted, thus affording opportunities to improve accuracy in a dynamic fashion that feeds immediately into vaccine uptake statistics and informs local vaccination efforts”[13].

The UKHSA also describes the advantages and disadvantages of using potential sources of the denominator data from either NIMS or Office for National Statistics (ONS) mid-year population estimates[13]. According the UKHSA, the “NIMS may over-estimate denominators in some age groups, for example because people are registered with the NHS but may have moved abroad. However, as it is a dynamic register, such patients, once identified by the NHS, are able to be removed from the denominator. On the other hand, ONS data uses population estimates based on the 2011 census and other sources of data. When using ONS, vaccine coverage exceeds 100% of the population in some age groups, which would in turn lead to a negative denominator when calculating the size of the unvaccinated population”[13].

Based on the above UKHSA assessment of available databases, we believe the NIMS database population is a reliable population denominator with a possibility of overestimation only when the population moved abroad, and checks and balances to remove them from database are identified. Based on our comparative analysis of VE using the two database denominators, the NIMS underestimates the VE and the England vaccination database overestimates the VE, especially during the period when the VE was waning fast . Additionally, the variation between the database denominators were in low range (4.0% to 13.6%) when the VE was above the FDA cutoff point of greater than 50% and both the database denominators variability is only in 4% range when the VE  is greater than 80%. Based on this data, our use of NIMS database as the denominator may have underestimated the vaccine effectiveness by about 5.0 to 12.5% range during the Omicron variant surge. Since both databases denominators show a negative VE for the third dose, and the cases for 100,000 population among those vaccinated with the third dose are higher than those vaccinated with two doses and unvaccinated during the latter part of the Omicron variant surge since February 2022, it is of utmost importance not to overestimate vaccine effectiveness especially in the elderly population.

Our use of the NIMS denominator did not alter the outcomes reported in our data analysis (SARS-CoV2 cases, hospitalizations and deaths per 100,000 population among the vaccinated populations and their trends in comparison with other vaccinated groups including the unvaccinated population trends). The increasing proportion of SARS-CoV2 cases, hospitalizations, and deaths in the vaccinated population (including the third dose) and decreasing proportion of  SARS-CoV2 cases, hospitalizations, and deaths in the unvaccinated population during the Omicron variant surge as shown in Figure 6 (Table S6a-c) are not affected by the NIMS denominator population.

### Figures and Tables

**Figures S1a: SARS-CoV2 cases per 100,000 population from August 16, 2021-May 27, 2022**

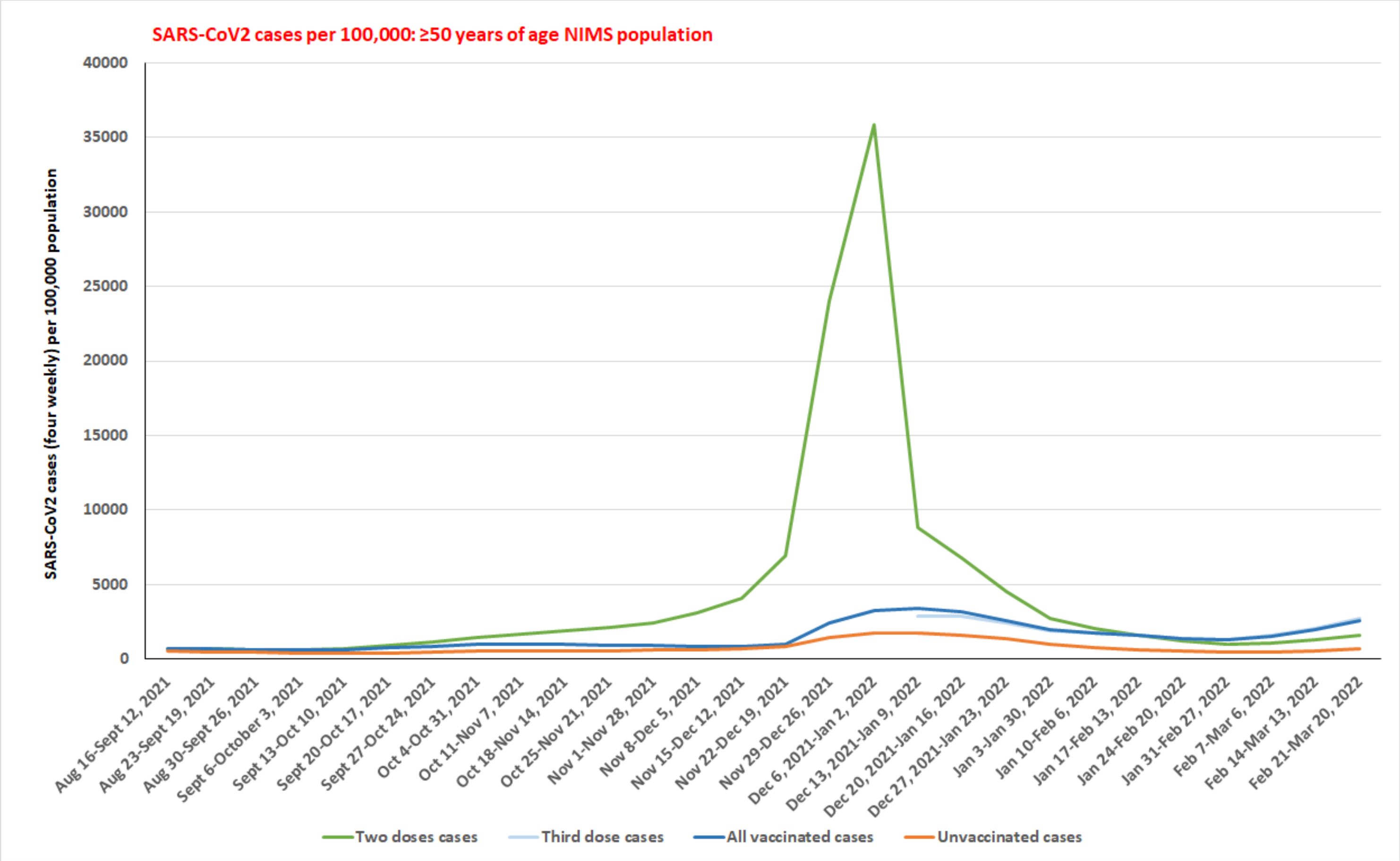

**Figure S1a:** SARS-CoV2 cases per 100,000 population among over 50 years of age group based on the vaccination status from August 16, 2021 to March 27, 2022.

**Figures S1b: SARS-CoV2 hospitalizations per 100,000 population from August 16, 2021-May 27, 2022**

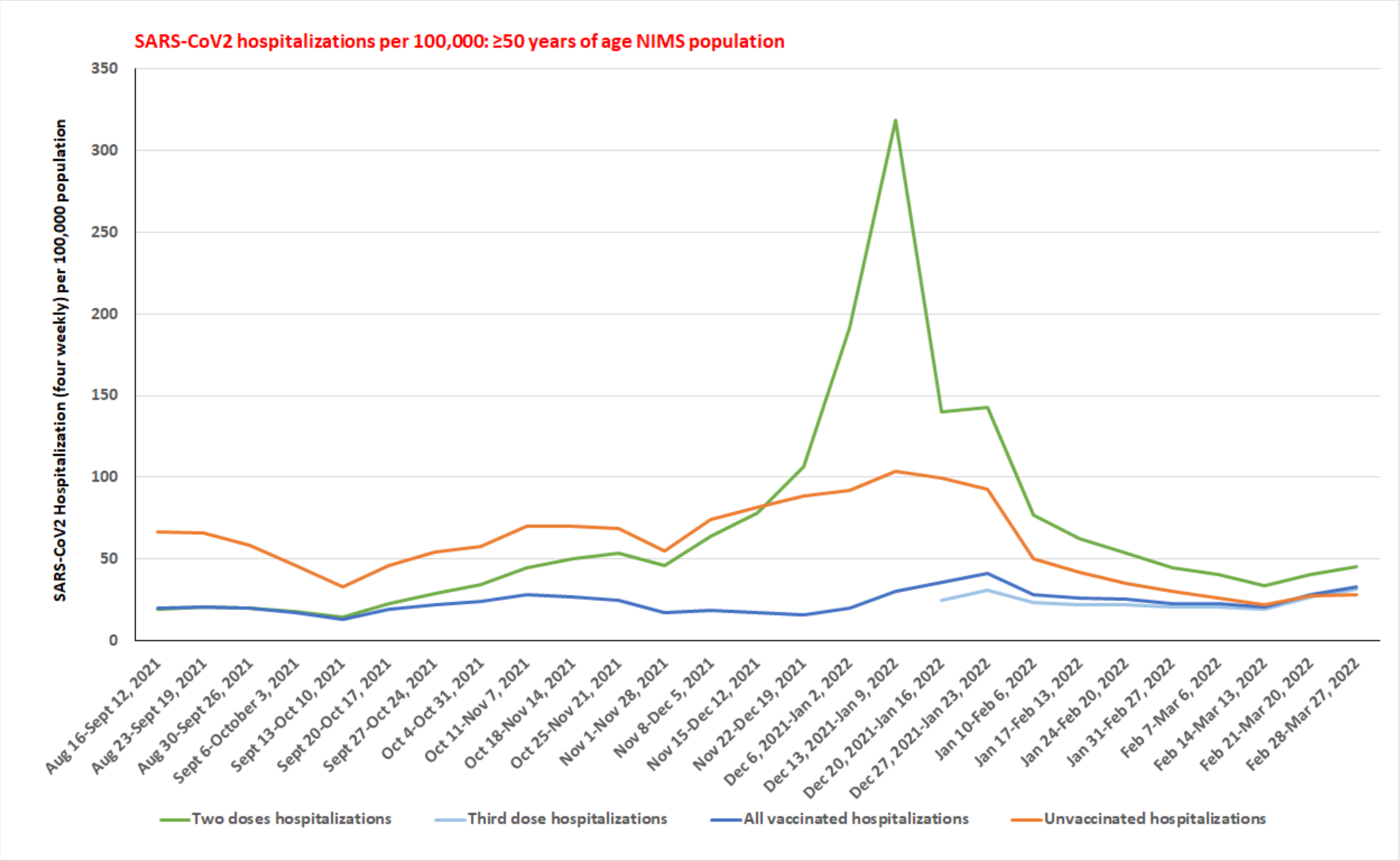

**Figure S1b:** SARS-CoV2 hospitalizations per 100,000 population among over 50 years of age group based on the vaccination status from August 16, 2021 to March 27, 2022.

**Figures S1c: SARS-CoV2 deaths per 100,000 population from August 16, 2021-May 27, 2022**

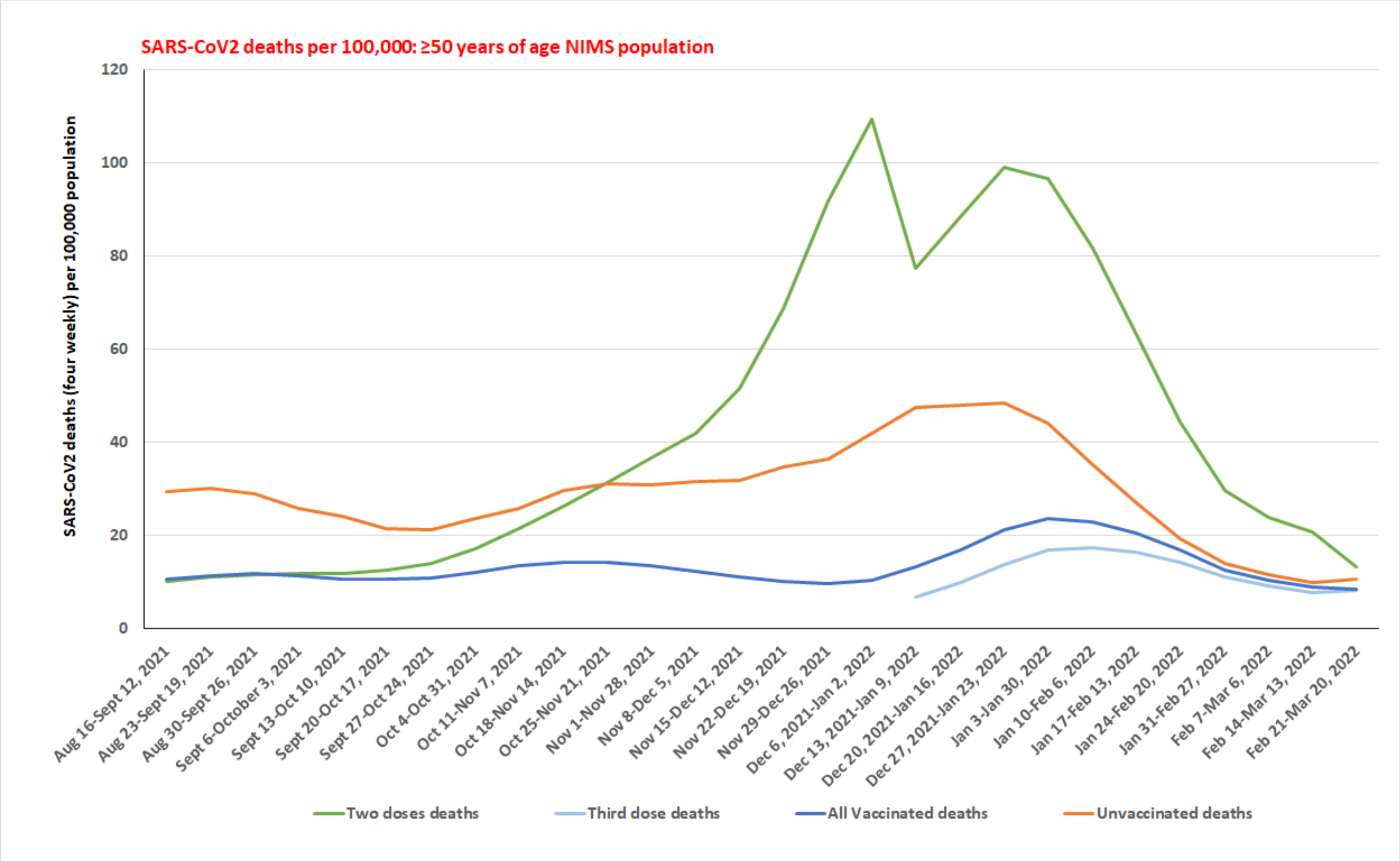

**Figure S1c:** SARS-CoV2 deaths per 100,000 population among over 50 years of age group based on the vaccination status from August 16, 2021 to March 27, 2022.

#### Table S1a-c: SARS-CoV2 cases and hospitalization in England, SARS-CoV2 deaths in England and Wales among age groups

| | Pre-Alpha | Alpha variant surge | | Delta variant surge | | Omicron variant surge | |
|---|---|---|---|---|---|---|---|
| **Study period** | **Sept 28, 2020-Dec 6, 2020** | **Dec 7,2020-Feb 28, 2021** | **Mar 1, 2021-May 23, 2021** | **May 24, 2021-Aug 1, 2021** | **Aug 2, 2021-Dec 5, 2021** | **Dec 6, 2021-Feb 27, 2022** | **Feb 28, 2022-May 1, 2022** |
| **COVID-19 cases; All ages** (n=) | 1,055,142 | 2,074,606 | 235,582 | 1,243,075 | 3,721,600 | 5,459,340 | 2,460,597 |
| **0-19 yrs** (n=%) | 190,310 (18.0%) | 289,018 (13.9%) | 65,475 (27.8%) | 368,060 (29.6%) | 1,395,180 (37.5%) | 1,380,507 (25.3%) | 345,279 (14.0%) |
| **20-29 yrs** (n=%) | 201,561 (19.1%) | 380,818 (18.4%) | 39,470 (16.8%) | 353,755 (28.5%) | 418,792 (11.3%) | 913,345 (16.7%) | 300,745 (12.2%) |
| **30-49 yrs** (n=%) | 332,709 (31.5%) | 728,813 (35.1%) | 81,706 (34.7%) | 355,519 (28.6%) | 1,091,029 (29.3%) | 1,873,509 (34.3%) | 823,063 (22.4%) |
| **50-69 yrs** (n=%) | 238,349 (22.6%) | 487,939 (23.5%) | 38,921 (16.5%) | 138,527 (11.1%) | 655,025 (17.6%) | 994,954 (18.2%) | 687,604 (27.9%) |
| **≥50 yrs** (n=%) | 330,562 (31.3%) | 675,957 (32.6%) | 48,931 (20.8%) | 165,741 (13.3%) | 816,599 (21.9%) | 1,291,979 (23.7%) | 991,510 (40.3%) |
| **≥70 yrs** (n=%) | 92,213 (8.7%) | 188,018 (9.1%) | 10,010 (4.2%) | 27,214 (2.2%) | 161,574 (4.3%) | 297,025 (5.4%) | 303,905 (12.4%) |
| **≥80 yrs** (n=%) | 46,640 (4.4%) | 101,056 (4.9%) | 4,845 (2.1%) | 8,910 (0.7%) | 48,156 (1.3%) | 110,302 (2.0%) | 115,288 (4.7%) |
| **Statistical analysis** | | **(Δ) from prior period** | **(Δ) from prior period** | **(Δ) from prior period** | **(Δ) from prior period** | **(Δ) from prior period** | **(Δ) from prior period** |
| **0-19 yrs** [RR (95%CI); p value] | | 0.77 (0.77-0.78); p<0.001 | 2.00 (1.98-2.01); p<0.001 | 1.07 (1.06-1.07); p<0.001 | 1.27 (1.26-1.27); p<0.001 | 0.67 (0.67-0.68); p<0.001 | 0.55 (0.55-0.56); p<0.001 |
| **20-29 yrs** [RR (95%CI); p value] | | 0.96 (0.96-0.97); p<0.001 | 0.91 (0.90-0.92); p<0.001 | 1.70 (1.68-1.71); p<0.001 | 0.40 (0.39-0.40); p<0.001 | 1.48 (1.48-1.49); p<0.001 | 0.73 (0.72-0.73); p<0.001 |
| **30-49 yrs** [RR (95%CI); p value] | | 1.11 (1.11-1.12); p<0.001 | 0.99 (0.98-0.99); p<0.001 | 0.82 (0.82-0.83); p<0.001 | 1.03 (1.02-1.03); p<0.001 | 1.17 (1.168-1.172); p<0.001 | 0.97 (0.97-0.98); p<0.001 |
| **50-69 yrs** [RR (95%CI); p value] | | 1.04 (1.04-1.05); p<0.001 | 0.70 (0.69-0.71); p<0.001 | 0.67 (0.66-0.68); p<0.001 | 1.58 (1.57-1.59); p<0.001 | 1.03 (1.03-1.04); p<0.001 | 1.53 (1.52-1.54); p<0.001 |
| **≥50 years** [RR (95%CI); p value] | | 1.04 (1.03-1.04); p<0.001 | 0.64 (0.63-0.64); p<0.001 | 0.64 (0.63-0.65); p<0.001 | 1.65 (1.63-1.65); p<0.001 | 1.07 (1.07-1.08); p<0.001 | 1.70 (1.70-1.71); p<0.001 |
| **≥70 yrs** [RR (95%CI); p value] | | 1.04 (1.03-1.04); p<0.001 | 0.47 (0.46-0.48); p<0.001 | 0.52 (0.50-0.53); p<0.001 | 1.98 (1.95-2.01); p<0.001 | 1.25 (1.24-1.26); p<0.001 | 2.27 (2.26-2.28); p<0.001 |
| **≥80 yrs** [RR (95%CI); p value] | | 1.10 (1.09-1.11); p<0.001 | 0.42 (0.41-0.43); p<0.001 | 0.35 (0.37-0.36); p<0.001 | 1.81 (1.76-1.85); p<0.001 | 1.56 (1.54-1.58); p<0.001 | 2.32 (2.30-2.34); p<0.001 |

**Table S1a: SARS-CoV2 cases in England among age groups in between September 28, 2020 to May 1, 2022.** The total SARS-CoV2 cases for each study period and the proportion of SARS-CoV2 cases among the various age groups (n=%) for the specific period shown on the table. The statistical analysis is shown on the lower portion of table for the specified time period. The analysis provides the proportion of change (Δ) in SARS-CoV2 infections in a particular age group compared to the prior period represented as relative risk (RR), 95% confidence interval (95% CI) and p-value.

| | Pre-Alpha | Alpha variant surge | | Delta variant surge | | Omicron variant surge | |
|---|---|---|---|---|---|---|---|
| **Time period** | **Oct 12, 2020-Dec 6, 2020** | **Dec 7, 2020-Feb 28, 2021** | **Mar 1, 2021-May 23, 2021** | **May 24-Aug 1, 2021** | **Aug 2-Dec 5, 2021** | **Dec 6,2021-Feb 27,2022** | **Feb 28-May 1, 2022** |
| **COVID-19 Hospitalizations; All ages (n=)** | 65,419 | 179,599 | 17,295 | 24,880 | 87,823 | 106,834 | 95,278 |
| **0-17 yrs** (n=%) | 952 (1.5%) | 2,496 (1.4%) | 599 (3.5%) | 1,602 (6.4%) | 4,779 (5.4%) | 7,396 (6.9%) | 4,971 (5.2%) |
| **18-34 yrs** (n=%) | 3,440 (5.3%) | 9,942 (5.5%) | 1,924 (11.1%) | 5,784 (23.2%) | 9,994 (11.4%) | 13,775 (12.9%) | 7,933 (8.3%) |
| **35-54 yrs** (n=%) | 8,970 (13.7%) | 29,483 (16.4%) | 3,759 (21.7%) | 6,831 (27.5%) | 18,847 (21.5%) | 16,710 (15.6%) | 10,362 (10.9%) |
| **55-64 yrs** (n=%) | 8,669 (13.3%) | 28,208 (15.7%) | 2,697 (15.6%) | 3,028 (12.2%) | 12,189 (13.9%) | 11,580 (10.8%) | 9,210 (9.7%) |
| **65-74 yrs** (n=%) | 12,107 (18.5%) | 32,426 (18.1%) | 2,540 (14.7%) | 2,770 (11.1%) | 14,944 (17.0%) | 15,466 (14.5%) | 14,794 (15.5%) |
| **≥75 yrs** (n=%) | 30,586 (46.8%) | 76,831 (42.8%) | 5,761 (33.3%) | 4,806 (19.3%) | 27,047 (30.8%) | 41,875 (39.2%) | 47,970 (50.3%) |
| **Statistical analysis** | | **(Δ) from prior period** | **(Δ) from prior period** | **(Δ) from prior period** | **(Δ) from prior period** | **(Δ) from prior period** | **(Δ) from prior period** |
| **0-17 yrs** [RR (95% CI); p value] | | 0.96 (0.88-1.02); p<0.223 | 2.49 (2.28-2.72); p<0.001 | 1.86 (1.70-2.03); p<0.001 | 0.85 (0.80-0.89); p<0.001 | 1.27 (1.22-1.31); p<0.001 | 0.75 (0.72-0.78); p<0.001 |
| **18-34 yrs** [RR (95% CI); p value] | | 1.05 (1.01-1.09); p=0.007 | 2.01 (1.91-2.10); p<0.001 | 2.09 (1.99-2.19); p<0.001 | 0.49 (0.47-0.50); p<0.001 | 1.13 (1.10-1.16); p<0.001 | 0.65 (0.62-0.66); p<0.001 |
| **35-54 yrs** [RR (95% CI); p value] | | 1.20 (1.17-1.22); p<0.001 | 1.32 (1.28-1.36); p<0.001 | 1.26 (1.22-1.31); p<0.001 | 0.78 (0.76-0.80); p<0.001 | 0.73 (0.71-0.74); p<0.001 | 0.70 (0.67-0.71); p<0.001 |
| **55-64 yrs** [RR (95% CI); p value] | | 1.19 (1.16-1.21); p<0.001 | 0.99 (0.95-1.03); p=0.699 | 0.78 (0.74-0.82); p<0.001 | 1.14 (1.10-1.18); p<0.001 | 0.78 (0.76-0.80); p<0.001 | 0.89 (0.86-0.91); p<0.001 |
| **65-74 yrs** [RR (95% CI); p value] | | 0.98 (0.95-0.99); p=0.010 | 0.81 (0.78-0.84); p<0.001 | 0.76 (0.72-0.80); p<0.001 | 1.53 (1.47-1.59); p<0.001 | 0.85 (0.83-0.87); p<0.001 | 1.07 (1.05-1.09); p<0.001 |
| **≥75 yrs** [RR (95% CI); p value] | | 0.92 (0.90-0.92); p<0.001 | 0.78 (0.76-0.80); p<0.001 | 0.58 (0.56-0.60); p<0.001 | 1.59 (1.55-1.64); p<0.001 | 1.27 (1.25-1.29); p<0.001 | 1.28 (1.27-1.30); p<0.001 |

**Table S1b: SARS-CoV2 hospitalizations in England among age groups in between October 12, 2020 to May 1, 2022.** The total SARS-CoV2 hospitalizations for each study period and the proportion of SARS-CoV2 hospitalizations among the various age groups (n=%) for the specific period shown on the table. The statistical analysis is shown on the lower portion of table for the specified time period. The analysis provides the proportion of change (Δ) in SARS-CoV2 hospitalizations in a particular age group compared to the prior period represented as relative risk (RR), 95% confidence interval (95% CI) and p-value.

| | First wave | Pre-alfa | Alpha variant surge | | Delta variant surge | | Omicron variant surge | |
|---|---|---|---|---|---|---|---|---|
| **Study period** | **Feb 24-Aug 16, 2020** | **Aug 17-Dec 6, 2020** | **Dec 7,2020-Feb 28, 2021** | **Mar 1, 2021-May 23, 2021** | **May 24, 2021-Aug 1, 2021** | **Aug 2, 2021-Dec 5, 2021** | **Dec 6, 2021-Feb 27, 2022** | **Feb 28, 2022-May 1, 2022** |
| **COVID-19 deaths, all ages (n=)** | 52,410 | 19,082 | 63,118 | 7,281 | 1,719 | 13,980 | 12,063 | 7,787 |
| **0-19 yrs (n= %)** | 15 (0.03%) | 2 (0.01%) | 16 (0.03%) | 4 (0.05%) | 6 (0.35%) | 33 (0.24%) | 27 (0.22%) | 12 (0.15%) |
| **20-29 yrs (n= %)** | 76 (0.15%) | 21 (0.11%) | 74 (0.12%) | 16 (0.22%) | 14 (0.81%) | 51 (0.36%) | 35 (0.29%) | 14 (0.18%) |
| **30-49 yrs (n= %)** | 886 (1.69%) | 266 (1.39%) | 1,153 (1.83%) | 241 (3.31%) | 114 (6.63%) | 611 (4.37%) | 399 (3.31%) | 118 (1.52%) |
| **50-69 yrs (n= %)** | 7,468 (14.25%) | 2,600 (13.63%) | 9,887 (15.66%) | 1,785 (24.52%) | 468 (27.23%) | 3,302 (23.62%) | 2,241 (18.58%) | 793 (10.18%) |
| **70-74 yrs (n= %)** | 4,817 (9.19%) | 1,956 (10.25%) | 6,077 (9.63%) | 708 (9.72%) | 166 (9.66%) | 1,619 (11.58%) | 1,933 (16.02%) | 599 (7.69%) |
| **≥ 50 yrs (n= %)** | 51345 (97.97%) | 18,765 (98.34%) | 61,771 (97.87%) | 6,987 (95.96%) | 1,570 (91.33%) | 13,215 (94.53%) | 11,546 (95.71%) | 7,627 (97.95%) |
| **≥ 75 yrs (n= %)** | 39060 (74.53%) | 14,209 (74.46%) | 45,807 (72.57%) | 4,494 (61.72%) | 936 (54.45%) | 8,294 (59.33%) | 8,189 (67.89%) | 6,235 (80.07%) |
| **Statistical analysis** | | **(Δ) from prior period** | **(Δ) from prior period** | **(Δ) from prior period** | **(Δ) from prior period** | **(Δ) from prior period** | **(Δ) from prior period** | **(Δ) from prior period** |
| **0-19 yrs** [RR (95% CI); p value] | | 0.36 (0.08-1.60); p=0.182 | 2.42 (0.55-10.51); p=0.238 | 2.17 (0.72-6.48); p=0.166 | 6.35 (1.79-22.49); p=0.004 | 0.68 (0.28-1.61); p=0.377 | 0.95 (0.57-1.58); p=0.837 | 0.69 (0.35-1.36); p=0.281 |
| **20-29 yrs** [RR (95% CI); p value] | | 0.75 (0.46-1.23); p=0.263 | 1.07 (0.66-1.73); p=0.797 | 1.87 (1.09-3.22); p=0.022 | 3.71 (1.81-7.57); p=0.0003 | 0.45 (0.24-0.80); p=0.007 | 0.80 (0.51-1.22); p=0.296 | 0.62 (0.33-1.15); p=0.130 |
| **30-49 yrs** [RR (95% CI); p value] | | 0.82 (0.72-0.944); p=0.005 | 1.31 (1.15-1.49); p=0.001 | 1.81 (1.58-2.08); p<0.001 | 2.00 (1.61-2.49); p<0.001 | 0.66 (0.54-0.80); p<0.001 | 0.76 (0.66-0.86); p<0.001 | 0.46 (0.37-0.56); p<0.001 |
| **50-69 yrs** [RR (95% CI); p value] | | 0.96 (0.92-0.99); p=0.034 | 1.15 (1.10-1.12); p<0.001 | 1.57 (1.49-1.64); p<0.001 | 1.11 (1.02-1.21); p=0.018 | 0.87 (0.79-0.94); p=0.001 | 0.79 (0.75-0.82); p<0.001 | 0.41 (0.38-0.45); p<0.001 |
| **70-74 yrs** [RR (95% CI); p value] | | 1.11 (1.06-1.17); p<0.001 | 0.94 (0.89-0.98); p=0.011 | 1.01 (0.93-1.09); p=0.792 | 0.99 (0.84-1.17); p=0.932 | 1.20 (1.03-1.39); p=0.019 | 1.38 (1.30-1.47); p<0.001 | 0.48 (0.44-0.52); p<0.001 |
| **≥ 50 yrs** [RR (95% CI); p value] | | 1.00 (1.00-1.01); p=0.001 | 0.995 (0.993-0.997); p<0.001 | 0.98 (0.97-0.99); p<0.001 | 0.95 (0.93-0.97); p<0.001 | 1.04 (1.01-1.05); p<0.001 | 1.01 (1.01-1.02); p<0.001 | 1.02 (1.02-0.03); p<0.001 |
| **≥ 75 yrs** [RR (95% CI); p value] | | 0.99 (0.98-1.01); p=0.860 | 0.97 (0.96-0.98); p<0.001 | 0.85 (0.83-0.87); p<0.001 | 0.88 (0.84-0.92); p<0.001 | 1.09 (1.04-1.14); p=0.001 | 1.14 (1.12-1.17); p<0.001 | 1.18 ((1.16-1.20); p<0.001 |

**Table S1c: SARS-CoV2 deaths in England and Wales among age groups in between February 24, 2020 to May 1, 2022.** The total SARS-CoV2 deaths for each study period and the proportion of SARS-CoV2 deaths among the various age groups (n=%) for the specific period shown on the table. The statistical analysis is shown on the lower portion of table for the specified time period. The analysis provides the proportion of change (Δ) in SARS-CoV2 deaths in a particular age group compared to the prior period represented as relative risk (RR), 95% confidence interval (95% CI) and p-value.

**Table S2:** **Pre-existing condition among deaths during the Omicron variant surge in England**

| Pre-existing conditions during the Omicron variant surge (December 6, 2021-May 1, 2022) | | | |
|---|---|---|---|
| **Age group (yrs)** | **Total SARS-COV2 deaths** | **Pre-existing conditions among the SARS-CoV2 deaths** | |
| | | **Present** | **Absent** |
| **All ages** | 17,142 | 16,387 (95.6%) | 755 (4.4%) |
| **0 - 19** | 39 | 27 (69.2%) | 12 (30.8%) |
| **20 - 39** | 198 | 164 (82.8%) | 34 (17.2%) |
| **40 - 59** | 1,206 | 1,093 (90.6%) | 113 (9.4%) |
| **60 - 79** | 5,803 | 5,537 (95.4%) | 266 (4.6%) |
| **80+** | 9,895 | 9,566 (96.7%) | 329 (3.3%) |
| **60+** | 15,698 | 15,103 (96.2%) | 595 (3.8%) |
| **Pre-existing conditions during the entire pandemic** | | | |
| **Date introduced** | **Pre-existing condition** | **Count of all deaths since condition introduced** | **Deaths with pre-existing condition (n=%)** |
| **31-Mar-20** | **Diabetes** | 115,463 | 29,572 (25.6%) |
| **31-Mar-20** | **Chronic Kidney Disease** | 115,463 | 19,548 (16.9%) |
| **31-Mar-20** | **Chronic Pulmonary Disease** | 115,463 | 18,455 (16.0%) |
| **31-Mar-20** | **Dementia** | 115,463 | 17,011 (14.7%) |
| **1-May-20** | **Ischaemic Heart Disease** | 96,645 | 12,468 (12.9%) |
| **31-Mar-20** | **Asthma** | 115,463 | 8,690 (7.5%) |
| **31-Mar-20** | **Rheumatological Disorder** | 115,463 | 3,904 (3.4%) |
| **31-Mar-20** | **Chronic Neurological Disorder** | 115,463 | 2,934 (2.5%) |
| **24-Mar-20** | **Received treatment for a Mental Health condition** | 117,059 | 2,653 (2.3%) |

| | | | |
|---|---|---|---|
| **24-Mar-20** | **Learning Disability and or Autism** | 117,059 | 1,397 (1.2%) |
| **31-Mar-20** | **Other** | 115,463 | 84,544 (73.2%) |

Table S2: **Pre-existing condition among deaths during the Omicron variant surge in England.** The lower portion of the table represent the cumulative (n=%) for each diagnosis since the condition date was introduced for the analysis. The age specific breakdown of the pre-existing condition diagnosis is not available. However, 95.71% to 97.5% of the total SARS-CoV2 deaths occurred among the over 50 years of age during the Omicron variant surge as noted in the Table S1c. During the initial and latter part of the Omicron variant surge 67.89% and 80.07% of total deaths occurred respectively among those over 75 years of age as shown the same table.

**Table S3a:** **NIMS database with vaccination status of all age groups , over 18 and 50 years of age**

| week ending | All ages, vaccination status; n=population (%) | | | | ≥ 18 yrs. of age vaccination status; n=population (%) | | | | ≥ 50 yrs. of age vaccination status; n=population (%) | | | |
|---|---|---|---|---|---|---|---|---|---|---|---|---|
| | 2 doses (no 3rd) | Third dose | 1 dose only | Unvaccinated | 2 doses (no 3rd) | Third dose | 1 dose only | Unvaccinated | 2 doses (no 3rd) | Third dose | 1 dose only | Unvaccinated |
| 3-Jan-21 | 29,783 (0.05%) | | 1,108,851 (1.8%) | 61,961,296 (98.2%) | 29,775 (0.1%) | | 1,108,534 (2.2%) | 49,085,900 (97.7%) | 18,145 (0.1%) | | 873,796 (3.9%) | 21,607,893 (96.0%) |
| 10-Jan-21 | 402,102 (0.6%) | | 1,661,407 (2.6%) | 61,036,379 (96.7%) | 402,058 (0.8%) | | 1,660,718 (3.3%) | 48,161,391 (95.9%) | 354,641 (1.6%) | | 1,207,864 (5.4%) | 20,937,311 (93.1%) |
| 17-Jan-21 | 442,698 (0.7%) | | 3,174,888 (5.0%) | 59,482,270 (94.3%) | 442,647 (0.9%) | | 3,173,666 (6.3%) | 46,607,822 (92.8%) | 384,361 (1.7%) | | 2,349,075 (10.4%) | 19,766,359 (87.9%) |
| 24-Jan-21 | 450,064 (0.7%) | | 5,359,396 (8.5%) | 57,290,372 (90.8%) | 450,010 (0.9%) | | 5,357,481 (10.7%) | 44,416,620 (88.4%) | 388,937 (1.7%) | | 4,123,495 (18.3%) | 17,987,350 (79.9%) |
| 31-Jan-21 | 460,029 (0.7%) | | 7,513,401 (11.9%) | 55,126,358 (87.4%) | 459,963 (0.9%) | | 7,510,529 (15.0%) | 42,253,575 (84.1%) | 394,382 (1.8%) | | 5,924,604 (26.3%) | 16,180,765 (71.9%) |
| 7-Feb-21 | 470,877 (0.7%) | | 9,874,536 (15.6%) | 52,754,344 (83.6%) | 470,811 (0.9%) | | 9,870,576 (19.7%) | 39,882,649 (79.4%) | 400,108 (1.8%) | | 7,960,696 (35.4%) | 14,138,919 (62.8%) |
| 14-Feb-21 | 490,579 (0.8%) | | 12,141,763 (19.2%) | 50,467,364 (80.0%) | 490,500 (1.0%) | | 12,136,095 (24.2%) | 37,597,390 (74.9%) | 411,497 (1.8%) | | 9,906,334 (44.0%) | 12,181,858 (54.1%) |
| 21-Feb-21 | 515,699 (0.8%) | | 14,167,952 (22.5%) | 48,416,011 (76.7%) | 515,602 (1.0%) | | 14,157,869 (28.2%) | 35,550,470 (70.8%) | 425,363 (1.9%) | | 11,453,744 (50.9%) | 10,620,547 (47.2%) |
| 28-Feb-21 | 606,844 (1.0%) | | 16,326,976 (25.9%) | 46,165,791 (73.2%) | 606,671 (1.2%) | | 16,307,128 (32.5%) | 33,310,091 (66.3%) | 477,899 (2.1%) | | 12,897,769 (57.3%) | 9,123,957 (40.6%) |
| 7-Mar-21 | 804,235 (1.3%) | | 17,915,499 (28.4%) | 44,379,802 (70.3%) | 803,960 (1.6%) | | 17,892,589 (35.6%) | 31,527,266 (62.8%) | 619,281 (2.8%) | | 13,951,393 (62.0%) | 7,928,897 (35.2%) |
| 14-Mar-21 | 1,117,297 (1.8%) | | 19,298,912 (30.6%) | 42,683,207 (67.6%) | 1,116,884 (2.2%) | | 19,272,701 (38.4%) | 29,834,110 (59.4%) | 837,736 (3.7%) | | 14,951,636 (66.5%) | 6,710,129 (29.8%) |
| 21-Mar-21 | 1,612,580 (2.6%) | | 21,881,719 (34.7%) | 39,604,914 (62.8%) | 1,611,856 (3.2%) | | 21,852,258 (43.5%) | 26,759,378 (53.3%) | 1,168,952 (5.2%) | | 16,952,088 (75.3%) | 4,378,368 (19.5%) |
| 28-Mar-21 | 2,799,963 (4.4%) | | 22,724,932 (36.0%) | 37,574,058 (59.5%) | 2,798,602 (5.6%) | | 22,689,611 (45.2%) | 24,735,019 (49.2%) | 2,061,993 (9.2%) | | 17,281,497 (76.8%) | 3,155,746 (14.0%) |
| 4-Apr-21 | 4,323,280 (6.9%) | | 22,014,103 (34.9%) | 36,761,369 (58.3%) | 4,321,262 (8.6%) | | 21,975,777 (43.8%) | 23,925,992 (47.6%) | 3,275,787 (14.6%) | | 16,413,863 (73.0%) | 2,809,437 (12.5%) |
| 11-Apr-21 | 6,297,786 (10.0%) | | 20,384,850 (32.3%) | 36,415,884 (57.7%) | 6,294,836 (12.5%) | | 20,344,816 (40.5%) | 23,583,147 (47.0%) | 4,894,124 (21.8%) | | 14,902,291 (66.2%) | 2,702,514 (12.0%) |

| | | | | | | | | | | | | |
|---|---|---|---|---|---|---|---|---|---|---|---|---|
| **18-Apr-21** | 8,436,940 (13.4%) | | 18,769,544 (29.7%) | 35,891,780 (56.9%) | 8,432,628 (16.8%) | | 18,728,628 (37.3%) | 23,061,287 (45.9%) | 6,685,917 (29.7%) | | 13,190,927 (58.6%) | 2,621,896 (11.7%) |
| **25-Apr-21** | 10,680,337 (16.9%) | | 17,186,764 (27.2%) | 35,230,749 (55.8%) | 10,674,312 (21.3%) | | 17,145,979 (34.1%) | 22,401,838 (44.6%) | 8,595,553 (38.2%) | | 11,356,042 (50.5%) | 2,546,853 (11.3%) |
| **2-May-21** | 12,822,577 (20.3%) | | 15,718,252 (24.9%) | 34,556,605 (54.8%) | 12,814,312 (25.5%) | | 15,678,228 (31.2%) | 21,729,174 (43.3%) | 10,380,479 (46.1%) | | 9,621,020 (42.8%) | 2,496,624 (11.1%) |
| **9-May-21** | 14,872,852 (23.6%) | | 14,359,970 (22.8%) | 33,864,269 (53.7%) | 14,859,446 (29.6%) | | 14,323,504 (28.5%) | 21,038,422 (41.9%) | 11,930,952 (53.0%) | | 8,108,973 (36.0%) | 2,457,945 (10.9%) |
| **16-May-21** | 17,003,352 (26.9%) | | 13,255,207 (21.0%) | 32,838,081 (52.0%) | 16,979,886 (33.8%) | | 13,223,273 (26.3%) | 20,017,762 (39.9%) | 13,397,393 (59.6%) | | 6,692,260 (29.7%) | 2,407,916 (10.7%) |
| **23-May-21** | 19,266,071 (30.5%) | | 12,201,728 (19.3%) | 31,628,350 (50.1%) | 19,238,435 (38.3%) | | 12,169,339 (24.2%) | 18,812,657 (37.5%) | 15,002,863 (66.7%) | | 5,141,253 (22.9%) | 2,353,113 (10.5%) |
| **30-May-21** | 21,501,133 (34.1%) | | 11,106,521 (17.6%) | 30,487,981 (48.3%) | 21,469,883 (42.7%) | | 11,074,839 (22.1%) | 17,675,195 (35.2%) | 16,624,527 (73.9%) | | 3,585,879 (15.9%) | 2,286,459 (10.2%) |
| **6-Jun-21** | 23,480,377 (37.2%) | | 9,997,612 (15.8%) | 29,617,087 (46.9%) | 23,446,395 (46.7%) | | 9,964,995 (19.8%) | 16,807,969 (33.5%) | 17,991,864 (80.0%) | | 2,251,444 (10.0%) | 2,253,174 (10.0%) |
| **13-Jun-21** | 25,151,891 (39.9%) | | 9,383,450 (14.9%) | 28,558,988 (45.3%) | 25,113,935 (50.0%) | | 9,350,595 (18.6%) | 15,754,083 (31.4%) | 18,951,557 (84.2%) | | 1,317,673 (5.9%) | 2,226,792 (9.9%) |
| **20-Jun-21** | 26,287,679 (41.7%) | | 9,504,211 (15.1%) | 27,301,589 (43.3%) | 26,246,201 (52.3%) | | 9,469,778 (18.9%) | 14,501,789 (28.9%) | 19,393,719 (86.2%) | | 900,099 (4.0%) | 2,201,669 (9.8%) |
| **27-Jun-21** | 27,168,998 (43.1%) | | 9,798,677 (15.5%) | 26,124,962 (41.4%) | 27,124,121 (54.0%) | | 9,763,129 (19.4%) | 13,329,676 (26.5%) | 19,567,593 (87.0%) | | 750,506 (3.3%) | 2,176,842 (9.7%) |
| **4-Jul-21** | 28,061,613 (44.5%) | | 9,648,230 (15.3%) | 25,382,133 (40.2%) | 28,013,518 (55.8%) | | 9,611,624 (19.1%) | 12,591,124 (25.1%) | 19,666,121 (87.4%) | | 671,838 (3.0%) | 2,156,617 (9.6%) |
| **11-Jul-21** | 28,950,150 (45.9%) | | 9,242,451 (14.6%) | 24,898,854 (39.5%) | 28,898,876 (57.5%) | | 9,205,675 (18.3%) | 12,111,194 (24.1%) | 19,739,279 (87.8%) | | 617,709 (2.7%) | 2,137,378 (9.5%) |
| **18-Jul-21** | 29,966,247 (47.5%) | | 8,554,088 (13.6%) | 24,570,647 (38.9%) | 29,911,287 (59.6%) | | 8,517,418 (17.0%) | 11,786,568 (23.5%) | 19,798,536 (88.0%) | | 575,609 (2.6%) | 2,120,034 (9.4%) |
| **25-Jul-21** | 30,932,504 (49.0%) | | 7,821,016 (12.4%) | 24,336,996 (38.6%) | 30,874,691 (61.5%) | | 7,783,137 (15.5%) | 11,556,979 (23.0%) | 19,843,303 (88.2%) | | 543,956 (2.4%) | 2,106,761 (9.4%) |
| **1-Aug-21** | 31,852,321 (50.5%) | | 7,098,507 (11.2%) | 24,139,246 (38.3%) | 31,791,048 (63.3%) | | 7,058,043 (14.1%) | 11,365,276 (22.6%) | 19,883,636 (88.4%) | | 515,571 (2.3%) | 2,094,646 (9.3%) |
| **8-Aug-21** | 32,756,024 (51.9%) | | 6,373,113 (10.1%) | 23,960,443 (38.0%) | 32,691,199 (65.1%) | | 6,320,771 (12.6%) | 11,201,905 (22.3%) | 19,916,778 (88.5%) | | 493,433 (2.2%) | 2,083,467 (9.3%) |
| **15-Aug-21** | 33,727,545 (53.5%) | | 5,611,293 (8.9%) | 23,750,160 (37.6%) | 33,659,120 (67.0%) | | 5,490,998 (10.9%) | 11,063,176 (22.0%) | 19,944,748 (88.7%) | | 475,305 (2.1%) | 2,073,405 (9.2%) |
| **22-Aug-21** | 34,687,269 (55.0%) | | 4,942,094 (7.8%) | 23,459,028 (37.2%) | 34,615,292 (68.9%) | | 4,655,978 (9.3%) | 10,941,417 (21.8%) | 19,969,187 (88.8%) | | 459,499 (2.0%) | 2,064,546 (9.2%) |

| | | | | | | | | | | | | |
|---|---|---|---|---|---|---|---|---|---|---|---|---|
| **29-Aug-21** | 35,479,763 (56.2%) | | 4,423,944 (7.0%) | 23,184,181 (36.7%) | 35,404,658 (70.5%) | | 3,972,304 (7.9%) | 10,835,225 (21.6%) | 19,992,043 (88.9%) | | 444,528 (2.0%) | 2,056,504 (9.1%) |
| **5-Sep-21** | 36,118,118 (57.2%) | 12,738 (0.02%) | 3,977,545 (6.3%) | 22,991,593 (36.4%) | 36,039,895 (71.8%) | 12,713 (0.03%) | 3,428,983 (6.8%) | 10,742,682 (21.4%) | 20,008,541 (88.9%) | 7,045 (0.03%) | 434,345 (1.9%) | 2,049,929 (9.1%) |
| **12-Sep-21** | 36,632,606 (58.1%) | 14,276 (0.02%) | 3,593,049 (5.7%) | 22,860,063 (36.2%) | 36,551,529 (72.8%) | 14,248 (0.03%) | 3,005,016 (6.0%) | 10,653,480 (21.2%) | 20,024,672 (89.0%) | 7,995 (0.04%) | 423,866 (1.9%) | 2,043,327 (9.1%) |
| **19-Sep-21** | 36,984,090 (58.6%) | 43,185 (0.1%) | 3,326,622 (5.3%) | 22,746,097 (36.0%) | 36,899,865 (73.5%) | 43,126 (0.1%) | 2,709,140 (5.4%) | 10,572,142 (21.0%) | 20,018,492 (89.0%) | 30,591 (0.1%) | 414,203 (1.8%) | 2,036,574 (9.1%) |
| **26-Sep-21** | 36,865,266 (58.4%) | 432,235 (0.7%) | 3,178,256 (5.0%) | 22,624,237 (35.9%) | 36,777,082 (73.2%) | 432,070 (0.9%) | 2,518,065 (5.0%) | 10,497,056 (20.9%) | 19,709,295 (87.6%) | 357,852 (1.6%) | 402,771 (1.8%) | 2,029,942 (9.0%) |
| **3-Oct-21** | 36,136,121 (57.3%) | 1,360,104 (2.2%) | 3,162,512 (5.0%) | 22,441,257 (35.6%) | 36,043,135 (71.8%) | 1,359,758 (2.7%) | 2,387,178 (4.8%) | 10,434,202 (20.8%) | 18,940,073 (84.2%) | 1,145,668 (5.1%) | 389,978 (1.7%) | 2,024,141 (9.0%) |
| **10-Oct-21** | 35,097,679 (55.6%) | 2,575,568 (4.1%) | 3,202,469 (5.1%) | 22,224,278 (35.2%) | 34,992,715 (69.7%) | 2,574,955 (5.1%) | 2,287,832 (4.6%) | 10,368,771 (20.6%) | 17,922,164 (79.7%) | 2,179,876 (9.7%) | 380,041 (1.7%) | 2,017,779 (9.0%) |
| **17-Oct-21** | 33,924,691 (53.8%) | 3,913,065 (6.2%) | 3,278,275 (5.2%) | 21,983,963 (34.8%) | 33,800,596 (67.3%) | 3,912,076 (7.8%) | 2,206,454 (4.4%) | 10,305,147 (20.5%) | 16,792,820 (74.6%) | 3,323,957 (14.8%) | 371,757 (1.7%) | 2,011,326 (8.9%) |
| **24-Oct-21** | 32,526,363 (51.5%) | 5,461,045 (8.7%) | 3,382,332 (5.4%) | 21,730,254 (34.4%) | 32,383,551 (64.5%) | 5,459,513 (10.9%) | 2,141,948 (4.3%) | 10,239,261 (20.4%) | 15,452,297 (68.7%) | 4,679,006 (20.8%) | 364,458 (1.6%) | 2,004,099 (8.9%) |
| **31-Oct-21** | 31,082,160 (49.3%) | 7,035,752 (11.2%) | 3,464,949 (5.5%) | 21,517,133 (34.1%) | 30,921,587 (61.6%) | 7,033,556 (14.0%) | 2,086,992 (4.2%) | 10,182,138 (20.3%) | 14,066,962 (62.5%) | 6,077,670 (27.0%) | 358,133 (1.6%) | 1,997,095 (8.9%) |
| **7-Nov-21** | 29,391,860 (46.6%) | 8,847,251 (14.0%) | 3,601,201 (5.7%) | 21,259,682 (33.7%) | 29,217,613 (58.2%) | 8,844,063 (17.6%) | 2,034,479 (4.1%) | 10,128,118 (20.2%) | 12,477,515 (55.5%) | 7,680,221 (34.1%) | 352,272 (1.6%) | 1,989,852 (8.8%) |
| **14-Nov-21** | 27,527,382 (43.6%) | 10,841,337 (17.2%) | 3,684,054 (5.8%) | 21,047,221 (33.4%) | 27,339,403 (54.4%) | 10,836,403 (21.6%) | 1,977,027 (3.9%) | 10,071,440 (20.1%) | 10,769,333 (47.9%) | 9,403,246 (41.8%) | 345,025 (1.5%) | 1,982,256 (8.8%) |
| **21-Nov-21** | 25,605,749 (40.6%) | 12,884,764 (20.4%) | 3,723,869 (5.9%) | 20,885,612 (33.1%) | 25,402,404 (50.6%) | 12,877,488 (25.6%) | 1,927,847 (3.8%) | 10,016,534 (19.9%) | 9,072,178 (40.3%) | 11,116,352 (49.4%) | 337,097 (1.5%) | 1,974,233 (8.8%) |
| **28-Nov-21** | 23,621,275 (37.4%) | 15,029,384 (23.8%) | 3,695,318 (5.9%) | 20,754,017 (32.9%) | 23,366,748 (46.5%) | 15,019,488 (29.9%) | 1,876,521 (3.7%) | 9,961,516 (19.8%) | 7,387,515 (32.8%) | 12,824,311 (57.0%) | 321,984 (1.4%) | 1,966,050 (8.7%) |
| **5-Dec-21** | 21,594,191 (34.2%) | 17,253,869 (27.3%) | 3,632,098 (5.8%) | 20,619,836 (32.7%) | 21,261,363 (42.3%) | 17,240,892 (34.3%) | 1,824,939 (3.6%) | 9,897,079 (19.7%) | 5,774,535 (25.7%) | 14,461,147 (64.3%) | 307,839 (1.4%) | 1,956,339 (8.7%) |
| **12-Dec-21** | 19,265,731 (30.5%) | 19,773,941 (31.3%) | 3,566,118 (5.7%) | 20,494,204 (32.5%) | 18,864,315 (37.6%) | 19,757,260 (39.3%) | 1,774,889 (3.5%) | 9,827,809 (19.6%) | 4,272,802 (19.0%) | 15,982,960 (71.0%) | 298,816 (1.3%) | 1,945,282 (8.6%) |
| **19-Dec-21** | 14,761,022 (23.4%) | 24,518,704 (38.9%) | 3,513,151 (5.6%) | 20,307,117 (32.2%) | 14,283,111 (28.4%) | 24,492,870 (48.8%) | 1,735,265 (3.5%) | 9,713,027 (19.3%) | 2,894,226 (12.9%) | 17,384,401 (77.3%) | 293,512 (1.3%) | 1,927,721 (8.6%) |
| **26-Dec-21** | 12,144,709 (19.2%) | 27,318,277 (43.3%) | 3,467,929 (5.5%) | 20,169,079 (32.0%) | 11,601,910 (23.1%) | 27,283,932 (54.3%) | 1,712,801 (3.4%) | 9,625,630 (19.2%) | 2,301,915 (10.2%) | 17,991,957 (80.0%) | 290,631 (1.3%) | 1,915,357 (8.5%) |
| **2-Jan-22** | 10,944,969 (17.3%) | 28,675,140 (45.4%) | 3,432,361 (5.4%) | 20,047,524 (31.8%) | 10,325,499 (20.6%) | 28,634,734 (57.0%) | 1,706,797 (3.4%) | 9,557,243 (19.0%) | 2,051,827 (9.1%) | 18,250,879 (81.1%) | 290,032 (1.3%) | 1,907,122 (8.5%) |
| **9-Jan-22** | 9,977,945 (15.8%) | 29,846,067 (47.3%) | 3,371,664 (5.3%) | 19,904,318 (31.5%) | 9,255,079 (18.4%) | 29,798,032 (59.3%) | 1,691,094 (3.4%) | 9,480,068 (18.9%) | 1,844,119 (8.2%) | 18,468,986 (82.1%) | 288,108 (1.3%) | 1,898,647 (8.4%) |

| | | | | | | | | | | | | |
|---|---|---|---|---|---|---|---|---|---|---|---|---|
| **16-Jan-22** | 9,546,563 (15.1%) | 30,459,199 (48.3%) | 3,307,547 (5.2%) | 19,786,685 (31.4%) | 8,725,007 (17.4%) | 30,401,739 (60.5%) | 1,684,402 (3.4%) | 9,413,125 (18.7%) | 1,739,053 (7.7%) | 18,582,419 (82.6%) | 286,899 (1.3%) | 1,891,489 (8.4%) |
| **23-Jan-22** | 9,390,117 (14.9%) | 30,805,729 (48.8%) | 3,217,300 (5.1%) | 19,686,848 (31.2%) | 8,454,911 (16.8%) | 30,734,485 (61.2%) | 1,677,190 (3.3%) | 9,357,687 (18.6%) | 1,685,346 (7.5%) | 18,642,856 (82.9%) | 285,949 (1.3%) | 1,885,709 (8.4%) |
| **30-Jan-22** | 9,296,619 (14.7%) | 31,087,436 (49.3%) | 3,117,686 (4.9%) | 19,598,253 (31.1%) | 8,243,011 (16.4%) | 31,004,983 (61.7%) | 1,668,538 (3.3%) | 9,307,741 (18.5%) | 1,639,123 (7.3%) | 18,695,457 (83.1%) | 284,923 (1.3%) | 1,880,357 (8.4%) |
| **6-Feb-22** | 9,262,440 (14.7%) | 31,274,875 (49.6%) | 3,032,392 (4.8%) | 19,530,287 (31.0%) | 8,110,384 (16.1%) | 31,184,760 (62.1%) | 1,654,914 (3.3%) | 9,274,215 (18.5%) | 1,611,069 (7.2%) | 18,728,231 (83.2%) | 283,227 (1.3%) | 1,877,333 (8.3%) |
| **13-Feb-22** | 9,241,569 (14.6%) | 31,442,367 (49.8%) | 2,950,014 (4.7%) | 19,466,044 (30.8%) | 7,995,490 (15.9%) | 31,345,394 (62.4%) | 1,634,487 (3.3%) | 9,248,902 (18.4%) | 1,589,530 (7.1%) | 18,754,378 (83.4%) | 280,876 (1.2%) | 1,875,076 (8.3%) |
| **20-Feb-22** | 9,209,801 (14.6%) | 31,581,864 (50.1%) | 2,892,982 (4.6%) | 19,415,347 (30.8%) | 7,902,747 (15.7%) | 31,476,584 (62.7%) | 1,615,652 (3.2%) | 9,229,290 (18.4%) | 1,572,678 (7.0%) | 18,775,259 (83.4%) | 278,643 (1.2%) | 1,873,280 (8.3%) |
| **27-Feb-22** | 9,175,261 (14.5%) | 31,712,912 (50.3%) | 2,842,449 (4.5%) | 19,369,372 (30.7%) | 7,824,511 (15.6%) | 31,592,299 (62.9%) | 1,598,003 (3.2%) | 9,209,460 (18.3%) | 1,556,155 (6.9%) | 18,795,475 (83.5%) | 276,809 (1.2%) | 1,871,421 (8.3%) |
| **6-Mar-22** | 9,156,404 (14.5%) | 31,823,142 (50.4%) | 2,787,551 (4.4%) | 19,332,897 (30.6%) | 7,767,075 (15.5%) | 31,683,458 (63.1%) | 1,580,454 (3.1%) | 9,193,286 (18.3%) | 1,543,289 (6.9%) | 18,811,620 (83.6%) | 274,926 (1.2%) | 1,870,025 (8.3%) |
| **13-Mar-22** | 9,141,789 (14.5%) | 31,926,564 (50.6%) | 2,731,039 (4.3%) | 19,300,602 (30.6%) | 7,717,387 (15.4%) | 31,766,822 (63.2%) | 1,561,398 (3.1%) | 9,178,666 (18.3%) | 1,532,248 (6.8%) | 18,825,918 (83.7%) | 272,981 (1.2%) | 1,868,713 (8.3%) |
| **20-Mar-22** | 9,131,054 (14.5%) | 32,024,760 (50.8%) | 2,672,836 (4.2%) | 19,271,344 (30.5%) | 7,672,917 (15.3%) | 31,844,165 (63.4%) | 1,541,996 (3.1%) | 9,165,195 (18.2%) | 1,522,702 (6.8%) | 18,838,824 (83.7%) | 270,793 (1.2%) | 1,867,541 (8.3%) |
| **27-Mar-22** | 9,117,101 (14.4%) | 32,115,500 (50.9%) | 2,622,602 (4.2%) | 19,244,791 (30.5%) | 7,629,664 (15.2%) | 31,915,631 (63.5%) | 1,525,760 (3.0%) | 9,153,218 (18.2%) | 1,511,149 (6.7%) | 18,853,412 (83.8%) | 269,098 (1.2%) | 1,866,201 (8.3%) |

**Table S3a: NIMS database with vaccination status of all age groups , over 18 and 50 years of age.** The weekly number of population in each vaccination group and percentage represented for each week based on the age. The total NIMS cohort of all age groups (n=63,099,994); over 18 years of age (n=50,224,273) and over 50 years of age (n=22,499,860) as of March 27, 2022. For the entire population (all ages), the NIMS database have 8.9% to 9.0% more percentage of unvaccinated population than the England database as shown on Table S3c in between December 19, 2021 to March 27, 2022.

#### Table S3b: Vaccination status of racial ethnic minorities and vaccination status based on IMD score of the over 18 years of age

| 18+ years of age | | Vaccination status; n=population (%) | | | |
|---|---|---|---|---|---|
| **Ethnicity status** | **Denominator** | **1 dose only** | **2 doses only** | **Third dose** | **Unvaccinated** |
| **White - British** | 30,755,212 | 654,705 (2.1%) | 3,847,492 (12.5%) | 23,506,318 (76.4%) | 2,746,697 (8.9%) |
| **White - Irish** | 305,107 | 6,836 (2.2%) | 32,959 (10.8%) | 212,706 (69.7%) | 52,606 (17.2%) |
| **White - Other** | 4,364,388 | 117,373 (2.7%) | 758,638 (17.4%) | 2,011,587 (46.1%) | 1,476,790 (33.8%) |
| Mixed - White and Black Caribbean | 163,185 | 7,854 (4.8%) | 34,452 (21.1%) | 58,500 (35.8%) | 62,379 (38.2%) |
| Mixed - White and Black African | 125,836 | 6,193 (4.9%) | 32,845 (26.1%) | 50,148 (39.9%) | 36,650 (29.1%) |
| Mixed - White and Asian | 141,390 | 5,274 (3.7%) | 26,766 (18.9%) | 78,564 (55.6%) | 30,786 (21.8%) |
| Mixed - Any other mixed background | 307,921 | 12,813 (4.2%) | 60,728 (19.7%) | 148,242 (48.1%) | 86,138 (28.0%) |
| Asian or Asian British - Indian | 1,459,219 | 46,807 (3.2%) | 268,328 (18.4%) | 896,598 (61.4%) | 247,486 (17.0%) |
| Asian or Asian British - Pakistani | 1,047,413 | 60,910 (5.8%) | 387,666 (37.0%) | 366,405 (35.0%) | 232,432 (22.2%) |
| Asian or Asian British - Bangladeshi | 379,935 | 18,740 (4.9%) | 117,464 (30.9%) | 176,429 (46.4%) | 67,302 (17.7%) |
| Asian or Asian British - Any other Asian background | 885,577 | 36,255 (4.1%) | 192,281 (21.7%) | 483,804 (54.6%) | 173,237 (19.6%) |
| Black or Black British - Caribbean | 413,011 | 16,529 (4.0%) | 79,798 (19.3%) | 145,374 (35.2%) | 171,310 (41.5%) |
| Black or Black British - African | 911,671 | 50,450 (5.5%) | 264,944 (29.1%) | 317,774 (34.9%) | 278,503 (30.5%) |
| Black or Black British - Any other Black background | 346,270 | 18,713 (5.4%) | 88,468 (25.5%) | 108,548 (31.3%) | 130,541 (37.7%) |
| Chinese | 451,559 | 24,059 (5.3%) | 52,763 (11.7%) | 203,476 (45.1%) | 171,261 (37.9%) |
| Other ethnic groups - Any other ethnic group | 1,122,853 | 46,019 (4.1%) | 238,690 (21.3%) | 475,598 (42.4%) | 362,546 (32.3%) |
| Not Stated/Unknown | 6,461,320 | 323,991 (5.0%) | 949,393 (14.7%) | 2,956,131 (45.8%) | 2,231,805 (34.5%) |
| **Non-white ethnic groups (all)** | 12,425,335 | 350,616 (4.5%) | 1,845,193 (23.8%) | 3,509,460 (45.2%) | 2,050,571 (26.4%) |

| 18+ years of Age | Vaccination status; n=population (%) | | | |
|---|---|---|---|---|
| **IMD decile** | **1 dose only** | **2 doses only** | **Third dose** | **Unvaccinated** |
| **1 (most deprived)** | 231,099 (5.6%) | 1,036,060 (25.2%) | 2,266,843 (55.2%) | 13.9% |
| **2** | 206,422 (4.8%) | 1,000,801 (23.2%) | 2,575,207 (59.7%) | 12.3% |
| **3** | 192,948 (4.3%) | 954,499 (21.1%) | 2,853,084 (63.0%) | 11.6% |

| | | | | |
|---|---|---|---|---|
| **4** | 172,467 (3.7%) | 853,092 (18.5%) | 3,117,666 (67.7%) | 10.0% |
| **5** | 144,138 (3.2%) | 750,866 (16.4%) | 3,289,305 (72.0%) | 8.4% |
| **6** | 130,418 (2.8%) | 688,136 (14.9%) | 3,491,214 (75.4%) | 6.9% |
| **7** | 110,690 (2.5%) | 610,577 (13.6%) | 3,539,976 (78.6%) | 5.3% |
| **8** | 103,928 (2.3%) | 582,920 (13.0%) | 3,637,946 (81.1%) | 3.6% |
| **9** | 86,911 (2.0%) | 522,274 (11.9%) | 3,672,750 (83.4%) | 2.8% |
| **10 (least deprived)** | 74,766 (1.7%) | 440,433 (10.2%) | 3,744,140 (86.7%) | 1.4% |

**Table S3b: Vaccination status of racial ethnic minorities and vaccination status based on IMD score of the over 18 years of age NIMS population as of May 1, 2022.**

**Table S3c:** **Vaccination status of all ages (entire population) of England based on England vaccination database.**

| Week ending | England vaccination database-All ages, Vaccination status; n=population (%) | | | |
|---|---|---|---|---|
| | Two doses (no 3rd dose) | Third dose | 1 dose only | Unvaccinated |
| **1/10/2021** | 374,613 (0.7%) | | 1,584,538 (2.8%) | 54,590,987 (96.5%) |
| **1/17/2021** | 427,386 (0.8%) | | 3,092,670 (5.5%) | 53,030,082 (93.8%) |
| **1/24/2021** | 441,684 (0.8%) | | 5,286,009 (9.3%) | 50,822,445 (89.9%) |
| **1/31/2021** | 460,907 (0.8%) | | 7,621,448 (13.5%) | 48,467,783 (85.7%) |
| **2/7/2021** | 471,636 (0.8%) | | 10,048,093 (17.8%) | 46,030,409 (81.4%) |
| **2/14/2021** | 490,722 (0.9%) | | 12,372,187 (21.9%) | 43,687,229 (77.3%) |
| **2/21/2021** | 513,435 (0.9%) | | 14,444,639 (25.5%) | 41,592,064 (73.5%) |
| **2/28/2021** | 599,935 (1.1%) | | 16,612,869 (29.4%) | 39,337,334 (69.6%) |
| **3/7/2021** | 797,321 (1.4%) | | 18,218,176 (32.2%) | 37,534,641 (66.4%) |
| **3/14/2021** | 1,129,444 (2.0%) | | 19,662,394 (34.8%) | 35,758,300 (63.2%) |
| **3/21/2021** | 1,621,547 (2.9%) | | 22,233,315 (39.3%) | 32,695,276 (57.8%) |
| **3/28/2021** | 2,806,124 (5.0%) | | 23,097,658 (40.8%) | 30,646,356 (54.2%) |
| **4/4/2021** | 4,344,251 (7.7%) | | 22,401,788 (39.6%) | 29,804,099 (52.7%) |
| **4/11/2021** | 6,338,332 (11.2%) | | 20,769,258 (36.7%) | 29,442,548 (52.1%) |
| **4/18/2021** | 8,518,498 (15.1%) | | 19,110,081 (33.8%) | 28,921,559 (51.1%) |
| **4/25/2021** | 10,791,851 (19.1%) | | 17,497,445 (30.9%) | 28,260,842 (50.0%) |
| **5/2/2021** | 12,972,758 (22.9%) | | 15,993,178 (28.3%) | 27,584,202 (48.8%) |
| **5/9/2021** | 15,031,521 (26.6%) | | 14,620,033 (25.9%) | 26,898,584 (47.6%) |
| **5/16/2021** | 17,172,209 (30.4%) | | 13,471,265 (23.8%) | 25,906,664 (45.8%) |
| **5/23/2021** | 19,427,631 (34.4%) | | 12,399,174 (21.9%) | 24,723,333 (43.7%) |
| **5/30/2021** | 21,719,461 (38.4%) | | 11,219,035 (19.8%) | 23,611,642 (41.8%) |
| **6/6/2021** | 23,710,646 (41.9%) | | 10,089,461 (17.8%) | 22,750,031 (40.2%) |
| **6/13/2021** | 25,391,916 (44.9%) | | 9,459,223 (16.7%) | 21,698,999 (38.4%) |
| **6/20/2021** | 26,534,936 (46.9%) | | 9,566,842 (16.9%) | 20,448,360 (36.2%) |

| | | | | |
|---|---|---|---|---|
| **6/27/2021** | 27,414,725 (48.5%) | | 9,861,169 (17.4%) | 19,274,244 (34.1%) |
| **7/4/2021** | 28,324,385 (50.1%) | | 9,719,772 (17.2%) | 18,505,981 (32.7%) |
| **7/11/2021** | 29,204,296 (51.6%) | | 9,324,907 (16.5%) | 18,020,935 (31.9%) |
| **7/18/2021** | 30,213,335 (53.4%) | | 8,650,475 (15.3%) | 17,686,328 (31.3%) |
| **7/25/2021** | 31,181,649 (55.1%) | | 7,922,798 (14.0%) | 17,445,691 (30.8%) |
| **8/1/2021** | 32,110,377 (56.8%) | | 7,198,835 (12.7%) | 17,240,926 (30.5%) |
| **8/8/2021** | 33,004,783 (58.4%) | | 6,490,321 (11.5%) | 17,055,034 (30.2%) |
| **8/15/2021** | 33,971,458 (60.1%) | | 5,737,867 (10.1%) | 16,840,813 (29.8%) |
| **8/22/2021** | 34,927,576 (61.8%) | | 5,076,721 (9.0%) | 16,545,841 (29.3%) |
| **8/29/2021** | 35,715,918 (63.2%) | | 4,572,956 (8.1%) | 16,261,264 (28.8%) |
| **9/5/2021** | 36,352,855 (64.3%) | | 4,137,264 (7.3%) | 16,060,019 (28.4%) |
| **9/12/2021** | 36,865,490 (65.2%) | | 3,765,715 (6.7%) | 15,918,933 (28.2%) |
| **9/19/2021** | 37,244,704 (65.9%) | | 3,508,137 (6.2%) | 15,797,297 (27.9%) |
| **9/26/2021** | 37,510,886 (66.3%) | | 3,362,885 (5.9%) | 15,676,367 (27.7%) |
| **10/3/2021** | 36,409,103 (64.4%) | 1,298,612 (2.3%) | 3,332,894 (5.9%) | 15,509,529 (27.4%) |
| **10/10/2021** | 35,449,302 (62.7%) | 2,426,155 (4.3%) | 3,334,052 (5.9%) | 15,340,629 (27.1%) |
| **10/17/2021** | 34,340,945 (60.7%) | 3,695,991 (6.5%) | 3,362,399 (5.9%) | 15,150,803 (26.8%) |
| **10/24/2021** | 32,950,633 (58.3%) | 5,235,928 (9.3%) | 3,469,279 (6.1%) | 14,894,298 (26.3%) |
| **10/31/2021** | 31,489,596 (55.7%) | 6,826,159 (12.1%) | 3,577,930 (6.3%) | 14,656,453 (25.9%) |
| **11/7/2021** | 29,820,842 (52.7%) | 8,616,120 (15.2%) | 3,714,085 (6.6%) | 14,399,091 (25.5%) |
| **11/14/2021** | 27,813,943 (49.2%) | 10,776,175 (19.1%) | 3,858,390 (6.8%) | 14,101,630 (24.9%) |
| **11/21/2021** | 25,879,170 (45.8%) | 12,830,865 (22.7%) | 3,907,598 (6.9%) | 13,932,505 (24.6%) |
| **11/28/2021** | 23,863,068 (42.2%) | 14,995,780 (26.5%) | 3,901,108 (6.9%) | 13,790,182 (24.4%) |
| **12/5/2021** | 21,803,895 (38.6%) | 17,242,093 (30.5%) | 3,867,342 (6.8%) | 13,636,808 (24.1%) |
| **12/12/2021** | 19,487,030 (34.5%) | 19,745,991 (34.9%) | 3,818,248 (6.8%) | 13,498,869 (23.9%) |
| **12/19/2021** | 15,100,851 (26.7%) | 24,376,660 (43.1%) | 3,772,998 (6.7%) | 13,299,629 (23.5%) |
| **12/26/2021** | 12,361,345 (21.9%) | 27,312,396 (48.3%) | 3,728,022 (6.6%) | 13,148,375 (23.3%) |
| **1/2/2022** | 11,129,075 (19.7%) | 28,706,633 (50.8%) | 3,692,190 (6.5%) | 13,022,240 (23.0%) |
| **1/9/2022** | 10,145,018 (17.9%) | 29,897,180 (52.9%) | 3,636,558 (6.4%) | 12,871,382 (22.8%) |
| **1/16/2022** | 9,700,362 (17.2%) | 30,526,563 (54.0%) | 3,578,827 (6.3%) | 12,744,386 (22.5%) |
| **1/23/2022** | 9,531,469 (16.9%) | 30,888,097 (54.6%) | 3,491,555 (6.2%) | 12,639,017 (22.4%) |

| | | | | |
|---|---|---|---|---|
| **1/30/2022** | 9,428,375 (16.7%) | 31,180,755 (55.1%) | 3,398,776 (6.0%) | 12,542,232 (22.2%) |
| **2/6/2022** | 9,390,711 (16.6%) | 31,378,540 (55.5%) | 3,314,512 (5.9%) | 12,466,375 (22.0%) |
| **2/13/2022** | 9,365,359 (16.6%) | 31,553,940 (55.8%) | 3,232,663 (5.7%) | 12,398,176 (21.9%) |
| **2/20/2022** | 9,329,464 (16.5%) | 31,702,092 (56.1%) | 3,172,802 (5.6%) | 12,345,780 (21.8%) |
| **2/27/2022** | 9,294,209 (16.4%) | 31,839,752 (56.3%) | 3,118,611 (5.5%) | 12,297,566 (21.7%) |
| **3/6/2022** | 9,263,733 (16.4%) | 31,966,231 (56.5%) | 3,061,901 (5.4%) | 12,258,273 (21.7%) |
| **3/13/2022** | 9,245,305 (16.3%) | 32,076,944 (56.7%) | 3,001,131 (5.3%) | 12,226,758 (21.6%) |
| **3/20/2022** | 9,228,619 (16.3%) | 32,184,060 (56.9%) | 2,936,877 (5.2%) | 12,200,582 (21.6%) |
| **3/27/2022** | 9,212,937 (16.3%) | 32,279,253 (57.1%) | 2,882,600 (5.1%) | 12,175,348 (21.5%) |

**Table S3c: England vaccination database (UK corona virus dashboard) with vaccination status of all age groups in England.** The weekly number of population in each vaccination group and percentage represented for each week based on the age. The total England cohort of all age groups (n=56,550,138) derived from the England mid 2020 census estimate as shown on Table S4. Age subgroup information is not available for age groups ≥18 yrs or ≥50 yrs of age. For the entire population (all ages), the England database have 8.9% to 9.0% less percentage of unvaccinated population than the NIMS database as shown on Table S3a in between December 19, 2021 to March 27, 2022.

### Table S4: Population of United Kingdom based on mid 2020 census estimate

| | United Kingdom | England and Wales | England |
|---|---|---|---|
| **Total population** | 67,081,234 | 59,719,724 | 56,550,138 |
| ***Age group*** | | | |
| **0-19 yrs** | 15,658,537 (23.3%) | 14,032,132 (23.5%) | 13,330,355 (23.6%) |
| **20-29 yrs** | 8,609,788 (12.8%) | 7,658,686 (12.8%) | 7,244,015 (12.8%) |
| **30-49 yrs** | 17,321,585 (25.8%) | 15,425,743 (25.8%) | 14,677,803 (26.0%) |
| **50-69 yrs** | 16,338,067 (24.4%) | 14,434,809 (24.2%) | 13,618,246 (24.1%) |
| **≥50 yrs** | 25,491,324 (38.0%) | 22,603,163 (37.8%) | 21,297,965 (37.7%) |
| **≥70 yrs** | 9,153,257 (13.6%) | 8,168,354 (13.7%) | 7,679,719 (13.6%) |
| **≥75 yrs** | 5,789,351 (8.6%) | 5,172,340 (8.7%) | 4,865,591 (8.6%) |
| **≥80 yrs** | 3,385,592 (5.0%) | 3,031,325 (5.1%) | 2,855,599 (5.0%) |

**Table S4: Population of United Kingdom based on mid 2020 census estimate**. Age group distribution of population in UK, England; England and Wales.

#### Table S5: Outcomes of Confirmed Delta variant cases, Hospitalizations and deaths

| Study period | Age (yrs) | Outcome | **Total** (n= % of all ages) | **Unvaccinated** (n=% of age group) | **Vaccinated** (n=% of age group) | | | **Vaccination status unknown** (n=% of age group) |
|---|---|---|---|---|---|---|---|---|
| | | | | | All Vaccinated | 1 dose* | Two doses‡ | |
| **Feb 1- June 20, 2021** | **All ages** | **Cases** | 123,620 (100%) | 71,932 (58.2%) | 37,329 (30.2%) | 26,495 (21.4%) | 10,834 (8.8%) | 14,359 (11.6%) |
| | | **Hospitalizations** | 1,904 (100%) | 1,182 (62.1%) | 688 (36.1%) | 375 (19.7%) | 313 (16.4%) | 34 (1.8%) |
| | | **Deaths** | 257 (100%) | 92 (35.8%) | 163 (63.4%) | 45 (17.5%) | 118 (45.9%) | 2 (0.8%) |
| | **<50 yrs** | **Cases** | 111,008 (89.8%) | 70,664 (63.7%) | 27,444 (24.7%) | 21,844 (19.7%) | 5,600 (5.0%) | 12,900 (11.6%) |
| | | **Hospitalizations** | 1,283 (67.4%) | 987 (76.9%) | 272 (21.2%) | 224 (17.5%) | 48 (3.7%) | 24 (1.9%) |
| | | **Deaths** | 26 (10.1%) | 21 (80.8%) | 5 (19.2%) | 3 (11.5%) | 2 (7.7%) | 0 (0.0%) |
| | **≥50 yrs** | **Cases** | 12,404 (10.0%) | 1,267 (10.2%) | 9,885 (79.7%) | 4,651 (37.5%) | 5,234 (42.2%) | 1,252 (10.1%) |
| | | **Hospitalizations** | 615 (32.3%) | 195 (31.7%) | 416 (67.6%) | 151 (24.6%) | 265 (43.1%) | 4 (0.7%) |
| | | **Deaths** | 231 (89.9%) | 71 (30.7%) | 158 (68.4%) | 42 (18.2%) | 116 (50.2%) | 2 (0.9%) |
| **June 21-July 18, 2021** | **All ages** | **Cases** | 105,598 (100%) | 49,470 (46.8%) | 45,535 (43.1%) | 27,596 (26.1%) | 17,939 (17.0%) | 10,593 (10.0%) |
| | | **Hospitalizations** | 1,788 (100%) | 970 (54.3%) | 788 (44.1%) | 258 (14.4%) | 530 (29.6%) | 30 (1.7%) |
| | | **Deaths** | 203 (100%) | 73 (36.0%) | 126 (62.1%) | 20 (9.9%) | 106 (52.2%) | 4 (2.0%) |
| | **<50 yrs** | **Cases** | 94,541 (89.5%) | 48,399 (51.2%) | 36,546 (38.7%) | 26,800 (28.3%) | 9,746 (10.3%) | 9,586 (10.1%) |
| | | **Hospitalizations** | 1,044 (58.4%) | 725 (69.4%) | 292 (28.0%) | 200 (19.2%) | 92 (8.8%) | 27 (2.6%) |
| | | **Deaths** | 19 (9.4%) | 13 (68.4%) | 5 (26.3%) | 3 (15.8%) | 2 (10.5%) | 1 (5.3%) |
| | **≥50 yrs** | **Cases** | 10,975 (10.4%) | 1,070 (9.7%) | 8,988 (81.9%) | 795 (7.2%) | 8,193 (74.7%) | 917 (8.4%) |
| | | **Hospitalizations** | 750 (41.9%) | 245 (32.7%) | 496 (66.1%) | 58 (7.7%) | 438 (58.4%) | 9 (1.2%) |
| | | **Deaths** | 184 (90.6%) | 60 (32.6%) | 121 (65.8%) | 17 (9.2%) | 104 (56.5%) | 3 (1.6%) |
| **July 19-Aug 1, 2021** | **All ages** | **Cases** | 70,792 (100%) | 29,652 (41.9%) | 34,251 (48.4%) | 16,016 (22.6%) | 18,235 (25.8%) | 6,889 (9.7%) |
| | | **Hospitalizations** | 1,467 (100%) | 808 (55.1%) | 641 (43.7%) | 129 (8.8%) | 512 (34.9%) | 18 (1.2%) |
| | | **Deaths** | 282 (100%) | 88 (31.2%) | 192 (68.1%) | 14 (5.0%) | 178 (63.1%) | 2 (0.7%) |
| | **<50 yrs** | **Cases** | 60,200 (85.0%) | 28,549 (47.4%) | 25,817 (42.9%) | 15,627 (26.0%) | 10,190 (16.9%) | 5,834 (9.7%) |

| | | | | | | | | |
|---|---|---|---|---|---|---|---|---|
| | | **Hospitalizations** | 757 (51.6%) | 578 (76.4%) | 169 (22.3%) | 85 (11.2%) | 84 (11.1%) | 10 (1.3%) |
| | | **Deaths** | 26 (9.2%) | 14 (53.8%) | 11 (42.3%) | 2 (7.7%) | 9 (34.6%) | 1 (3.8%) |
| | **≥50 yrs** | **Cases** | 10,357 (14.6%) | 1,103 (10.6%) | 8,434 (81.4%) | 389 (3.8%) | 8,045 (77.7%) | 820 (7.9%) |
| | | **Hospitalizations** | 709 (48.3%) | 230 (32.4%) | 472 (66.6%) | 44 (6.2%) | 428 (60.4%) | 7 (1.0%) |
| | | **Deaths** | 255 (90.4%) | 74 (29.0%) | 181 (71.0%) | 12 (4.7%) | 169 (66.3%) | - (0.0%) |
| **Aug 2-Aug 15, 2021** | **All ages** | **Cases** | 86,725 (100%) | 32,079 (37.0%) | 46,214 (53.3%) | 19,850 (22.9%) | 26,364 (30.4%) | 8,432 (9.7%) |
| | | **Hospitalizations** | 2,126 (100%) | 1,073 (50.5%) | 1,036 (48.7%) | 187 (8.8%) | 849 (39.9%) | 17 (0.8%) |
| | | **Deaths** | 447 (100%) | 137 (30.6%) | 302 (67.6%) | 25 (5.6%) | 277 (62.0%) | 8 (1.8%) |
| | **<50 yrs** | **Cases** | 72,085 (83.1%) | 30,628 (42.5%) | 34,390 (47.7%) | 19,382 (26.9%) | 15,008 (20.8%) | 7,067 (9.8%) |
| | | **Hospitalizations** | 1,028 (48.4%) | 754 (73.3%) | 264 (25.7%) | 122 (11.9%) | 142 (13.8%) | 10 (1.0%) |
| | | **Deaths** | 42 (9.4%) | 24 (57.1%) | 17 (40.5%) | 3 (7.1%) | 14 (33.3%) | 1 (2.4%) |
| | **≥50 yrs** | **Cases** | 14,528 (16.8%) | 1,451 (10.0%) | 11,824 (81.4%) | 468 (3.2%) | 11,356 (78.2%) | 1,253 (8.6%) |
| | | **Hospitalizations** | 1,099 (51.7%) | 319 (29.0%) | 772 (70.2%) | 65 (5.9%) | 707 (64.3%) | 8 (0.7%) |
| | | **Deaths** | 406 (90.8%) | 113 (27.8%) | 285 (70.2%) | 22 (5.4%) | 263 (64.8%) | 8 (2.0%) |
| **Aug 16-Aug 29, 2021** | **All ages** | **Cases** | 105,793 (100%) | 36,583 (34.6%) | 59,364 (56.1%) | 18,913 (17.9%) | 40,451 (38.2%) | 9,846 (9.3%) |
| | | **Hospitalizations** | 2,187 (100%) | 1,031 (47.1%) | 1,115 (51.0%) | 147 (6.7%) | 968 (44.3%) | 41 (1.9%) |
| | | **Deaths** | 609 (100%) | 146 (24.0%) | 450 (73.9%) | 38 (6.2%) | 412 (67.7%) | 13 (2.1%) |
| | **<50 yrs** | **Cases** | 82,855 (78.3%) | 34,749 (41.9%) | 40,176 (48.5%) | 18,317 (22.1%) | 21,859 (26.4%) | 7,930 (9.6%) |
| | | **Hospitalizations** | 986 (45.1%) | 698 (70.8%) | 248 (25.2%) | 93 (9.4%) | 155 (15.7%) | 40 (4.1%) |
| | | **Deaths** | 41 (6.7%) | 27 (65.9%) | 13 (31.7%) | 3 (7.3%) | 10 (24.4%) | 1 (2.4%) |
| | **≥50 yrs** | **Cases** | 22,843 (21.6%) | 1,833 (8.0%) | 19,188 (84.0%) | 596 (2.6%) | 18,592 (81.4%) | 1,822 (8.0%) |
| | | **Hospitalizations** | 1,201 (54.9%) | 333 (27.7%) | 867 (72.2%) | 54 (4.5%) | 813 (67.7%) | 1 (0.1%) |
| | | **Deaths** | 568 (93.3%) | 119 (21.0%) | 437 (76.9%) | 35 (6.2%) | 402 (70.8%) | 12 (2.1%) |
| **Aug 30-Sept 12, 2021** | **All ages** | **Cases** | 101,044 (100%) | 37,641 (37.3%) | 55,519 (54.9%) | 11,942 (11.8%) | 43,577 (43.1%) | 7,884 (7.8%) |
| | | **Hospitalizations** | 2,935 (100%) | 1,239 (42.2%) | 1,649 (56.2%) | 187 (6.4%) | 1,462 (49.8%) | 47 (1.6%) |
| | | **Deaths** | 744 (100%) | 186 (25.0%) | 546 (73.4%) | 24 (3.2%) | 522 (70.2%) | 12 (1.6%) |
| | **<50 yrs** | **Cases** | 76,416 (75.6%) | 35,814 (46.9%) | 34,402 (45.0%) | 11,398 (14.9%) | 23,004 (30.1%) | 6,200 (8.1%) |
| | | **Hospitalizations** | 1,132 (38.6%) | 775 (68.5%) | 324 (28.6%) | 124 (11.0%) | 200 (17.7%) | 33 (2.9%) |

| | | | | | | | | |
|---|---|---|---|---|---|---|---|---|
| | | **Deaths** | 50 (6.7%) | 33 (66.0%) | 14 (28.0%) | 3 (6.0%) | 11 (22.0%) | 3 (6.0%) |
| | **≥50 yrs** | **Cases** | 24,480 (24.2%) | 1,827 (7.5%) | 21,115 (86.3%) | 544 (2.2%) | 20,571 (84.0%) | 1,538 (6.3%) |
| | | **Hospitalizations** | 1,793 (61.1%) | 464 (25.9%) | 1,325 (73.9%) | 63 (3.5%) | 1,262 (70.4%) | 4 (0.2%) |
| | | **Deaths** | 692 (93.0%) | 153 (22.1%) | 532 (76.9%) | 21 (3.0%) | 511 (73.8%) | 7 (1.0%) |

**Table S5: Outcomes of Confirmed Delta variant cases, Hospitalizations and deaths based on the vaccination status reported in Public Health England technical briefings until September 12, 2021**. *SARS-CoV2 cases tested positive with the specimen date post receiving the first dose without additional doses of vaccination are considered one dose. ‡SARS-CoV2 cases tested positive with the specimen date ≥14 days post second dose **are** considered vaccinated with two doses. *The age specified denominator (population) for the ≥50 years of age group based on the vaccination status (1 dose, two doses and unvaccinated), for each specified period ending date was shown on the Table S3a.*

**Table S6a**: **SARS-CoV2 cases reported by specimen date (reported weekly for prior 4 weeks period) in NIMS-database based on vaccination status**

| Outcome | Study period (rolling four weeks) | Age (yrs) | Total cases (n=% of cases all ages) | Unvaccinated Cases (n=% for age group) | Vaccinated cases (cases) (n=% for age group) | | | | Vaccination status unknown (n=% for age group) |
|---|---|---|---|---|---|---|---|---|---|
| | | | | | All vaccinated | 1 dose* | Two doses‡ | Third dose ø | |
| Cases | Aug 16-Sept 12, 2021 | All ages | 711,304 (100%) | 711,304 (37.0%) | 382,422 (53.8%) | 93,143 (13.1%) | 289,279 (40.7%) | | 65,597 (9.2%) |
| | | ≥18 | 520,441 (73.2%) | 101,867 (19.6%) | 369,802 (71.1%) | 81,332 (15.6%) | 288,470 (55.4%) | | 48,772 (9.4%) |
| | | ≥50 | 170,525 (24.0%) | 11,265 (6.6%) | 146,085 (85.7%) | 3,575 (2.1%) | 142,510 (83.6%) | | 13,175 (7.7%) |
| Cases | Aug 23-Sept 19, 2021 | All ages | 680,582 (100%) | 680,582 (40.5%) | 344,691 (50.6%) | 67,187 (9.9%) | 277,504 (40.8%) | | 60,046 (8.8%) |
| | | ≥18 | 461,551 (67.8%) | 87,377 (18.9%) | 331,990 (71.9%) | 55,159 (12.0%) | 276,831 (60.0%) | | 42,184 (9.1%) |
| | | ≥50 | 160,432 (23.6%) | 10,247 (6.4%) | 138,185 (86.1%) | 3,129 (2.0%) | 135,056 (84.2%) | | 12,000 (7.5%) |
| Cases | Aug 30-Sept 26, 2021 | All ages | 699,489 (100%) | 699,489 (45.2%) | 326,250 (46.6%) | 53,070 (7.6%) | 273,180 (39.1%) | | 57,237 (8.2%) |
| | | ≥18 | 425,804 (60.9%) | 75,925 (17.8%) | 312,932 (73.5%) | 40,434 (9.5%) | 272,498 (64.0%) | | 36,947 (8.7%) |
| | | ≥50 | 149,993 (21.4%) | 9,131 (6.1%) | 130,176 (86.8%) | 2,790 (1.9%) | 127,386 (84.9%) | | 10,686 (7.1%) |
| Cases | Sept 6-October 3, 2021 | All ages | 696,281 (100%) | 696,281 (48.5%) | 305,326 (43.9%) | 39,232 (5.6%) | 266,094 (38.2%) | | 53,385 (7.7%) |
| | | ≥18 | 390,853 (56.1%) | 64,589 (16.5%) | 293,846 (75.2%) | 28,361 (7.3%) | 265,485 (67.9%) | | 32,418 (8.3%) |
| | | ≥50 | 140,407 (20.2%) | 8,038 (5.7%) | 122,880 (87.5%) | 2,541 (1.8%) | 120,339 (85.7%) | | 9,489 (6.8%) |
| Cases | Sept 13-Oct 10, 2021 | All ages | 750,518 (100%) | 750,518 (49.5%) | 325,759 (43.4%) | 37,578 (5.0%) | 288,181 (38.4%) | | 53,063 (7.1%) |
| | | ≥18 | 402,004 (53.6%) | 60,497 (15.0%) | 310,745 (77.3%) | 23,218 (5.8%) | 287,527 (71.5%) | | 30,762 (7.7%) |
| | | ≥50 | 147,044 (19.6%) | 7,689 (5.2%) | 130,192 (88.5%) | 2,496 (1.7%) | 127,696 (86.8%) | | 9,163 (6.2%) |
| Cases | | All ages | 856,370 (100%) | 856,370 (48.5%) | 384,997 (45.0%) | 44,937 (5.2%) | 340,060 (39.7%) | | 56,264 (6.6%) |

| | | | | | | | | | |
|---|---|---|---|---|---|---|---|---|---|
| | **Sept 20-Oct 17, 2021** | **≥18** | 458,488 (53.5%) | 63,961 (14.0%) | 362,555 (79.1%) | 23,238 (5.1%) | 339,317 (74.0%) | | 31,972 (7.0%) |
| | | **≥50** | 172,442 (20.1%) | 8,311 (4.8%) | 154,314 (89.5%) | 2,802 (1.6%) | 151,512 (87.9%) | | 9,817 (5.7%) |
| **Cases** | **Sept 27-Oct 24, 2021** | **All ages** | 930,013 (100%) | 930,013 (45.7%) | 445,585 (47.9%) | 54,490 (5.9%) | 391,095 (42.1%) | | 59,414 (6.4%) |
| | | **≥18** | 518,934 (55.8%) | 70,006 (13.5%) | 414,312 (79.8%) | 24,038 (4.6%) | 390,274 (75.2%) | | 34,616 (6.7%) |
| | | **≥50** | 202,169 (21.7%) | 9,486 (4.7%) | 181,627 (89.8%) | 3,066 (1.5%) | 178,561 (88.3%) | | 11,056 (5.5%) |
| **Cases** | **Oct 4-Oct 31, 2021** | **All ages** | 975,224 (100%) | 975,224 (42.4%) | 501,078 (51.4%) | 61,288 (6.3%) | 439,790 (45.1%) | | 61,034 (6.3%) |
| | | **≥18** | 577,740 (59.2%) | 76,219 (13.2%) | 464,265 (80.4%) | 25,293 (4.4%) | 438,972 (76.0%) | | 37,256 (6.4%) |
| | | **≥50** | 229,334 (23.5%) | 10,573 (4.6%) | 206,516 (90.1%) | 3,371 (1.5%) | 203,145 (88.6%) | | 12,245 (5.3%) |
| **Cases** | **Oct 11-Nov 7, 2021** | **All ages** | 950,837 (100%) | 950,837 (39.8%) | 513,146 (54.0%) | 62,154 (6.5%) | 450,992 (47.4%) | | 59,668 (6.3%) |
| | | **≥18** | 593,357 (62.4%) | 79,516 (13.4%) | 475,740 (80.2%) | 25,554 (4.3%) | 450,186 (75.9%) | | 38,101 (6.4%) |
| | | **≥50** | 235,549 (24.8%) | 11,088 (4.7%) | 212,064 (90.0%) | 3,431 (1.5%) | 208,633 (88.6%) | | 12,397 (5.3%) |
| **Cases** | **Oct 18-Nov 14, 2021** | **All ages** | 907,478 (100%) | 907,478 (37.2%) | 513,094 (56.5%) | 63,339 (7.0%) | 449,755 (49.6%) | | 57,040 (6.3%) |
| | | **≥18** | 593,762 (65.4%) | 81,589 (13.7%) | 474,266 (79.9%) | 25,350 (4.3%) | 448,916 (75.6%) | | 37,907 (6.4%) |
| | | **≥50** | 232,221 (25.6%) | 11,498 (5.0%) | 208,679 (89.9%) | 3,405 (1.5%) | 205,274 (88.4%) | | 12,044 (5.2%) |
| **Cases** | **Oct 25-Nov 21, 2021** | **All ages** | 888,449 (100%) | 888,449 (36.8%) | 506,459 (57.0%) | 62,428 (7.0%) | 444,031 (50.0%) | | 55,147 (6.2%) |
| | | **≥18** | 588,131 (66.2%) | 83,121 (14.1%) | 468,060 (79.6%) | 24,894 (4.2%) | 443,166 (75.4%) | | 36,950 (6.3%) |
| | | **≥50** | 219,859 (24.7%) | 11,586 (5.3%) | 197,217 (89.7%) | 3,375 (1.5%) | 193,842 (88.2%) | | 11,056 (5.0%) |
| **Cases** | **Nov 1-Nov 28, 2021** | **All ages** | 905,267 (100%) | 905,267 (38.1%) | 503,519 (55.6%) | 65,115 (7.2%) | 438,404 (48.4%) | | 56,836 (6.3%) |
| | | **≥18** | 584,205 (64.5%) | 85,038 (14.6%) | 461,939 (79.1%) | 24,489 (4.2%) | 437,450 (74.9%) | | 37,228 (6.4%) |
| | | **≥50** | 205,829 (22.7%) | 11,804 (5.7%) | 183,675 (89.2%) | 3,248 (1.6%) | 180,427 (87.7%) | | 10,350 (5.0%) |
| **Cases** | **Nov 8-Dec 5, 2021** | **All ages** | 997,322 (100%) | 997,322 (39.1%) | 544,391 (54.6%) | 73,786 (7.4%) | 470,605 (47.2%) | | 62,835 (6.3%) |
| | | **≥18** | 628,872 (63.1%) | 92,746 (14.7%) | 495,707 (78.8%) | 26,242 (4.2%) | 469,465 (74.7%) | | 40,419 (6.4%) |
| | | **≥50** | 204,824 (20.5%) | 12,754 (6.2%) | 181,506 (88.6%) | 3,447 (1.7%) | 178,059 (86.9%) | | 10,564 (5.2%) |

| | | | | | | | | | |
|---|---|---|---|---|---|---|---|---|---|
| Cases | Nov 15-Dec 12, 2021 | All ages | 1,086,083 (100%) | 1,086,083 (38.5%) | 597,751 (55.0%) | 79,378 (7.3%) | 518,373 (47.7%) | | 70,726 (6.5%) |
| | | ≥18 | 697,726 (64.2%) | 104,612 (15.0%) | 546,335 (78.3%) | 29,274 (4.2%) | 517,061 (74.1%) | | 46,779 (6.7%) |
| | | ≥50 | 201,843 (18.6%) | 13,710 (6.8%) | 177,349 (87.9%) | 3,615 (1.8%) | 173,734 (86.1%) | | 10,784 (5.3%) |
| Cases | Nov 22-Dec 19, 2021 | All ages | 1,371,157 (100%) | 1,371,157 (32.5%) | 832,918 (60.7%) | 94,734 (6.9%) | 738,184 (53.8%) | | 92,902 (6.8%) |
| | | ≥18 | 975,822 (71.2%) | 132,809 (13.6%) | 775,634 (79.5%) | 39,618 (4.1%) | 736,016 (75.4%) | | 67,379 (6.9%) |
| | | ≥50 | 233,191 (17.0%) | 15,774 (6.8%) | 204,398 (87.7%) | 4,116 (1.8%) | 200,282 (85.9%) | | 13,019 (5.6%) |
| Cases | Dec 6, 2021-Jan 2, 2022 | All ages | 2,556,580 (100%) | 2,556,580 (21.5%) | 1,827,454 (71.5%) | 163,826 (6.4%) | 1,663,628 (65.1%) | | 180,064 (7.0%) |
| | | ≥18 | 2,127 (83.2%) | 240,879 (11.3%) | 1,738,627 (81.7%) | 84,102 (4.0%) | 1,654,525 (77.8%) | | 147,919 (7.0%) |
| | | ≥50 | 560,458 (21.9%) | 27,116 (4.8%) | 500,304 (89.3%) | 8,046 (1.4%) | 492,258 (87.8%) | | 33,038 (5.9%) |
| Cases | Dec 13, 2021-Jan 9, 2022 | All ages | 3,161,258 (100%) | 3,161,258 (20.3%) | 2,291,288 (72.5%) | 202,336 (6.4%) | 2,088,952 (66.1%) | | 229,101 (7.2%) |
| | | ≥18 | 2,659 (84.1%) | 292,438 (11.0%) | 2,177,293 (81.9%) | 103,771 (3.9%) | 2,073,522 (78.0%) | | 189,614 (7.1%) |
| | | ≥50 | 749,614 (23.7%) | 33,156 (4.4%) | 670,870 (89.5%) | 9,977 (1.3%) | 660,893 (88.2%) | | 45,588 (6.1%) |
| Cases | Dec 20, 2021-Jan 16, 2022 | All ages | 3,183,566 (100%) | 3,183,566 (21.9%) | 2,254,937 (70.8%) | 206,329 (6.5%) | 1,112,775 (35.0%) | 935,833 (29.4%) | 230,878 (7.3%) |
| | | ≥18 | 2,605 (81.8%) | 285,948 (11.0%) | 2,131,103 (81.8%) | 100,960 (3.9%) | 1,095,228 (42.0%) | 934,915 (35.9%) | 188,677 (7.2%) |
| | | ≥50 | 786,950 (24.7%) | 33,742 (4.3%) | 704,355 (89.5%) | 10,093 (1.3%) | 153,439 (19.5%) | 540,823 (68.7%) | 48,853 (6.2%) |
| Cases | Dec 27, 2021-Jan 23, 2022 | All ages | 3,034,409 (100%) | 3,034,409 (25.1%) | 2,058,948 (67.9%) | 199,979 (6.6%) | 844,861 (27.8%) | 1,014,108 (33.4%) | 213,040 (7.0%) |
| | | ≥18 | 2,351 (77.5%) | 255,701 (10.9%) | 1,926,523 (81.9%) | 88,213 (3.8%) | 825,293 (35.1%) | 1,013,017 (43.1%) | 168,964 (7.2%) |
| | | ≥50 | 735,711 (24.2%) | 30,660 (4.2%) | 659,811 (89.7%) | 8,958 (1.2%) | 114,649 (15.6%) | 536,204 (72.9%) | 45,240 (6.1%) |
| Cases | Jan 3-Jan 30, 2022 | All ages | 2,872,077 (100%) | 2,872,077 (29.4%) | 1,832,328 (63.8%) | 205,387 (7.2%) | 607,331 (21.1%) | 1,019,610 (35.5%) | 195,470 (6.8%) |
| | | ≥18 | 2,057 (71.6%) | 231,750 (11.3%) | 1,677,414 (81.5%) | 75,802 (3.7%) | 583,313 (28.3%) | 1,018,299 (49.5%) | 148,566 (7.2%) |
| | | ≥50 | 600,808 (20.9%) | 25,917 (4.3%) | 535,661 (89.2%) | 7,130 (1.2%) | 74,353 (12.4%) | 454,178 (75.6%) | 39,230 (6.5%) |
| Cases | | All ages | 2,293,771 (100%) | 2,293,771 (31.7%) | 1,426,498 (62.2%) | 171,086 (7.5%) | 369,295 (16.1%) | 886,117 (38.6%) | 140,444 (6.1%) |
| | | ≥18 | 1,544 (67.3%) | 159,596 (10.3%) | 1,281,180 (83.0%) | 51,615 (3.3%) | 344,772 (22.3%) | 884,793 (57.3%) | 103,389 (6.7%) |

| | | | | | | | | | |
|---|---|---|---|---|---|---|---|---|---|
| | Jan 10-Feb 6, 2022 | ≥50 | 448,840 (19.6%) | 18,505 (4.1%) | 402,612 (89.7%) | 5,060 (1.1%) | 43,512 (9.7%) | 354,040 (78.9%) | 27,723 (6.2%) |
| Cases | Jan 17-Feb 13, 2022 | All ages | 1,980,778 (100%) | 1,980,778 (29.8%) | 1,277,533 (64.5%) | 146,856 (7.4%) | 289,924 (14.6%) | 840,753 (42.4%) | 113,796 (5.7%) |
| | | ≥18 | 1,358 (68.6%) | 128,278 (9.4%) | 1,145,794 (84.3%) | 42,017 (3.1%) | 264,366 (19.5%) | 839,411 (61.8%) | 84,457 (6.2%) |
| | | ≥50 | 402,677 (20.3%) | 14,945 (3.7%) | 365,185 (90.7%) | 4,197 (1.0%) | 32,500 (8.1%) | 328,488 (81.6%) | 22,547 (5.6%) |
| Cases | Jan 24-Feb 20, 2022 | All ages | 1,578,203 (100%) | 1,578,203 (25.6%) | 1,086,434 (68.8%) | 109,803 (7.0%) | 224,505 (14.2%) | 752,126 (47.7%) | 87,739 (5.6%) |
| | | ≥18 | 1,153 (73.1%) | 100,923 (8.7%) | 984,555 (85.4%) | 33,209 (2.9%) | 200,489 (17.4%) | 750,857 (65.1%) | 68,036 (5.9%) |
| | | ≥50 | 366,989 (23.3%) | 12,308 (3.4%) | 335,767 (91.5%) | 3,484 (0.9%) | 25,606 (7.0%) | 306,677 (83.6%) | 18,914 (5.2%) |
| Cases | Jan 31-Feb 27, 2022 | All ages | 1,154,859 (100%) | 1,154,859 (21.2%) | 846,616 (73.3%) | 67,669 (5.9%) | 162,998 (14.1%) | 615,949 (53.3%) | 63,930 (5.5%) |
| | | ≥18 | 910,456 (78.8%) | 74,831 (8.2%) | 783,337 (86.0%) | 24,454 (2.7%) | 144,037 (15.8%) | 614,846 (67.5%) | 52,288 (5.7%) |
| | | ≥50 | 314,123 (27.2%) | 9,821 (3.1%) | 288,812 (91.9%) | 2,775 (0.9%) | 19,389 (6.2%) | 266,648 (84.9%) | 15,490 (4.9%) |
| Cases | Feb 7-Mar 6, 2022 | All ages | 927,841 (100%) | 927,841 (18.0%) | 710,601 (76.6%) | 41,523 (4.5%) | 124,333 (13.4%) | 544,745 (58.7%) | 49,838 (5.4%) |
| | | ≥18 | 774,595 (83.5%) | 59,904 (7.7%) | 672,261 (86.8%) | 18,878 (2.4%) | 109,575 (14.1%) | 543,808 (70.2%) | 42,430 (5.5%) |
| | | ≥50 | 291,177 (31.4%) | 8,351 (2.9%) | 269,443 (92.5%) | 2,305 (0.8%) | 16,012 (5.5%) | 251,126 (86.2%) | 13,383 (4.6%) |
| Cases | Feb 14-Mar 13, 2022 | All ages | 995,166 (100%) | 995,166 (17.3%) | 774,991 (77.9%) | 35,325 (3.5%) | 120,538 (12.1%) | 619,128 (62.2%) | 47,611 (4.8%) |
| | | ≥18 | 841,958 (84.6%) | 60,372 (7.2%) | 740,601 (88.0%) | 18,039 (2.1%) | 104,580 (12.4%) | 617,982 (73.4%) | 40,985 (4.9%) |
| | | ≥50 | 339,806 (34.1%) | 8,713 (2.6%) | 317,352 (93.4%) | 2,356 (0.7%) | 16,427 (4.8%) | 298,569 (87.9%) | 13,741 (4.0%) |
| Cases | Feb 21-Mar 20, 2022 | All ages | 1,237,498 (100%) | 1,237,498 (17.1%) | 970,347 (78.4%) | 39,117 (3.2%) | 139,110 (11.2%) | 792,120 (64.0%) | 55,172 (4.5%) |
| | | ≥18 | 1,045 (84.5%) | 70,018 (6.7%) | 928,447 (88.8%) | 20,229 (1.9%) | 117,728 (11.3%) | 790,490 (75.6%) | 47,259 (4.5%) |
| | | ≥50 | 442,044 (35.7%) | 10,397 (2.4%) | 415,176 (93.9%) | 2,733 (0.6%) | 19,477 (4.4%) | 392,966 (88.9%) | 16,471 (3.7%) |
| Cases | Feb 28-Mar 27, 2022 | All ages | 1,553,609 (100%) | 1,553,609 (16.6%) | 1,228,616 (79.1%) | 46,866 (3.0%) | 170,597 (11.0%) | 1,011,153 (65.1%) | 66,636 (4.3%) |
| | | ≥18 | 1,318 (84.9%) | 85,614 (6.5%) | 1,176,134 (89.2%) | 24,334 (1.8%) | 142,964 (10.8%) | 1,008,836 (76.5%) | 56,942 (4.3%) |
| | | ≥50 | 572,612 (36.9%) | 12,871 (2.2%) | 539,347 (94.2%) | 3,396 (0.6%) | 24,122 (4.2%) | 511,829 (89.4%) | 20,394 (3.6%) |

**Table S6a:** SARS-CoV2 cases reported by specimen date (reported weekly for prior 4 weeks period) in NIMS-database based on vaccination status since August 16, 2021 until May 27, 2022 until USHSA stopped reporting of SARS-CoV2 cases, hospitalization and deaths bases on vaccination status. *****SARS-CoV2 cases tested positive with the specimen date post receiving the first dose without additional doses of vaccination are considered one dose. ‡SARS-CoV2 cases tested positive with the specimen date ≥14 days post second dose **are** considered vaccinated with two doses. ø SARS-CoV2 cases tested positive with the specimen date ≥14 days post third dose **are** considered vaccinated with the third dose. *The age specified denominator (population) for the All ages, ≥18 years & ≥50 years of age groups based on the vaccination status (1 dose, two dose, third dose and unvaccinated), for each specified period ending date was shown on the Table S3a.*

**Table S6b: SARS-CoV2 Hospitalization within 28 days of a positive test resulting in overnight inpatient admission in NIMS-database based on vaccination status**

| Outcome | Study period (rolling four weeks) | Age | Total hospitalizations | Hospitalizations Unvaccinated | Hospitalizations Vaccinated | | | | Vaccination status unknown |
|---|---|---|---|---|---|---|---|---|---|
| | | | | | All vaccinated | 1 dose* | Two doses‡ | Third dose ø | |
| Hospitalizations | Aug 16-Sept 12, 2021 | All ages | 8,356 (100%) | 3,425 (41.0%) | 4,810 (57.6%) | 434 (5.2%) | 4,376 (52.4%) | | 121 (1.4%) |
| | | ≥18 | 7,817 (93.5%) | 2,931 (37.5%) | 4,794 (61.3%) | 418 (5.3%) | 4,376 (56.0%) | | 92 (1.2%) |
| | | ≥50 | 5,431 (65.0%) | 1,358 (25.0%) | 4,043 (74.4%) | 175 (3.2%) | 3,868 (71.2%) | | 30 (0.6%) |
| Hospitalizations | Aug 23-Sept 19, 2021 | All ages | 8,255 (100%) | 3,220 (39.0%) | 4,940 (59.8%) | 383 (4.6%%) | 4,557 (55.2%) | | 95 (1.2%) |
| | | ≥18 | 7,744 (93.8%) | 2,742 (35.4%) | 4,925 (63.6%) | 368 (4.8%) | 4,557 (58.8%) | | 77 (1.0%) |
| | | ≥50 | 5,589 (67.7%) | 1,339 (24.0%) | 4,227 (75.6%) | 164 (2.9%) | 4,063 (72.7%) | | 23 (0.4%) |
| Hospitalizations | Aug 30-Sept 26, 2021 | All ages | 7,729 (100%) | 2,922 (37.8%) | 4,734 (61.2%) | 356 (4.6%%) | 4,378 (56.6%) | | 73 (0.9%) |
| | | ≥18 | 7,204 (93.2%) | 2,435 (33.8%) | 4,710 (65.4%) | 332 (4.6%) | 4,378 (60.8%) | | 59 (0.8%) |
| | | ≥50 | 5,227 (67.6%) | 1,185 (22.7%) | 4,024 (77.0%) | 158 (3.0%) | 3,866 (74.0%) | | 18 (0.3%) |
| Hospitalizations | Sept 6-October 3, 2021 | All ages | 6,691 (100%) | 2,411 (36.0%) | 4,206 (62.9%) | 296 (4.4%%) | 3,910 (58.4%) | | 74 (1.1%) |
| | | ≥18 | 6,205 (92.7%) | 1,956 (31.5%) | 4,195 (67.6%) | 286 (4.6%) | 3,909 (63.0%) | | 54 (0.9%) |
| | | ≥50 | 4,500 (67.3%) | 931 (20.7%) | 3,551 (78.9%) | 139 (3.1%) | 3,412 (75.8%) | | 18 (0.4%) |
| Hospitalizations | Sept 13-Oct 10, 2021 | All ages | 5,148 (100%) | 1,842 (35.8%) | 3,258 (63.3%) | 224 (4.4%%) | 3,034 (58.9%) | | 48 (0.9%) |
| | | ≥18 | 4,740 (92.1%) | 1,456 (30.7%) | 3,247 (68.5%) | 214 (4.5%) | 3,033 (64.0%) | | 37 (0.8%) |
| | | ≥50 | 3,360 (65.3%) | 660 (19.6%) | 2,689 (80.0%) | 106 (3.2%) | 2,583 (76.9%) | | 11 (0.3%) |
| Hospitalizations | Sept 20-Oct 17, 2021 | All ages | 7,392 (100%) | 2,534 (34.3%) | 4,791 (64.8%) | 305 (4.1%%) | 4,486 (60.7%) | | 67 (0.9%) |
| | | ≥18 | 6,791 (91.9%) | 1,974 (29.1%) | 4,770 (70.2%) | 286 (4.2%) | 4,484 (66.0%) | | 47 (0.7%) |
| | | ≥50 | 4,946 (66.9%) | 926 (18.7%) | 3,997 (80.8%) | 169 (3.4%) | 3,828 (77.4%) | | 23 (0.5%) |
| Hospitalizations | | All ages | 8,338 (100%) | 2,832 (34.0%) | 5,429 (65.1%) | 305 (3.7%%) | 5,124 (61.5%) | | 77 (0.9%) |

| | | | | | | | | | |
|---|---|---|---|---|---|---|---|---|---|
| | **Sept 27-Oct 24, 2021** | **≥18** | 7,705 (92.4%) | 2,240 (29.1%) | 5,405 (70.1%) | 282 (3.7%) | 5,123 (66.5%) | | 60 (0.8%) |
| | | **≥50** | 5,682 (68.1%) | 1,087 (19.1%) | 4,567 (80.4%) | 158 (2.8%) | 4,409 (77.6%) | | 28 (0.5%) |
| **Hospitalizations** | **Oct 4-Oct 31, 2021** | **All ages** | 8,722 (100%) | 2,806 (32.2%) | 5,831 (66.9%) | 297 (3.4%%) | 5,534 (63.4%) | | 85 (1.0%) |
| | | **≥18** | 8,141 (93.3%) | 2,267 (27.8%) | 5,809 (71.4%) | 276 (3.4%) | 5,533 (68.0%) | | 65 (0.8%) |
| | | **≥50** | 6,147 (70.5%) | 1,158 (18.8%) | 4,956 (80.6%) | 151 (2.5%) | 4,805 (78.2%) | | 33 (0.5%) |
| **Hospitalizations** | **Oct 11-Nov 7, 2021** | **All ages** | 10,179 (100%) | 3,313 (32.5%) | 6,786 (66.7%) | 325 (3.2%%) | 6,461 (63.5%) | | 80 (0.8%) |
| | | **≥18** | 9,548 (93.8%) | 2,722 (28.5%) | 6,759 (70.8%) | 300 (3.1%) | 6,459 (67.6%) | | 67 (0.7%) |
| | | **≥50** | 7,179 (70.5%) | 1,390 (19.4%) | 5,753 (80.1%) | 153 (2.1%) | 5,600 (78.0%) | | 36 (0.5%) |
| **Hospitalizations** | **Oct 18-Nov 14, 2021** | **All ages** | 9,831 (100%) | 3,200 (32.6%) | 6,560 (66.7%) | 327 (3.3%%) | 6,233 (63.4%) | | 71 (0.7%) |
| | | **≥18** | 9,278 (94.4%) | 2,692 (29.0%) | 6,529 (70.4%) | 296 (3.2%) | 6,233 (67.2%) | | 57 (0.6%) |
| | | **≥50** | 6,956 (70.8%) | 1,391 (20.0%) | 5,538 (79.6%) | 149 (2.1%) | 5,389 (77.5%) | | 27 (0.4%) |
| **Hospitalizations** | **Oct 25-Nov 21, 2021** | **All ages** | 9,174 (100%) | 3,071 (33.5%) | 6,023 (65.7%) | 334 (3.6%%) | 5,689 (62.0%) | | 80 (0.9%) |
| | | **≥18** | 8,658 (94.4%) | 2,604 (30.1%) | 5,991 (69.2%) | 303 (3.5%) | 5,688 (65.7%) | | 63 (0.7%) |
| | | **≥50** | 6,420 (70.0%) | 1,358 (21.2%) | 5,027 (78.3%) | 154 (2.4%) | 4,873 (75.9%) | | 35 (0.5%) |
| **Hospitalizations** | **Nov 1-Nov 28, 2021** | **All ages** | 6,639 (100%) | 2,355 (35.5%) | 4,231 (63.7%) | 233 (3.5%%) | 3,998 (60.2%) | | 53 (0.8%) |
| | | **≥18** | 6,254 (94.2%) | 2,004 (32.0%) | 4,211 (67.3%) | 214 (3.4%) | 3,997 (63.9%) | | 39 (0.6%) |
| | | **≥50** | 4,618 (69.6%) | 1,077 (23.3%) | 3,520 (76.2%) | 110 (2.4%) | 3,410 (73.8%) | | 21 (0.5%) |
| **Hospitalizations** | **Nov 8-Dec 5, 2021** | **All ages** | 8,156 (100%) | 3,187 (39.1%) | 4,875 (59.8%) | 340 (4.2%%) | 4,535 (55.6%) | | 94 (1.2%%) |
| | | **≥18** | 7,575 (92.9%) | 2,676 (35.3%) | 4,830 (63.8%) | 304 (4.0%) | 4,526 (59.7%) | | 69 (0.9%) |
| | | **≥50** | 5,330 (65.4%) | 1,451 (27.2%) | 3,845 (72.1%) | 158 (3.0%) | 3,687 (69.2%) | | 34 (0.6%) |
| **Hospitalizations** | **Nov 15-Dec 12, 2021** | **All ages** | 8,235 (100%) | 3,532 (42.9%) | 4,602 (55.9%) | 346 (4.2%%) | 4,256 (51.7%) | | 101 (1.2%) |
| | | **≥18** | 7,601 (92.3%) | 2,965 (39.0%) | 4,556 (59.9%) | 309 (4.1%) | 4,247 (55.9%) | | 80 (1.1%) |
| | | **≥50** | 5,153 (62.6%) | 1,591 (30.9%) | 3,524 (68.4%) | 170 (3.3%) | 3,354 (65.1%) | | 38 (0.7%) |
| **Hospitalizations** | **Nov 22-Dec 19, 2021** | **All ages** | 8,190 (100%) | 3,693 (45.1%) | 4,388 (53.6%) | 361 (4.4%%) | 4,027 (49.2%) | | 109 (1.3%) |
| | | **≥18** | 7,542 (92.1%) | 3,115 (41.3%) | 4,339 (57.5%) | 319 (4.2%) | 4,020 (53.3%) | | 88 (1.2%) |
| | | **≥50** | 5,007 (61.1%) | 1,708 (34.1%) | 3,253 (65.0%) | 177 (3.5%) | 3,076 (61.4%) | | 46 (0.9%) |
| **Hospitalizations** | | **All ages** | 9,981 (100%) | 4,056 (40.6%) | 5,791 (58.0%) | 508 (5.1%%) | 5,283 (52.9%) | | 134 (1.3%) |
| | | **≥18** | 9,082 (91.0%) | 3,260 (35.9%) | 5,735 (63.1%) | 462 (5.1%) | 5,273 (58.1%) | | 87 (1.0%) |

| | | | | | | | | | |
|---|---|---|---|---|---|---|---|---|---|
| | Dec 6, 2021-Jan 2, 2022 | ≥50 | 5,955 (59.7%) | 1,756 (29.5%) | 4,159 (69.8%) | 234 (3.9%) | 3,925 (65.9%) | | 40 (0.7%) |
| Hospitalizations | Dec 13, 2021-Jan 9, 2022 | All ages | 13,514 (100%) | 4,738 (35.1%) | 8,566 (63.4%) | 689 (5.1%%) | 7,877 (58.3%) | | 210 (1.6%) |
| | | ≥18 | 12,219 (90.4%) | 3,614 (29.6%) | 8,473 (69.3%) | 608 (5.0%) | 7,865 (64.4%) | | 132 (1.1%) |
| | | ≥50 | 8,206 (60.7%) | 1,966 (24.0%) | 6,192 (75.5%) | 309 (3.8%) | 5,883 (71.7%) | | 48 (0.6%) |
| Hospitalizations | Dec 20, 2021-Jan 16, 2022 | All ages | 15,012 (100%) | 4,752 (31.7%) | 10,051 (67.0%) | 781 (5.2%%) | 4,127 (27.5%) | 5,143 (34.3%) | 209 (1.4%) |
| | | ≥18 | 13,503 (89.9%) | 3,445 (25.5%) | 9,925 (73.5%) | 670 (5.0%) | 4,116 (30.5%) | 5,139 (38.1%) | 133 (1.0%) |
| | | ≥50 | 9,311 (62.0%) | 1,880 (20.2%) | 7,389 (79.4%) | 340 (3.7%) | 2,438 (26.2%) | 4,611 (49.5%) | 42 (0.5%) |
| Hospitalizations | Dec 27, 2021-Jan 23, 2022 | All ages | 16,294 (100%) | 4,807 (29.5%) | 11,277 (69.2%) | 829 (5.1%%) | 3,999 (24.5%) | 6,449 (39.6%) | 210 (1.3%) |
| | | ≥18 | 14,459 (88.7%) | 3,210 (22.2%) | 11,124 (76.9%) | 701 (4.8%) | 3,978 (27.5%) | 6,445 (44.6%) | 125 (0.9%) |
| | | ≥50 | 10,270 (63.0%) | 1,753 (17.1%) | 8,462 (82.4%) | 338 (3.3%) | 2,403 (23.4%) | 5,721 (55.7%) | 55 (0.5%) |
| Hospitalizations | Jan 10-Feb 6, 2022 | All ages | 11,197 (100%) | 3,261 (29.1%) | 7,804 (69.7%) | 607 (5.4%%) | 2,121 (18.9%) | 5,076 (45.3%) | 132 (1.2%) |
| | | ≥18 | 9,463 (84.5%) | 1,737 (18.4%) | 7,641 (80.7%) | 476 (5.0%) | 2,092 (22.1%) | 5,073 (53.6%) | 85 (0.9%) |
| | | ≥50 | 6,814 (60.9%) | 946 (13.9%) | 5,827 (85.5%) | 225 (3.3%) | 1,235 (18.1%) | 4,367 (64.1%) | 41 (0.6%) |
| Hospitalizations | Jan 17-Feb 13, 2022 | All ages | 10,222 (100%) | 2,858 (28.0%) | 7,294 (71.4%) | 545 (5.3%%) | 1,810 (17.7%) | 4,939 (48.3%) | 70 (0.7%) |
| | | ≥18 | 8,599 (84.1%) | 1,441 (16.8%) | 7,118 (82.8%) | 411 (4.8%) | 1,769 (20.6%) | 4,938 (57.4%) | 40 (0.5%) |
| | | ≥50 | 6,158 (60.2%) | 791 (12.8%) | 5,349 (86.9%) | 199 (3.2%) | 996 (16.2%) | 4,154 (67.5%) | 18 (0.3%) |
| Hospitalizations | Jan 24-Feb 20, 2022 | All ages | 9,309 (100%) | 2,341 (25.1%) | 6,889 (74.0%) | 463 (5.0%%) | 1,490 (16.0%) | 4,936 (53.0%) | 79 (0.8%) |
| | | ≥18 | 7,978 (85.7%) | 1,209 (15.2%) | 6,723 (84.3%) | 340 (4.3%) | 1,449 (18.2%) | 4,934 (61.8%) | 46 (0.6%) |
| | | ≥50 | 5,858 (62.9%) | 662 (11.3%) | 5,183 (88.5%) | 166 (2.8%) | 844 (14.4%) | 4,173 (71.2%) | 12 (0.2%) |
| Hospitalizations | Jan 31-Feb 27, 2022 | All ages | 7,975 (100%) | 1,832 (23.0%) | 6,098 (76.5%) | 362 (4.5%%) | 1,178 (14.8%) | 4,558 (57.2%) | 45 (0.6%) |
| | | ≥18 | 7,015 (88.0%) | 1,020 (14.5%) | 5,973 (85.1%) | 268 (3.8%) | 1,149 (16.4%) | 4,556 (64.9%) | 22 (0.3%) |
| | | ≥50 | 5,291 (66.3%) | 560 (10.6%) | 4,726 (89.3%) | 135 (2.6%) | 692 (13.1%) | 3,899 (73.7%) | 5 (0.1%) |
| Hospitalizations | Feb 7-Mar 6, 2022 | All ages | 7,378 (100%) | 1,504 (20.4%) | 5,831 (79.0%) | 301 (4.1%%) | 1,025 (13.9%) | 4,505 (61.1%) | 43 (0.6%) |
| | | ≥18 | 6,644 (90.1%) | 882 (13.3%) | 5,740 (86.4%) | 238 (3.6%) | 1,000 (15.1%) | 4,502 (67.8%) | 22 (0.3%) |
| | | ≥50 | 5,095 (69.1%) | 483 (9.5%) | 4,607 (90.4%) | 111 (2.2%) | 627 (12.3%) | 3,869 (75.9%) | 5 (0.1%) |
| Hospitalizations | Feb 14-Mar 13, 2022 | All ages | 6,573 (100%) | 1,349 (20.5%) | 5,193 (79.0%) | 241 (3.7%%) | 837 (12.7%) | 4,115 (62.6%) | 31 (0.5%) |
| | | ≥18 | 5,915 (90.0%) | 774 (13.1%) | 5,124 (86.6%) | 190 (3.2%) | 822 (13.9%) | 4,112 (69.5%) | 17 (0.3%) |
| | | ≥50 | 4,606 (70.1%) | 409 (8.9%) | 4,192 (91.0%) | 94 (2.0%) | 518 (11.2%) | 3,580 (77.7%) | 5 (0.1%) |

| | | | | | | | | | |
|---|---|---|---|---|---|---|---|---|---|
| **Hospitalizations** | **Feb 21-Mar 20, 2022** | **All ages** | 8,990 (100%) | 1,848 (20.6%) | 7,088 (78.8%) | 309 (3.4%%) | 1,025 (11.4%) | 5,754 (64.0%) | 54 (0.6%) |
| | | **≥18** | 8,043 (89.5%) | 1,015 (12.6%) | 6,996 (87.0%) | 252 (3.1%) | 993 (12.3%) | 5,751 (71.5%) | 32 (0.4%) |
| | | **≥50** | 6,328 (70.4%) | 509 (8.0%) | 5,807 (91.8%) | 132 (2.1%) | 615 (9.7%) | 5,060 (80.0%) | 12 (0.2%) |
| **Hospitalizations** | **Feb 28-Mar 27, 2022** | **All ages** | 10,389 (100%) | 2,065 (19.9%) | 8,261 (79.5%) | 331 (3.2%%) | 1,180 (11.4%) | 6,750 (65.0%) | 63 (0.6%) |
| | | **≥18** | 9,293 (89.5%) | 1,100 (11.8%) | 8,157 (87.8%) | 270 (2.9%) | 1,142 (12.3%) | 6,745 (72.6%) | 36 (0.4%) |
| | | **≥50** | 7,329 (70.5%) | 530 (7.2%) | 6,786 (92.6%) | 148 (2.0%) | 685 (9.3%) | 5,953 (81.2%) | 12 (0.2%) |

**Table S6b:** SARS-CoV2 Hospitalization within 28 days of a positive test resulting in overnight inpatient admission (reported weekly for prior 4 weeks period) in NIMS-database based on vaccination status since August 16, 2021 until May 27, 2022 until USHSA stopped reporting of SARS-CoV2 cases, hospitalization and deaths bases on vaccination status. *SARS-CoV2 cases tested positive with the specimen date post receiving the first dose without additional doses of vaccination are considered one dose. ‡SARS-CoV2 cases tested positive with the specimen date ≥14 days post second dose **are** considered vaccinated with two doses. ø SARS-CoV2 cases tested positive with the specimen date ≥14 days post third dose **are** considered vaccinated with the third dose. *The age specified denominator (population) for the All ages, ≥18 years & ≥50 years of age groups based on the vaccination status (1 dose, two doses, third dose and unvaccinated), for each specified period ending date was shown on the Table S3a.*

#### Table S6c: SARS-CoV2 deaths within 28 days of positive COVID-19 test by date of death in NIMS-database based on vaccination

| Outcome | Study period (rolling four weeks) | Age (yrs) | Total deaths (n=% of all ages) | Unvaccinated deaths (n=% for age group) | Vaccinated deaths (n=% for age group) | | | | Vaccination status unknown |
|---|---|---|---|---|---|---|---|---|---|
| | | | | | All Vaccinated | 1 dose* | Two doses‡ | Third dose ø | |
| Deaths | Aug 16-Sept 12, 2021 | All ages | 2,961 (100%) | 726 (24.5%) | 2,206 (74.5%) | 112 (3.8%) | 2,094 (70.7%) | | 29 (1.0%) |
| | | ≥18 | 2,957 (99.9%) | 723 (24.5%) | 2,206 (74.6%) | 112 (3.8%) | 2,094 (70.8%) | | 28 (0.9%) |
| | | ≥50 | 2,775 (93.7%) | 599 (21.6%) | 2,153 (77.6%) | 101 (3.6%) | 2,052 (73.9%) | | 23 (0.8%) |
| Deaths | Aug 23-Sept 19, 2021 | All ages | 3,158 (100%) | 730 (23.1%) | 2,395 (75.8%) | 111 (3.5%) | 2,284 (72.3%) | | 33 (1.0%) |
| | | ≥18 | 3,152 (99.8%) | 726 (23.0%) | 2,395 (76.0%) | 111 (3.5%) | 2,284 (72.5%) | | 31 (1.0%) |
| | | ≥50 | 2,983 (94.5%) | 615 (20.6%) | 2,342 (78.5%) | 103 (3.5%) | 2,239 (75.1%) | | 26 (0.9%) |
| Deaths | Aug 30-Sept 26, 2021 | All ages | 3,165 (100%) | 687 (21.7%) | 2,448 (77.3%) | 110 (3.5%) | 2,338 (73.9%) | | 30 (0.9%) |
| | | ≥18 | 3,159 (99.8%) | 683 (21.6%) | 2,448 (77.5%) | 110 (3.5%) | 2,338 (74.0%) | | 28 (0.9%) |
| | | ≥50 | 3,005 (94.9%) | 586 (19.5%) | 2,396 (79.7%) | 103 (3.4%) | 2,293 (76.3%) | | 23 (0.8%) |
| Deaths | Sept 6-October 3, 2021 | All ages | 3,026 (100%) | 611 (20.2%) | 2,382 (78.7%) | 101 (3.3%) | 2,281 (75.4%) | | 33 (1.1%) |
| | | ≥18 | 3,020 (99.8%) | 609 (20.2%) | 2,381 (78.8%) | 100 (3.3%) | 2,281 (75.5%) | | 30 (1.0%) |
| | | ≥50 | 2,887 (95.4%) | 522 (18.1%) | 2,341 (81.1%) | 95 (3.3%) | 2,246 (77.8%) | | 24 (0.8%) |
| Deaths | Sept 13-Oct 10, 2021 | All ages | 2,805 (100%) | 557 (19.9%) | 2,228 (79.4%) | 92 (3.3%) | 2,136 (76.1%) | | 20 (0.7%) |
| | | ≥18 | 2,801 (99.9%) | 554 (19.8%) | 2,227 (79.5%) | 91 (3.2%) | 2,136 (76.3%) | | 20 (0.7%) |
| | | ≥50 | 2,689 (95.9%) | 487 (18.1%) | 2,185 (81.3%) | 88 (3.3%) | 2,097 (78.0%) | | 17 (0.6%) |
| Deaths | Sept 20-Oct 17, 2021 | All ages | 2,745 (100%) | 502 (18.3%) | 2,224 (81.0%) | 88 (3.2%) | 2,136 (77.8%) | | 19 (0.7%) |
| | | ≥18 | 2,740 (99.8%) | 498 (18.2%) | 2,223 (81.1%) | 87 (3.2%) | 2,136 (78.0%) | | 19 (0.7%) |
| | | ≥50 | 2,635 (96.0%) | 433 (16.4%) | 2,186 (83.0%) | 86 (3.3%) | 2,100 (79.7%) | | 16 (0.6%) |
| Deaths | | All ages | 2,772 (100%) | 487 (17.6%) | 2,270 (81.9%) | 85 (3.1%) | 2,185 (78.8%) | | 15 (0.5%) |
| | | ≥18 | 2,767 (99.8%) | 483 (17.5%) | 2,269 (82.0%) | 84 (3.0%) | 2,185 (79.0%) | | 15 (0.5%) |

| | | | | | | | | | |
|---|---|---|---|---|---|---|---|---|---|
| | **Sept 27-Oct 24, 2021** | **≥50** | 2,666 (96.2%) | 423 (15.9%) | 2,230 (83.6%) | 82 (3.1%) | 2,148 (80.6%) | | 13 (0.5%) |
| **Deaths** | **Oct 4-Oct 31, 2021** | **All ages** | 3,085 (100%) | 538 (17.4%) | 2,532 (82.1%) | 85 (2.8%) | 2,447 (79.3%) | | 15 (0.5%) |
| | | **≥18** | 3,079 (99.8%) | 532 (17.3%) | 2,532 (82.2%) | 85 (2.8%) | 2,447 (79.5%) | | 15 (0.5%) |
| | | **≥50** | 2,972 (96.3%) | 471 (15.8%) | 2,488 (83.7%) | 82 (2.8%) | 2,406 (81.0%) | | 13 (0.4%) |
| **Deaths** | **Oct 11-Nov 7, 2021** | **All ages** | 3,430 (100%) | 587 (17.1%) | 2,822 (82.3%) | 90 (2.6%) | 2,732 (79.7%) | | 21 (0.6%) |
| | | **≥18** | 3,422 (99.8%) | 580 (16.9%) | 2,822 (82.5%) | 90 (2.6%) | 2,732 (79.8%) | | 20 (0.6%) |
| | | **≥50** | 3,297 (96.1%) | 511 (15.5%) | 2,769 (84.0%) | 86 (2.6%) | 2,683 (81.4%) | | 17 (0.5%) |
| **Deaths** | **Oct 18-Nov 14, 2021** | **All ages** | 3,676 (100%) | 675 (18.4%) | 2,975 (80.9%) | 100 (2.7%) | 2,875 (78.2%) | | 26 (0.7%) |
| | | **≥18** | 3,665 (99.7%) | 665 (18.1%) | 2,975 (81.2%) | 100 (2.7%) | 2,875 (78.4%) | | 25 (0.7%) |
| | | **≥50** | 3,522 (95.8%) | 587 (16.7%) | 2,914 (82.7%) | 91 (2.6%) | 2,823 (80.2%) | | 21 (0.6%) |
| **Deaths** | **Oct 25-Nov 21, 2021** | **All ages** | 3,726 (100%) | 708 (19.0%) | 2,992 (80.3%) | 89 (2.4%) | 2,903 (77.9%) | | 26 (0.7%) |
| | | **≥18** | 3,717 (99.8%) | 700 (18.8%) | 2,992 (80.5%) | 89 (2.4%) | 2,903 (78.1%) | | 25 (0.7%) |
| | | **≥50** | 3,556 (95.4%) | 612 (17.2%) | 2,923 (82.2%) | 79 (2.2%) | 2,844 (80.0%) | | 21 (0.6%) |
| **Deaths** | **Nov 1-Nov 28, 2021** | **All ages** | 3,571 (100%) | 695 (19.5%) | 2,846 (79.7%) | 96 (2.7%) | 2,750 (77.0%) | | 30 (0.8%) |
| | | **≥18** | 3,563 (99.8%) | 688 (19.3%) | 2,846 (79.9%) | 96 (2.7%) | 2,750 (77.2%) | | 29 (0.8%) |
| | | **≥50** | 3,415 (95.6%) | 606 (17.7%) | 2,785 (81.6%) | 87 (2.5%) | 2,698 (79.0%) | | 24 (0.7%) |
| **Deaths** | **Nov 8-Dec 5, 2021** | **All ages** | 3,310 (100%) | 708 (21.4%) | 2,572 (77.7%) | 92 (2.8%) | 2,480 (74.9%) | | 30 (0.9%) |
| | | **≥18** | 3,306 (99.9%) | 704 (21.3%) | 2,572 (77.8%) | 92 (2.8%) | 2,480 (75.0%) | | 30 (0.9%) |
| | | **≥50** | 3,151 (95.2%) | 619 (19.6%) | 2,506 (79.5%) | 85 (2.7%) | 2,421 (76.8%) | | 26 (0.8%) |
| **Deaths** | **Nov 15-Dec 12, 2021** | **All ages** | 3,087 (100%) | 718 (23.3%) | 2,341 (75.8%) | 82 (2.7%) | 2,259 (73.2%) | | 28 (0.9%) |
| | | **≥18** | 3,083 (99.9%) | 715 (23.2%) | 2,340 (75.9%) | 82 (2.7%) | 2,258 (73.2%) | | 28 (0.9%) |
| | | **≥50** | 2,920 (94.6%) | 618 (21.2%) | 2,277 (78.0%) | 76 (2.6%) | 2,201 (75.4%) | | 25 (0.9%) |
| **Deaths** | **Nov 22-Dec 19, 2021** | **All ages** | 2,956 (100%) | 782 (26.5%) | 2,140 (72.4%) | 90 (3.0%) | 2,050 (69.4%) | | 34 (1.2%) |
| | | **≥18** | 2,952 (99.9%) | 779 (26.4%) | 2,139 (72.5%) | 90 (3.0%) | 2,049 (69.4%) | | 34 (1.2%) |
| | | **≥50** | 2,772 (93.8%) | 667 (24.1%) | 2,074 (74.8%) | 84 (3.0%) | 1,990 (71.8%) | | 31 (1.1%) |
| **Deaths** | | **All ages** | 2,890 (100%) | 809 (28.0%) | 2,055 (71.1%) | 106 (3.7%) | 1,949 (67.4%) | | 26 (0.9%) |
| | | **≥18** | 2,885 (99.8%) | 805 (27.9%) | 2,054 (71.2%) | 106 (3.7%) | 1,948 (67.5%) | | 26 (0.9%) |

| | | | | | | | | | |
|---|---|---|---|---|---|---|---|---|---|
| | Dec 6, 2021-Jan 2, 2022 | ≥50 | 2,703 (93.5%) | 694 (25.7%) | 1,985 (73.4%) | 99 (3.7%) | 1,886 (69.8%) | | 24 (0.9%) |
| Deaths | Dec 13, 2021-Jan 9, 2022 | All ages | 3,174 (100%) | 924 (29.1%) | 2,219 (69.9%) | 130 (4.1%) | 2,089 (65.8%) | | 31 (1.0%) |
| | | ≥18 | 3,166 (99.7%) | 916 (28.9%) | 2,219 (70.1%) | 130 (4.1%) | 2,089 (66.0%) | | 31 (1.0%) |
| | | ≥50 | 2,964 (93.4%) | 795 (26.8%) | 2,140 (72.2%) | 122 (4.1%) | 2,018 (68.1%) | | 29 (1.0%) |
| Deaths | Dec 20, 2021-Jan 16, 2022 | All ages | 3,893 (100%) | 1,015 (26.1%) | 2,845,73 (1%) | 171,4 (4%) | 1,418 (36.4%) | 1,256 (32.3%) | 33 (0.8%) |
| | | ≥18 | 3,884 (99.8%) | 1,006 (25.9%) | 2,845,73 (2%) | 171 (4.4%) | 1,418 (36.5%) | 1,256 (32.3%) | 33 (0.8%) |
| | | ≥50 | 3,672 (94.3%) | 898 (24.5%) | 2,743 (74.7%) | 158 (4.3%) | 1,344 (36.6%) | 1,241 (33.8%) | 31 (0.8%) |
| Deaths | Dec 27, 2021-Jan 23, 2022 | All ages | 4,637 (100%) | 1,010 (21.8%) | 3,591,77 (4%) | 190 (4.1%) | 1,561 (33.7%) | 1,840 (39.7%) | 36 (0.8%) |
| | | ≥18 | 4,624 (99.7%) | 1,000 (21.6%) | 3,588,77 (6%) | 188 (4.1%%) | 1,560 (33.7%) | 1,840 (39.8%) | 36 (0.8%) |
| | | ≥50 | 4,423 (95.4%) | 903 (20.4%) | 3,486 (78.8%) | 174 (3.9%) | 1,489 (33.7%) | 1,823 (41.2%) | 34 (0.8%) |
| Deaths | Jan 3-Jan 30, 2022 | All ages | 5,554 (100%) | 1,015 (18.3%) | 4,501,81 (0%) | 213 (3.8%) | 1,703 (30.7%) | 2,585 (46.5%) | 38 (0.7%) |
| | | ≥18 | 5,542 (99.8%) | 1,007 (18.2%) | 4,497,81 (1%) | 211 (3.8%) | 1,701 (30.7%) | 2,585 (46.6%) | 38 (0.7%) |
| | | ≥50 | 5,310 (95.6%) | 912 (17.2%) | 4,362 (82.1%) | 191 (3.6%) | 1,624 (30.6%) | 2,547 (48.0%) | 36 (0.7%) |
| Deaths | Jan 10-Feb 6, 2022 | All ages | 5,978 (100%) | 911 (15.2%) | 5,035 (84.2%) | 208 (3.5%) | 1,631 (27.3%) | 3,196 (53.5%) | 32 (0.5%) |
| | | ≥18 | 5,967 (99.8%) | 904 (15.1%) | 5,031 (84.3%) | 206 (3.5%) | 1,629 (27.3%) | 3,196 (53.6%) | 32 (0.5%) |
| | | ≥50 | 5,752 (96.2%) | 829 (14.4%) | 4,894 (85.1%) | 189 (3.3%) | 1,556 (27.1%) | 3,149 (54.7%) | 29 (0.5%) |
| Deaths | Jan 17-Feb 13, 2022 | All ages | 5,592 (100%) | 728 (13.0%) | 4,836 (86.5%) | 179 (3.2%) | 1,350 (24.1%) | 3,307 (59.1%) | 28 (0.5%) |
| | | ≥18 | 5,584 (99.9%) | 723 (12.9%) | 4,833 (86.6%) | 178 (3.2%) | 1,348 (24.1%) | 3,307 (59.2%) | 28 (0.5%) |
| | | ≥50 | 5,404 (96.6%) | 659 (12.2%) | 4,720 (87.3%) | 161 (3.0%) | 1,299 (24.0%) | 3,260 (60.3%) | 25 (0.5%) |
| Deaths | Jan 24-Feb 20, 2022 | All ages | 4,883 (100%) | 559 (11.4%) | 4,302 (88.1%) | 147 (3.0%) | 1,035 (21.2%) | 3,120 (63.9%) | 22 (0.5%) |
| | | ≥18 | 4,877 (99.9%) | 555 (11.4%) | 4,300 (88.2%) | 147 (3.0%) | 1,033 (21.2%) | 3,120 (64.0%) | 22 (0.5%) |
| | | ≥50 | 4,731 (96.9%) | 506 (10.7%) | 4,206 (88.9%) | 131 (2.8%) | 991 (20.9%) | 3,084 (65.2%) | 19 (0.4%) |
| Deaths | Jan 31-Feb 27, 2022 | All ages | 3,957 (100%) | 397 (10.0%) | 3,542 (89.5%) | 113 (2.9%) | 725 (18.3%) | 2,704 (68.3%) | 18 (0.5%) |
| | | ≥18 | 3,953 (99.9%) | 394 (10.0%) | 3,541 (89.6%) | 112 (2.8%) | 725 (18.3%) | 2,704 (68.4%) | 18 (0.5%) |
| | | ≥50 | 3,843 (97.1%) | 359 (9.3%) | 3,469 (90.3%) | 103 (2.7%) | 689 (17.9%) | 2,677 (69.7%) | 15 (0.4%) |
| Deaths | Feb 7-Mar 6, 2022 | All ages | 2,943 (100%) | 286 (9.7%) | 2,643 (89.8%) | 79 (2.7%) | 482 (16.4%) | 2,082 (70.7%) | 14 (0.5%) |
| | | ≥18 | 2,940 (99.9%) | 284 (9.7%) | 2,642 (89.9%) | 78 (2.7%) | 482 (16.4%) | 2,082 (70.8%) | 14 (0.5%) |
| | | ≥50 | 2,867 (97.4%) | 260 (9.1%) | 2,594 (90.5%) | 70 (2.4%) | 458 (16.0%) | 2,066 (72.1%) | 13 (0.5%) |

| | | | | | | | | | |
|---|---|---|---|---|---|---|---|---|---|
| **Deaths** | **Feb 14-Mar 13, 2022** | **All ages** | 2,421 (100%) | 237 (9.8%) | 2,173 (89.8%) | 58 (2.4%) | 382 (15.8%) | 1,733 (71.6%) | 11 (0.5%) |
| | | **≥18** | 2,419 (99.9%) | 236 (9.8%) | 2,172 (89.8%) | 57 (2.4%) | 382 (15.8%) | 1,733 (71.6%) | 11 (0.5%) |
| | | **≥50** | 2,364 (97.6%) | 216 (9.1%) | 2,139 (90.5%) | 52 (2.2%) | 365 (15.4%) | 1,722 (72.8%) | 9 (0.4%) |
| **Deaths** | **Feb 21-Mar 20, 2022** | **All ages** | 2,084 (100%) | 202 (9.7%) | 1,873 (89.9%) | 49 (2.4%) | 333 (16.0%) | 1,491 (71.5%) | 9 (0.4%) |
| | | **≥18** | 2,083 (99.95%) | 202 (9.7%) | 1,872 (89.9%) | 48 (2.3%) | 333 (16.0%) | 1,491 (71.6%) | 9 (0.4%) |
| | | **≥50** | 2,030 (97.4%) | 186 (9.2%) | 1,836 (90.4%) | 45 (2.2%) | 315 (15.5%) | 1,476 (72.7%) | 8 (0.4%) |
| **Deaths** | **Feb 28-Mar 27, 2022** | **All ages** | 2,144 (100%) | 214 (10.0%) | 1,773 (82.7%) | 1 (0.05%) | 215 (10.0%) | 1,557 (72.6%) | 10 (0.5%) |
| | | **≥18** | 2,142 (99.9%) | 213 (9.9%) | 1,772 (82.7%) | 1 (0.05%) | 214 (10.0%) | 1,557 (72.7%) | 10 (0.5%) |
| | | **≥50** | 2,090 (97.5%) | 199 (9.5%) | 1,736 (83.1%) | 1 (0.05%) | 200 (9.6%) | 1,535 (73.4%) | 9 (0.4%) |

**Table S6c:** SARS-CoV2 deaths within 28 days of positive COVID-19 test by date of death (reported weekly for prior 4 weeks period) in NIMS-database based on vaccination status since August 16, 2021 until May 27, 2022 until USHSA stopped reporting of SARS-CoV2 cases, hospitalization and deaths bases on vaccination status. *SARS-CoV2 cases tested positive with the specimen date post receiving the first dose without additional doses of vaccination are considered one dose. ‡SARS-CoV2 cases tested positive with the specimen date ≥14 days post second dose **are** considered vaccinated with two doses. ᴓ SARS-CoV2 cases tested positive with the specimen date ≥14 days post third dose **are** considered vaccinated with the third dose. *The age specified denominator (population) for the All ages, ≥18 years & ≥50 years of age groups based on the vaccination status (1 dose, two doses, third dose and unvaccinated), for each specified period ending date was shown on the Table S3a.*

#### Table S7a: Vaccine effectiveness of over 18 years of age NIMS population

| Rolling Four weeks period | SARS-CoV2 cases; ≥18 yrs of age (during four weeks period) | | | | Incidence rate per 100,000 population | | | | Vaccine effectiveness (95% CI) | | |
|---|---|---|---|---|---|---|---|---|---|---|---|
| | 1 dose | 2 dose | 3rd dose | Unvaccinated | 1 dose | 2 doses | 3rd dose | Unvaccinated | 1 dose* | Two doses‡ | Third dose ø |
| Aug 16-Sept 12, 2021 | 81332 | 288470 | | 101867 | 2706.5 | 789.2 | | 956.2 | -183.1% (-187% to -178.9%) | 17.5% (16.5% to 18.4%) | |
| Aug 23-Sept 19, 2021 | 55159 | 276831 | | 87377 | 2036.0 | 750.2 | | 826.5 | -146.3% (-150.3% to -142.2%) | 9.2% (8.1% to 10.3%) | |
| Aug 30-Sept 26, 2021 | 40434 | 272498 | | 75925 | 1605.8 | 740.9 | | 723.3 | -122% (-126% to -117.8%) | -2.4% (-3.7% to -1.2%) | |
| Sept 6-October 3, 2021 | 28361 | 265485 | | 64589 | 1188.1 | 736.6 | | 619.0 | -91.9% (-96% to -87.7%) | -19.0% (-20.6% to -17.4%) | |
| Sept 13-Oct 10, 2021 | 23218 | 287527 | | 60497 | 1014.8 | 821.7 | | 583.5 | -73.9% (-78% to -69.9%) | -40.8% (-42.7% to -39.0%) | |
| Sept 20-Oct 17, 2021 | 23238 | 339317 | | 63961 | 1053.2 | 1003.9 | | 620.7 | -69.7% (-73.7% to -65.7%) | -61.7% (-63.8% to -59.7%) | |
| Sept 27-Oct 24, 2021 | 24038 | 390274 | | 70006 | 1122.2 | 1205.2 | | 683.7 | -64.1% (-67.9% to -60.4%) | -76.3% (-78.4% to -74.2%) | |
| Oct 4-Oct 31, 2021 | 25293 | 438972 | | 76219 | 1211.9 | 1419.6 | | 748.6 | -61.9% (-65.5% to -58.4%) | -89.6% (-91.8% to -87.5%) | |
| Oct 11-Nov 7, 2021 | 25554 | 450186 | | 79516 | 1256.0 | 1540.8 | | 785.1 | -60.0% (63.5% to -56.5%) | -96.3% (-98.5% to -94.1%) | |

| | | | | | | | | | | | |
|---|---|---|---|---|---|---|---|---|---|---|---|
| **Oct 18-Nov 14, 2021** | 25350 | 448916 | | 81589 | 1282.2 | 1642.0 | | 810.1 | -58.3% (-61.8% to -54.9%) | -102.7% (-105.0% to -100.5%) | |
| **Oct 25-Nov 21, 2021** | 24894 | 443166 | | 83121 | 1291.3 | 1744.6 | | 829.8 | -55.6% (-59.0% to -52.2%) | -110.2% (-112.6% to -107.9%) | |
| **Nov 1-Nov 28, 2021** | 24489 | 437450 | | 85038 | 1305.0 | 1872.1 | | 853.7 | -52.9% (-56.2% to -49.6%) | -119.3% (-121.7% to -116.9%) | |
| **Nov 8-Dec 5, 2021** | 26242 | 469465 | | 92746 | 1438.0 | 2208.1 | | 937.1 | -53.4% (-56.7% to -50.2%) | -135.6% (-138.1% to -133.2%) | |
| **Nov 15-Dec 12, 2021** | 29274 | 517061 | | 104612 | 1649.3 | 2740.9 | | 1064.4 | -54.9% (-58.0% to -51.9%) | -157.5% (-160.1% to -155.0%) | |
| **Nov 22-Dec 19, 2021** | 39618 | 736016 | | 132809 | 2283.1 | 5153.1 | | 1367.3 | -67.0% (-69.9% to -64.1%) | -276.9% (-280.1% to -273.6%) | |
| **Dec 6, 2021-Jan 2, 2022** | 84102 | 1654525 | | 240879 | 4927.5 | 16023.7 | | 2520.4 | -95.5% (-97.9% to -93.2%) | -535.8% (-539.7% to -531.9%) | |
| **Dec 13, 2021-Jan 9, 2022** | 103771 | 2073522 | | 292438 | 6136.3 | 22404.2 | | 3084.8 | -98.9% (-101.1% to -96.8%) | -626.3% (-630.3% to -622.3%) | |
| **Dec 20, 2021-Jan 16, 2022** | 100960 | 1095228 | 934915 | 285948 | 5993.8 | 12552.7 | 3075.2 | 3037.8 | -97.3% (-99.5% to -95.2%) | -313.2% (-315.8% to -310.7%) | -1.2% (-1.9% to -0.6%) |
| **Dec 27, 2021-Jan 23, 2022** | 88213 | 825293 | 1013017 | 255701 | 5259.6 | 9761.1 | 3296.0 | 2732.5 | -92.5% (-94.7% to -90.2%) | -257.2% (-259.6% to -254.8%) | -20.6% (-21.4% to -19.8%) |
| **Jan 3-Jan 30, 2022** | 75802 | 583313 | 1018299 | 231750 | 4543.0 | 7076.5 | 3284.3 | 2489.9 | -82.5% (-84.8% to -80.2%) | -184.2% (-186.3% to -182.1%) | -31.9% (-32.8% to -31.0%) |
| **Jan 10-Feb 6, 2022** | 51615 | 344772 | 884793 | 159596 | 3118.9 | 4251.0 | 2837.3 | 1720.9 | -81.2% (-84.0% to -78.5%) | -147.0% (-149.3% to -144.7%) | -64.9% (-66.2% to -63.6%) |

| | | | | | | | | | | | |
|---|---|---|---|---|---|---|---|---|---|---|---|
| **Jan 17-Feb 13, 2022** | 42017 | 264366 | 839411 | 128278 | 2570.7 | 3306.4 | 2677.9 | 1387.0 | -85.3% (-88.5% to - 82.2%) | -138.4% (-140.9% to - 135.9%) | -93.1% (- 94.8% to - 91.4%) |
| **Jan 24-Feb 20, 2022** | 33209 | 200489 | 750857 | 100923 | 2055.5 | 2537.0 | 2385.4 | 1093.5 | -88.0% (-91.6% to - 84.4%) | -132.0% (-134.8% to - 129.2%) | -118.1% (- 120.2% to - 116.1%) |
| **Jan 31-Feb 27, 2022** | 24454 | 144037 | 614846 | 74831 | 1530.3 | 1840.8 | 1946.2 | 812.5 | -88.3% (-92.6% to - 84.2%) | -126.6% (-129.8% to - 123.4%) | -139.5% (- 142.2% to - 136.9%) |
| **Feb 7-Mar 6, 2022** | 18878 | 109575 | 543808 | 59904 | 1194.5 | 1410.8 | 1716.4 | 651.6 | -83.3% (-88.0% to - 78.7%) | -116.5% (-120.0% to - 113.1%) | -163.4% (- 166.6% to - 160.2%) |
| **Feb 14-Mar 13, 2022** | 18039 | 104580 | 617982 | 60372 | 1155.3 | 1355.1 | 1945.4 | 657.7 | -75.6% (-80.2% to - 71.2%) | -106.0% (-109.3% to - 102.8%) | -195.8% (- 199.3% to - 192.3%) |
| **Feb 21-Mar 20, 2022** | 20229 | 117728 | 790490 | 70018 | 1311.9 | 1534.3 | 2482.4 | 764.0 | -71.7% (-75.9% to - 67.6%) | -100.8% (-103.9% to - 97.9%) | -224.9% (- 228.5% to - 221.4%) |
| **Feb 28-Mar 27, 2022** | 24334 | 142964 | 1008836 | 85614 | 1594.9 | 1873.8 | 3160.9 | 935.3 | -70.5% (-74.3% to - 66.8%) | -100.3% (-103.1% to - 97.7%) | -237.9% (- 241.3% to - 234.7%) |

**Table S7a: Vaccine effectiveness of over 18 years of age NIMS population (rolling four weeks cases reported weekly) since August 16, 2021 to March 27, 2022** until UKHSA stopped reporting data of weekly cases among age groups based on vaccination status.

*****SARS-CoV2 cases tested positive with the specimen date post receiving the first dose without additional doses of vaccination are considered one dose. ‡SARS-CoV2 cases tested positive with the specimen date ≥14 days post second dose **are** considered vaccinated with two doses. ø SARS-CoV2 cases tested positive with the specimen date ≥14 days post third dose **are** considered vaccinated with the third dose. *The age specified denominator (population) for the ≥18 years of age group based on the vaccination status (1 dose, two doses, third dose and unvaccinated), for each specified period ending date was shown on the Table S3a.*

#### Table S7b: Vaccine effectiveness of over 50 years of age confirmed Delta variant SARS-CoV2 cases reported by Public Health England

| Time period | Confirmed Delta variant COVID-19 cases (≥50 yrs of age) | | | Incidence rate per 100,000 population | | | Vaccine effectiveness (95% CI) | |
|---|---|---|---|---|---|---|---|---|
| | 1 dose | 2 doses | Unvaccinated | 1 dose | 2 doses | Unvaccinated | 1 dose | 2 doses |
| **Feb 1-June 20, 2021** | 4651 | 5234 | 1267 | 516.7 | 27.0 | 57.5 | -797.9% (-889.6% to -717.1%) | 53.1% (48.4% to 57.2%) |
| **June 21-July 18, 2021** | 795 | 8193 | 1070 | 138.1 | 41.4 | 50.5 | -173.7% (-217.1% to -135.8%) | 18.0% (9.8% to 25.2%) |
| **July 19-Aug 1, 2021** | 389 | 8045 | 1103 | 75.5 | 40.5 | 52.7 | -43.3% (-71.1% to -19.0%) | 23.2% (15.6% to 29.8%) |
| **Aug 2-Aug 15, 2021** | 468 | 11356 | 1451 | 98.5 | 56.9 | 70.0 | -40.7% (-65.0% to -19.1%) | 18.6% (11.7% to 24.8%) |
| **Aug 16-Aug 29, 2021** | 596 | 18592 | 1833 | 134.1 | 93.0 | 89.1 | -50.4% (-73.3% to -29.8%) | -4.3% (-11.9% to 2.5%) |
| **Aug 30-Sept 12, 2021** | 544 | 20571 | 1827 | 128.3 | 102.7 | 89.4 | -43.5% (-66.0% to -23.3%) | -14.9% (-23.1% to -7.5%) |

**Table S7b: Vaccine effectiveness of over 50 years of age confirmed Delta variant SARS-CoV2 cases reported by Public Health England until September 12, 2021.** *SARS-CoV2 cases tested positive with the specimen date post receiving the first dose without additional doses of vaccination are considered one dose. ‡SARS-CoV2 cases tested positive with the specimen date ≥14 days post second dose **are** considered vaccinated with two doses. *The age specified denominator (population) for the ≥50 years of age group based on the vaccination status (1 dose, two doses and unvaccinated), for each specified period ending date was shown on the Table S3a.*

### Table S7c: Vaccine effectiveness of over 50 years of age NIMS population

| Rolling Four weeks period | COVID-19 cases; ≥50 yrs of age (during four weeks period) | | | | Incidence rate per 100,000 population | | | | Vaccine effectiveness (95% CI) | | |
|---|---|---|---|---|---|---|---|---|---|---|---|
| | 1 dose* | Two doses‡ | Third dose ø | Unvaccinated | 1 dose | 2 doses | 3rd dose | Unvaccinated | 1 dose | 2 doses | 3rd dose |
| Aug 16-Sept 12, 2021 | 81332 | 288470 | | 101867 | 843.4 | 711.7 | | 551.3 | -183.1% (-187% to -178.9%) | 17.5% (16.5% to 18.4%) | |
| Aug 23-Sept 19, 2021 | 55159 | 276831 | | 87377 | 755.4 | 674.7 | | 503.1 | -146.3% (-150.3% to -142.2%) | 9.2% (8.1% to 10.3%) | |
| Aug 30-Sept 26, 2021 | 40434 | 272498 | | 75925 | 692.7 | 646.3 | | 449.8 | -122% (-126% to -117.8%) | -2.4% (-3.7% to -1.2%) | |
| Sept 6-October 3, 2021 | 28361 | 265485 | | 64589 | 651.6 | 635.4 | | 397.1 | -91.9% (-96% to -87.7%) | -19.0% (-20.6% to -17.4%) | |
| Sept 13-Oct 10, 2021 | 23218 | 287527 | | 60497 | 656.8 | 712.5 | | 381.1 | -73.9% (-78% to -69.9%) | -40.8% (-42.7% to -39.0%) | |
| Sept 20-Oct 17, 2021 | 23238 | 339317 | | 63961 | 753.7 | 902.2 | | 413.2 | -69.7% (-73.7% to -65.7%) | -61.7% (-63.8% to -59.7%) | |
| Sept 27-Oct 24, 2021 | 24038 | 390274 | | 70006 | 841.2 | 1155.6 | | 473.3 | -64.1% (-67.9% to -60.4%) | -76.3% (-78.4% to -74.2%) | |
| Oct 4-Oct 31, 2021 | 25293 | 438972 | | 76219 | 941.3 | 1444.1 | | 529.4 | -61.9% (-65.5% to -58.4%) | -89.6% (-91.8% to -87.5%) | |
| Oct 11-Nov 7, 2021 | 25554 | 450186 | | 79516 | 974.0 | 1672.1 | | 557.2 | -60.0% (63.5% to -56.5%) | -96.3% (-98.5% to -94.1%) | |
| Oct 18-Nov 14, 2021 | 25350 | 448916 | | 81589 | 986.9 | 1906.1 | | 580.0 | -58.3% (-61.8% to -54.9%) | -102.7% (-105.0% to -100.5%) | |
| Oct 25-Nov 21, 2021 | 24894 | 443166 | | 83121 | 1001.2 | 2136.7 | | 586.9 | -55.6% (-59.0% to -52.2%) | -110.2% (-112.6% to -107.9%) | |

| | | | | | | | | | | | |
|---|---|---|---|---|---|---|---|---|---|---|---|
| **Nov 1-Nov 28, 2021** | 24489 | 437450 | | 85038 | 1008.7 | 2442.3 | | 600.4 | -52.9% (-56.2% to -49.6%) | -119.3% (-121.7% to -116.9%) | |
| **Nov 8-Dec 5, 2021** | 26242 | 469465 | | 92746 | 1119.7 | 3083.5 | | 651.9 | -53.4% (-56.7% to -50.2%) | -135.6% (-138.1% to -133.2%) | |
| **Nov 15-Dec 12, 2021** | 29274 | 517061 | | 104612 | 1209.8 | 4066.0 | | 704.8 | -54.9% (-58.0% to -51.9%) | -157.5% (-160.1% to -155.0%) | |
| **Nov 22-Dec 19, 2021** | 39618 | 736016 | | 132809 | 1402.3 | 6920.1 | | 818.3 | -67.0% (-69.9% to -64.1%) | -276.9% (-280.1% to -273.6%) | |
| **Dec 6, 2021-Jan 2, 2022** | 84102 | 1654525 | | 240879 | 2774.2 | 23991.2 | | 1421.8 | -95.5% (-97.9% to -93.2%) | -535.8% (-539.7% to -531.9%) | |
| **Dec 13, 2021-Jan 9, 2022** | 103771 | 2073522 | | 292438 | 3462.9 | 35837.9 | | 1746.3 | -98.9% (-101.1% to -96.8%) | -626.3% (-630.3% to -622.3%) | |
| **Dec 20, 2021-Jan 16, 2022** | 100960 | 1095228 | 934915 | 285948 | 3518.0 | 8823.1 | 2910.4 | 1783.9 | -97.3% (-99.5% to -95.2%) | -313.2% (-315.8% to -310.7%) | -1.2% (-1.9% to -0.6%) |
| **Dec 27, 2021-Jan 23, 2022** | 88213 | 825293 | 1013017 | 255701 | 3132.7 | 6802.7 | 2876.2 | 1625.9 | -92.5% (-94.7% to -90.2%) | -257.2% (-259.6% to -254.8%) | -20.6% (-21.4% to -19.8%) |
| **Jan 3-Jan 30, 2022** | 75802 | 583313 | 1018299 | 231750 | 2502.4 | 4536.1 | 2429.3 | 1378.3 | -82.5% (-84.8% to -80.2%) | -184.2% (-186.3% to -182.1%) | -31.9% (-32.8% to -31.0%) |
| **Jan 10-Feb 6, 2022** | 51615 | 344772 | 884793 | 159596 | 1786.6 | 2700.8 | 1890.4 | 985.7 | -81.2% (-84.0% to -78.5%) | -147.0% (-149.3% to -144.7%) | -64.9% (-66.2% to -63.6%) |
| **Jan 17-Feb 13, 2022** | 42017 | 264366 | 839411 | 128278 | 1494.3 | 2044.6 | 1751.5 | 797.0 | -85.3% (-88.5% to -82.2%) | -138.4% (-140.9% to -135.9%) | -93.1% (-94.8% to -91.4%) |
| **Jan 24-Feb 20, 2022** | 33209 | 200489 | 750857 | 100923 | 1250.3 | 1628.2 | 1633.4 | 657.0 | -88.0% (-91.6% to -84.4%) | -132.0% (-134.8% to -129.2%) | -118.1% (-120.2% to -116.1%) |
| **Jan 31-Feb 27, 2022** | 24454 | 144037 | 614846 | 74831 | 1002.5 | 1246.0 | 1418.7 | 524.8 | -88.3% (-92.6% to -84.2%) | -126.6% (-129.8% to -123.4%) | -139.5% (-142.2% to -136.9%) |

| | | | | | | | | | | | |
|---|---|---|---|---|---|---|---|---|---|---|---|
| **Feb 7-Mar 6, 2022** | 18878 | 109575 | 543808 | 59904 | 838.4 | 1037.5 | 1335.0 | 446.6 | -83.3% (-88.0% to -78.7%) | -116.5% (-120.0% to -113.1%) | -163.4% (-166.6% to -160.2%) |
| **Feb 14-Mar 13, 2022** | 18039 | 104580 | 617982 | 60372 | 863.1 | 1072.1 | 1585.9 | 466.3 | -75.6% (-80.2% to -71.2%) | -106.0% (-109.3% to -102.8%) | -195.8% (-199.3% to -192.3%) |
| **Feb 21-Mar 20, 2022** | 20229 | 117728 | 790490 | 70018 | 1009.3 | 1279.1 | 2085.9 | 556.7 | -71.7% (-75.9% to -67.6%) | -100.8% (-103.9% to -97.9%) | -224.9% (-228.5% to -221.4%) |
| **Feb 28-Mar 27, 2022** | 24334 | 142964 | 1008836 | 85614 | 1262.0 | 1596.3 | 2714.8 | 689.7 | -70.5% (-74.3% to -66.8%) | -100.3% (-103.1% to -97.7%) | -237.9% (-241.3% to -234.7%) |

**Table S7c: Vaccine effectiveness of over 50 years of age NIMS population (rolling four weeks cases reported weekly) since August 16, 2021 to March 27, 2022** until UKHSA stopped reporting data of weekly cases among age groups based on vaccination status.

*SARS-CoV2 cases tested positive with the specimen date post receiving the first dose without additional doses of vaccination are considered one dose. ‡SARS-CoV2 cases tested positive with the specimen date ≥14 days post second dose **are** considered vaccinated with two doses. ø SARS-CoV2 cases tested positive with the specimen date ≥14 days post third dose **are** considered vaccinated with the third dose. *The age specified denominator (population) for the ≥50 years of age group based on the vaccination status (1 dose, two doses, third dose and unvaccinated), for each specified period ending date was shown on the Table S3a.*

**Table S7d:** **The comparable SARS-CoV2 cases per 100,000 population of entire population (all ages), based on the NIMS population estimate and England vaccinated population database estimate**

| Study period | SARS-CoV2 infections (all ages, n=total cases) | | | | Incidence rate per 100,000 population (NIMS database estimate) | | | | Incidence rate per 100,000 population (England vaccinated database estimate) | | | |
|---|---|---|---|---|---|---|---|---|---|---|---|---|
| | 1 dose only | Two doses (no 3rd dose) | Third dose | Unvaccinated | 1 dose only | Two doses (no 3rd dose) | Third dose | Unvaccinated | 1 dose only | Two doses (no 3rd dose) | This dose | Unvaccinated |
| Feb 1-June 20, 2021 | 26495 | 10834 | | 71932 | 278.8 | 41.2 | | 263.5 | 276.9 | 40.8 | | 351.8 |
| June 21-July 18, 2021 | 27596 | 17939 | | 49470 | 322.6 | 59.9 | | 201.3 | 319.0 | 59.4 | | 279.7 |
| July 19-Aug 1, 2021 | 16016 | 18235 | | 29652 | 225.6 | 57.2 | | 122.8 | 222.5 | 56.8 | | 172.0 |
| Aug 2-Aug 15, 2021 | 19850 | 26364 | | 32079 | 353.8 | 78.2 | | 135.1 | 345.9 | 77.6 | | 190.5 |
| Aug 16-Aug 29, 2021 | 18913 | 40451 | | 36583 | 427.5 | 114.0 | | 157.8 | 413.6 | 113.3 | | 225.0 |
| Aug 30-Sept 12, 2021 | 11942 | 43577 | | 37641 | 332.4 | 119.0 | | 164.7 | 317.1 | 118.2 | | 236.5 |
| Aug 16-Sept 12, 2021 | 93143 | 289279 | | 263285 | 2592.3 | 789.7 | | 1151.7 | 2473.4 | 784.7 | | 1653.9 |
| Aug 23-Sept 19, 2021 | 67187 | 277504 | | 275845 | 2019.7 | 750.3 | | 1212.7 | 1915.2 | 745.1 | | 1746.2 |
| Aug 30-Sept 26, 2021 | 53070 | 273180 | | 316002 | 1669.8 | 741.0 | | 1396.7 | 1578.1 | 728.3 | | 2015.8 |
| Sept 6-October 3, 2021 | 39232 | 266094 | | 337570 | 1240.5 | 736.4 | | 1504.2 | 1177.1 | 730.8 | | 2176.5 |
| Sept 13-Oct 10, 2021 | 37578 | 288181 | | 371696 | 1173.4 | 821.1 | | 1672.5 | 1127.1 | 812.9 | | 2423.0 |
| Sept 20-Oct 17, 2021 | 44937 | 340060 | | 415109 | 1370.8 | 1002.4 | | 1888.2 | 1336.5 | 990.2 | | 2739.8 |
| Sept 27-Oct 24, 2021 | 54490 | 391095 | | 425014 | 1611.0 | 1202.4 | | 1955.9 | 1570.6 | 1186.9 | | 2853.5 |

| | | | | | | | | | | | | |
|---|---|---|---|---|---|---|---|---|---|---|---|---|
| Oct 4-Oct 31, 2021 | 61288 | 439790 | | 413112 | 1768.8 | 1414.9 | | 1919.9 | 1712.9 | 1396.6 | | 2818.6 |
| Oct 11-Nov 7, 2021 | 62154 | 450992 | | 378023 | 1725.9 | 1534.4 | | 1778.1 | 1673.5 | 1512.3 | | 2625.3 |
| Oct 18-Nov 14, 2021 | 63339 | 449755 | | 337344 | 1719.3 | 1633.8 | | 1602.8 | 1641.6 | 1617.0 | | 2392.2 |
| Oct 25-Nov 21, 2021 | 62428 | 444031 | | 326843 | 1676.4 | 1734.1 | | 1564.9 | 1597.6 | 1715.8 | | 2345.9 |
| Nov 1-Nov 28, 2021 | 65115 | 438404 | | 344912 | 1762.1 | 1856.0 | | 1661.9 | 1669.1 | 1837.2 | | 2501.1 |
| Nov 8-Dec 5, 2021 | 73786 | 470605 | | 390096 | 2031.5 | 2179.3 | | 1891.8 | 1907.9 | 2158.4 | | 2860.6 |
| Nov 15-Dec 12, 2021 | 79378 | 518373 | | 417606 | 2225.9 | 2690.6 | | 2037.7 | 2078.9 | 2660.1 | | 3093.6 |
| Nov 22-Dec 19, 2021 | 94734 | 738184 | | 445337 | 2696.6 | 5000.9 | | 2193.0 | 2510.8 | 4888.4 | | 3348.5 |
| Dec 6, 2021-Jan 2, 2022 | 163826 | 1663628 | | 549062 | 4773.0 | 15199.9 | | 2738.8 | 4437.1 | 14948.5 | | 4216.3 |
| Dec 13, 2021-Jan 9, 2022 | 202336 | 2088952 | | 640869 | 6001.1 | 20935.7 | | 3219.7 | 5563.9 | 20590.9 | | 4979.0 |
| Dec 20, 2021-Jan 16, 2022 | 206329 | 1112775 | 935833 | 697751 | 6238.1 | 11656.3 | 3072.4 | 3526.4 | 5765.3 | 11471.5 | 3065.6 | 5475.0 |
| Dec 27, 2021-Jan 23, 2022 | 199979 | 844861 | 1014108 | 762421 | 6215.7 | 8997.3 | 3291.9 | 3872.7 | 5727.5 | 8863.9 | 3283.2 | 6032.3 |
| Jan 3-Jan 30, 2022 | 205387 | 607331 | 1019610 | 844279 | 6587.8 | 6532.8 | 3279.8 | 4307.9 | 6043.0 | 6441.5 | 3270.0 | 6731.5 |
| Jan 10-Feb 6, 2022 | 171086 | 369295 | 886117 | 726829 | 5641.9 | 3987.0 | 2833.3 | 3721.5 | 5161.7 | 3932.6 | 2824.0 | 5830.3 |
| Jan 17-Feb 13, 2022 | 146856 | 289924 | 840753 | 589449 | 4978.1 | 3137.2 | 2673.9 | 3028.1 | 4542.9 | 3095.7 | 2664.5 | 4754.3 |
| Jan 24-Feb 20, 2022 | 109803 | 224505 | 752126 | 404030 | 3795.5 | 2437.7 | 2381.5 | 2081.0 | 3460.8 | 2406.4 | 2372.5 | 3272.6 |
| Jan 31-Feb 27, 2022 | 67669 | 162998 | 615949 | 244313 | 2380.7 | 1776.5 | 1942.3 | 1261.3 | 2169.8 | 1753.8 | 1934.5 | 1986.7 |
| Feb 7-Mar 6, 2022 | 41523 | 124333 | 544745 | 167402 | 1489.6 | 1357.9 | 1711.8 | 865.9 | 1356.1 | 1342.1 | 1704.1 | 1365.6 |
| Feb 14-Mar 13, 2022 | 35325 | 120538 | 619128 | 172564 | 1293.5 | 1318.5 | 1939.2 | 894.1 | 1177.1 | 1303.8 | 1930.1 | 1411.4 |

| | | | | | | | | | | | | |
|---|---|---|---|---|---|---|---|---|---|---|---|---|
| Feb 21-Mar 20, 2022 | 39117 | 139110 | 792120 | 211979 | 1463.5 | 1523.5 | 2473.5 | 1100.0 | 1331.9 | 1507.4 | 2461.2 | 1737.4 |
| Feb 28-Mar 27, 2022 | 46866 | 170597 | 1011153 | 258357 | 1787.0 | 1871.2 | 3148.5 | 1342.5 | 1625.8 | 1851.7 | 3132.5 | 2122.0 |

**Table S7d: The comparable SARS-CoV2 cases per 100,000 population of entire population (all ages), of the Delta variant SARS CoV2 cases in England (Feb 1 - Sept 12, 2021) and NIMS database linked SARS-CoV2 cases (August 16, 2021 - March 27, 2022) based on the NIMS population estimate and England vaccinated population database estimate.** The denominator (population) for each period ending date was based on the respective population numbers among vaccinated groups of NIMS database (Table S3a) and England vaccination database (Table S3c). As shown on the table the incidence rate per 100,000 population is similar among vaccinated populations (1 dose, two doses and third dose) and how ever the unvaccinated population differences as shown Tables Table S3a and Table S3c variability among unvaccinated incidence rate per 100, 000 population.

**Table S7e: The comparable vaccine effectiveness of entire population (all ages) based on the NIMS population denominator estimate and the England vaccinated population database denominator estimate.**

| Study period | Vaccine effectiveness (95% CI) NIMS vaccinated Population | | | Vaccine effectiveness (95% CI) England vaccinated population | | |
|---|---|---|---|---|---|---|
| | 1 dose VE (95%CI) | 2 doses VE (95%CI) | VE 3rd dose (95% CI) | 1 dose (95% CI) | 2 doses VE (95%CI) | VE 3rd dose (95% CI) |
| Feb 1-June 20, 2021 | -5.8% (-8.2% to -3.5%) | 84.4% (83.9% to 84.8%) | | 21.3% (19.5% to 23.0%) | 88.8% (88.0% to 88.7%) | |
| June 21-July 18, 2021 | -60.2% (-64.0% to -56.5%) | 70.3% (69.5% to 71.1%) | | -14.1% (-16.8% to -11.4%) | 78.7% (78.2% to 79.3%) | |
| July 19-Aug 1, 2021 | -83.7% (-89.4% to -78.1%) | 53.4% (52.0% to 54.8%) | | -29.4% (-33.4% to -25.4%) | 67.6% (66.0% to 67.9%) | |
| Aug 2-Aug 15, 2021 | -161.9% (-169.4% to -154.6%) | 42.1% (40.6% to 43.6%) | | - 81.6% (-86.8% to -76.5%) | 59.5% (58.2% to 60.3%) | |
| Aug 16-Aug 29, 2021 | -170.9% (-178.6% to -163.4%) | 27.7% (26.1% to 29.4%) | | -83.8 (-89.0% to -78.8%) | 49.4% (48.5% to 50.8%) | |
| Aug 30-Sept 12, 2021 | -101.9% (-108.4% to -95.5%) | 27.8% (26.1% to 29.3%) | | -34.1% (-38.5% to -29.9%) | 50.5% (48.9% to 51.1%) | |
| Aug 16-Sept 12, 2021 | -125.1% (-127.7% to -122.5%) | 31.4% (30.9% to 32.0%) | | -49.6% (-51.3% to -47.8%) | 52.6% (52.2% to 53.0%) | |
| Aug 23-Sept 19, 2021 | -66.5% (-68.7% to -64.4%) | 38.1% (37.6% to 38.6%) | | -9.7% (-11.1% to -8.3%) | 57.3% (57.0% to 57.7%) | |
| Aug 30-Sept 26, 2021 | -19.5% (-21.2% to -17.9%) | 46.9% (46.5% to 47.4%) | | 21.7% (20.6% to -22.8%) | 63.9% (63.6% to 64.2%) | |
| Sept 6-October 3, 2021 | 17.5% (16.3% to -18.8%) | 51.0% (50.6% to 51.4%) | | 45.9% (45.1% to -46.7%) | 66.4% (66.1% to 66.7%) | |
| Sept 13-Oct 10, 2021 | 29.8% (28.8% to -30.9%) | 50.9% (50.5% to 51.3%) | | 53.5% (52.8% to -54.2%) | 66.4% (66.2% to 66.7%) | |
| Sept 20-Oct 17, 2021 | 27.4% (26.4% to -28.4%) | 46.9% (46.5% to 47.3%) | | 51.2% (50.5% to -51.9%) | 63.9% (63.6% to 64.1%) | |
| Sept 27-Oct 24, 2021 | 17.6% (16.6% to -18.7%) | 38.5% (38.1% to 39.0%) | | 45.0% (44.2% to -45.7%) | 58.4% (58.1% to 58.7%) | |
| Oct 4-Oct 31, 2021 | 7.9% (6.7% to -9.0%) | 26.3% (25.8% to 26.8%) | | 39.2% (38.5% to -40.0%) | 50.5% (50.1% to 50.8%) | |
| Oct 11-Nov 7, 2021 | 2.9% (1.7% to -4.1%) | 13.7% (13.1% to 14.3%) | | 36.3% (35.5% to -37.1%) | 42.4% (42.0% to 42.8%) | |
| Oct 18-Nov 14, 2021 | -7.3% (-8.6% to -5.9%) | -1.9% (-2.7% to -1.2%) | | 31.4% (30.5% to -32.2%) | 32.4% (31.9% to 32.9%) | |

| | | | | | | |
|---|---|---|---|---|---|---|
| **Oct 25-Nov 21, 2021** | -7.1% (-8.5% to -5.8%) | -10.8% (-11.6% to -10.0%) | | 31.9% (31.0% to -32.8%) | 26.9% (26.3% to 27.4%) | |
| **Nov 1-Nov 28, 2021** | -6.0% (-7.4% to -4.7%) | -11.7% (-12.5% to -10.9%) | | 33.3% (32.4% to -34.1%) | 26.5% (26.0% to 27.1%) | |
| **Nov 8-Dec 5, 2021** | -7.4% (-8.6% to -6.1%) | -15.2% (-16.0% to -14.4%) | | 33.3% (32.5% to -34.1%) | 24.5% (24.0% to 25.1%) | |
| **Nov 15-Dec 12, 2021** | -9.2% (-10.5% to -8.0%) | -32.0% (-32.9% to -31.2%) | | 32.8% (32.0% to -33.6%) | 14.0% (13.5% to 14.6%) | |
| **Nov 22-Dec 19, 2021** | -23.0% (-24.3% to -21.7%) | -128.0% (-129.4% to -126.7%) | | 25.0% (24.2% to -25.8%) | -46.0% (-46.8% to -45.1%) | |
| **Dec 6, 2021-Jan 2, 2022** | -74.3% (-75.7% to -72.8%) | -455.0% (-457.5% to -452.5%) | | -5.2% (-6.1% to -4.4%) | -254.5% (-256.2% to -252.9%) | |
| **Dec 13, 2021-Jan 9, 2022** | -86.4% (-87.8% to -85.0%) | -550.2% (-552.9% to -547.5%) | | -11.7% (-12.6% to -10.9%) | -313.6% (-315.3% to -311.9%) | |
| **Dec 20, 2021-Jan 16, 2022** | -76.9% (-78.2% to -75.6%) | -230.5% (-232.1% to -229.0%) | 12.9% (12.4% to 13.3%) | -5.3% (-6.1% to -4.5%) | -109.5% (-110.5% to -108.6%) | 44.0% (43.7% to 44.3%) |
| **Dec 27, 2021-Jan 23, 2022** | -60.5% (-61.7% to -59.3%) | -132.3% (-133.5% to -131.2%) | 15.0% (14.6% to 15.4%) | 5.1% (4.4% to -5.7%) | -46.9% (-47.7% to -46.2%) | 45.6% (45.3% to 45.8%) |
| **Jan 3-Jan 30, 2022** | -52.9% (-54.0% to -51.8%) | -51.6% (-52.4% to -50.9%) | 23.9% (23.5% to 24.2%) | 10.2% (9.6% to -10.9%) | 4.3% (3.8% to 4.8%) | 51.4% (51.2% to 51.6%) |
| **Jan 10-Feb 6, 2022** | -51.6% (-52.8% to -50.4%) | -7.1% (-7.8% to -6.5%) | 23.9% (23.5% to 24.2%) | 11.5% (10.8% to -12.2%) | 32.5% (32.1% to 33.0%) | 51.6% (51.3% to 51.8%) |
| **Jan 17-Feb 13, 2022** | -64.4% (-65.8% to -63.0%) | -3.6% (-4.3% to -2.9%) | 11.7% (11.2% to 12.2%) | 4.4% (3.6% to -5.3%) | 34.9% (34.4% to 35.3%) | 44.0% (43.7% to 44.3%) |
| **Jan 24-Feb 20, 2022** | -82.4% (-84.2% to -80.6%) | -17.1% (-18.1% to -16.2%) | -14.4% (-15.1% to -13.8%) | -5.7% (-6.8% to -4.7%) | 26.5% (25.9% to 27.1%) | 27.5% (27.1% to 27.9%) |
| **Jan 31-Feb 27, 2022** | -88.7% (-91.2% to -86.3%) | -40.8% (-42.3% to -39.4%) | -54.0% (-55.1% to -52.9%) | -9.2% (-10.6% to -7.8%) | 11.7% (10.8% to 12.6%) | 2.6% (1.9% to 3.3%) |
| **Feb 7-Mar 6, 2022** | -72.0% (-74.9% to -69.2%) | -56.8% (-58.7% to -55.0%) | -97.7% (-99.4% to -96.0%) | 0.7% (-0.9% to -2.3%) | 1.7% (0.6% to 2.9%) | -24.8% (-25.8% to -23.7%) |
| **Feb 14-Mar 13, 2022** | -44.7% (-47.2% to -42.2%) | -47.5% (-49.2% to -45.7%) | -116.9% (-118.7% to -115.1%) | 16.6% (15.2% to -18.0%) | 7.6% (6.5% to 8.7%) | -36.8% (-37.9% to -35.6%) |
| **Feb 21-Mar 20, 2022** | -33.0% (-35.2% to -30.9%) | -38.5% (-40.0% to -37.0%) | -124.9% (-126.5% to -123.2%) | 23.3% (22.1% to -24.6%) | 13.2% (12.3% to 14.2%) | -41.7% (-42.7% to -40.6%) |
| **Feb 28-Mar 27, 2022** | -33.1% (-35.1% to -31.2%) | -39.4% (-40.7% to -38.0%) | -134.5% (-136.1% to -133.0%) | 23.4% (22.3% to -24.5%) | 12.7% (11.9% to 13.6%) | -47.6% (-48.6% to -46.7%) |

**Table S7e: The comparable vaccine effectiveness of entire population (all ages) of the Delta variant SARS CoV2 cases in England (Feb 1-Sept 12, 2021) and NIMS database linked SARS-CoV2 cases (August 16, 2021-March 27, 2022) based on the NIMS**

**population denominator estimate and the England vaccinated population database denominator estimate.** The denominator (population) for each period ending date was based on the respective population numbers among vaccinated groups of NIMS database (Table S3a) and England vaccination database (Table S3c).

**Table S8a-f: Proportion test of SARS-CoV2 cases, hospitalizations and deaths among NIMS vaccinated population of over 18 and over 50 years of age.**

| Study period (rolling 4 weeks) | Proportion of population (cases/population) | | | | Compared groups | Statistical analysis Two-Proportions test with continuity adjustment (X2=; df=; p-value) |
|---|---|---|---|---|---|---|
| | All vaccinated | 2 doses‡ | Third dose ø | Unvaccinated | | |
| **Aug 16-Sept 12, 2021** | 0.009345 | 0.007892 | | 0.009562 | All vaccinated vs. Unvaccinated<br>Two doses vs. Unvaccinated | $X^2$ = 42.277, df = 1, p- = 1<br>$X^2$= 2804.2, df = 1, p-= 1 |
| **Aug 23-Sept 19, 2021** | 0.008373 | 0.007502 | | 0.008265 | All vaccinated vs. Unvaccinated<br>Two doses vs. Unvaccinated | $X^2$ = 11.686, df = 1, p = 0.0003149<br>$X^2$ = 627.65, df = 1, p = 1 |
| **Aug 30-Sept 26, 2021** | 0.007877 | 0.007409 | | 0.007233 | All vaccinated vs. Unvaccinated<br>Two doses vs. Unvaccinated | $X^2$= 448.21, df = 1, p<0.001<br>$X^2$= 34.737, df = 1, p<0.001 |
| **Sept 13-Oct 10, 2021** | 0.007797 | 0.008217 | | 0.005835 | All vaccinated vs. Unvaccinated<br>Two doses vs. Unvaccinated | $X^2$= 4317.8, df = 1, p<0.001<br>$X^2$= 5961.9, df = 1, p<0.001 |
| **Oct 11-Nov 7, 2021** | 0.011865 | 0.015408 | | 0.007851 | All vaccinated vs. Unvaccinated<br>Two doses vs. Unvaccinated | $X^2$= 11915, df = 1, p<0.001<br>$X^2$= 32339, df = 1, p<0.001 |
| **Nov 8-Dec 5, 2021** | 0.012292 | 0.022081 | | 0.009371 | All vaccinated vs. Unvaccinated<br>Two doses vs. Unvaccinated | $X^2$= 5855.7, df = 1, p<0.001<br>$X^2$= 61570, df = 1, p<0.001 |
| **Dec 6, 2021-Jan 2, 2022** | 0.042753 | 0.160237 | | 0.025204 | All vaccinated vs. Unvaccinated<br>Two doses vs. Unvaccinated | $X^2$= 62948, df = 1, p<0.001<br>$X^2$= 1049374, df = 1, p<0.001 |
| **Dec 20, 2021-Jan 16, 2022** | 0.052219 | 0.125527 | 0.030752 | 0.030378 | All vaccinated vs. Unvaccinated<br>Third dose vs. Unvaccinated<br>Two doses vs. Unvaccinated<br>Third dose vs. Two doses | $X^2$= 79651, df = 1, p<0.001<br>$X^2$= 33.892, df = 1, p<0.001<br>$X^2$= 582725, df = 1, p<0.001<br>$X^2$= 1237850, df = 1, p = 1 |

| | | | | | | |
|---|---|---|---|---|---|---|
| **Dec 27, 2021-Jan 23, 2022** | 0.062683 | 0.097611 | 0.032960 | 0.027325 | All vaccinated vs. Unvaccinated<br>Third dose vs. Unvaccinated<br>Two doses vs. Unvaccinated<br>Third dose vs. Two doses | $X^2$= 174246, df = 1, p<0.001<br>$X^2$= 7433.2, df = 1, p<0.001<br>$X^2$ = 384927, df = 1, p<0.001<br>$X^2$= 619910, df = 1, p = 1 |
| **Jan 3-Jan 30, 2022** | 0.054101 | 0.070765 | 0.032843 | 0.024899 | All vaccinated vs. Unvaccinated<br>Third dose vs. Unvaccinated<br>Two doses vs. Unvaccinated<br>Third dose vs. Two doses | $X^2$= 135316, df = 1, p<0.001<br>$X^2$ = 15037, df = 1, p<0.001<br>$X^2$= 207668, df = 1, p<0.001<br>$X^2$ = 239234, df = 1, p = 1 |
| **Jan 17-Feb 13, 2022** | 0.027963 | 0.033064 | 0.026779 | 0.013870 | All vaccinated vs. Unvaccinated<br>Third dose vs. Unvaccinated<br>Two doses vs. Unvaccinated<br>Third dose vs. Two doses | $X^2$= 60619, df = 1, p<0.001<br>$X^2$ = 51150, df = 1, p<0.001<br>$X^2$= 71007, df = 1, p<0.001<br>$X^2$ = 9227.7, df = 1, p = 1 |
| **Jan 31-Feb 27, 2022** | 0.019099 | 0.018408 | 0.019462 | 0.008125 | All vaccinated vs. Unvaccinated<br>Third dose vs. Unvaccinated<br>Two doses vs. Unvaccinated<br>Third dose vs. Two doses | $X^2$ = 53922, df = 1, p<0.001<br>$X^2$ = 55147, df = 1, p<0.001<br>$X^2$ = 35266, df = 1, p<0.001<br>$X^2$ = 368.53, df = 1, p<0.001 |
| **Feb 14-Mar 13, 2022** | 0.018043 | 0.013551 | 0.019454 | 0.006577 | All vaccinated vs. Unvaccinated<br>Third dose vs. Unvaccinated<br>Two doses vs. Unvaccinated<br>Third dose vs. Two doses | $X^2$ = 62838, df = 1, p<0.001<br>$X^2$ = 72465, df = 1, p<0.001<br>$X^2$ = 21090, df = 1, p<0.001<br>$X^2$ = 12041, df = 1, p<0.001 |
| **Feb 28-Mar 27, 2022** | 0.028637 | 0.018738 | 0.031609 | 0.009353 | All vaccinated vs. Unvaccinated<br>Third dose vs. Unvaccinated<br>Two doses vs. Unvaccinated<br>Third dose vs. Two doses | $X^2$ = 113642, df = 1, p<0.001<br>$X^2$ = 135833, df = 1, p<0.001<br>$X^2$ = 27278, df = 1, p<0.001<br>$X^2$ = 36077, df = 1, p<0.001 |

**Table S8a: Two-proportions test with continuity adjustment of SARS-CoV2 cases among NIMS vaccinated population of over 18 years of age since August 16, 2021 to March 27, 2022.** [‡]SARS-CoV2 cases tested positive with the specimen date ≥14 days post second dose **are** considered vaccinated with two doses. [ø] SARS-CoV2 cases tested positive with the specimen date ≥14 days post third dose **are** considered vaccinated with the third dose.

| Study period (rolling 4 weeks) | **Proportion of population** (hospitalizations/population) | | | | Groups compared | Statistical analysis |
|---|---|---|---|---|---|---|
| | All vaccinated | 2 doses‡ | Third dose ø | Unvaccinated | | **Two-Proportions test with continuity adjustment** (X2=; df=; p-value) |
| **Aug 16-Sept 12, 2021** | 0.000121 | 0.000120 | | 0.000275 | All vaccinated vs. Unvaccinated<br>Two doses vs. Unvaccinated | $X^2$ = 1292.9, df = 1, p = 1<br>$X^2$ = 1286.1, df = 1, p = 1 |
| **Aug 23-Sept 19, 2021** | 0.000124 | 0.000123 | | 0.000259 | All vaccinated vs. Unvaccinated<br>Two doses vs. Unvaccinated | $X^2$ = 998.05, df = 1, p = 1<br>$X^2$ = 985.86, df = 1, p = 1 |
| **Aug 30-Sept 26, 2021** | 0.000119 | 0.000119 | | 0.000232 | All vaccinated vs. Unvaccinated<br>Two doses vs. Unvaccinated | $X^2$ = 750.01, df = 1, p = 1<br>$X^2$ = 721.94, df = 1, p = 1 |
| **Sept 13-Oct 10, 2021** | 0.000081 | 0.000087 | | 0.000140 | All vaccinated vs. Unvaccinated<br>Two doses vs. Unvaccinated | $X^2$ = 304.78, df = 1, p = 1<br>X-squared = 232.96, df = 1, p = 1 |
| **Oct 11-Nov 7, 2021** | 0.000169 | 0.000221 | | 0.000269 | All vaccinated vs. Unvaccinated<br>Two doses vs. Unvaccinated | $X^2$ = 429.48, df = 1, p- = 1<br>$X^2$ = 73.123, df = 1, p = 1 |
| **Nov 8-Dec 5, 2021** | 0.000120 | 0.000213 | | 0.000270 | All vaccinated vs. Unvaccinated<br>Two doses vs. Unvaccinated | $X^2$ = 1205.4, df = 1, p = 1<br>$X^2$ = 96.403, df = 1, p = 1 |
| **Nov 22-Dec 19, 2021** | 0.000107 | 0.000281 | | 0.000321 | All vaccinated vs. Unvaccinated<br>Two doses vs. Unvaccinated | $X^2$ = 2407.3, df = 1, p = 1<br>$X^2$ = 29.835, df = 1, p = 1 |
| **Dec 6, 2021-Jan 2, 2022** | 0.000141 | 0.000511 | | 0.000341 | All vaccinated vs. Unvaccinated<br>Two doses vs. Unvaccinated | $X^2$ = 1728.9, df = 1, p = 1<br>$X^2$ = 332.3, df = 1, p<0.001 |
| **Dec 20, 2021-Jan 16, 2022** | 0.000243 | 0.000472 | 0.000169 | 0.000366 | All vaccinated vs. Unvaccinated<br>Third dose vs. Unvaccinated<br>Two doses vs. Unvaccinated<br>Third dose vs. Two doses | $X^2$ = 432.84, df = 1, p= 1<br>$X^2$ = 1292.4, df = 1, p = 1<br>$X^2$ = 121.31, df = 1, p<0.001<br>$X^2$ = 2625.6, df = 1, p = 1 |
| **Dec 27, 2021-Jan 23, 2022** | 0.000272 | 0.000470 | 0.000210 | 0.000343 | All vaccinated vs. Unvaccinated<br>Third dose vs. Unvaccinated<br>Two doses vs. Unvaccinated<br>Third dose vs. Two doses | $X^2$ = 133.64, df = 1, p = 1<br>$X^2$ = 529.15, df = 1, p = 1<br>$X^2$ = 178.58, df = 1, p<0.001<br>$X^2$ = 1695.2, df = 1, p = 1 |

| | | | | | | |
|---|---|---|---|---|---|---|
| **Jan 10-Feb 6, 2022** | 0.000187 | 0.000258 | 0.000163 | 0.000187 | All vaccinated vs. Unvaccinated<br>Third dose vs. Unvaccinated<br>Two doses vs. Unvaccinated<br>Third dose vs. Two doses | $X^2$ = 0.016292, df = 1, p = 0.5508<br>$X^2$ = 25.596, df = 1, p = 1<br>$X^2$ = 97.746, df = 1, p<0.001<br>$X^2$ = 319.9, df = 1, p = 1 |
| **Jan 17-Feb 13, 2022** | 0.000174 | 0.000221 | 0.000158 | 0.000156 | All vaccinated vs. Unvaccinated<br>Third dose vs. Unvaccinated<br>Two doses vs. Unvaccinated<br>Third dose vs. Two doses | $X^2$ = 14.103, df = 1, p<0.001<br>$X^2$ = 0.12568, df = 1, p- = 0.3615<br>$X^2$ = 98.344, df = 1, p<0.001<br>$X^2$ = 151.35, df = 1, p = 1 |
| **Jan 24-Feb 20, 2022** | 0.000164 | 0.000183 | 0.000157 | 0.000131 | All vaccinated vs. Unvaccinated<br>Third dose vs. Unvaccinated<br>Two doses vs. Unvaccinated<br>Third dose vs. Two doses | $X^2$ = 52.732, df = 1, p<0.001<br>$X^2$ = 33.446, df = 1, p<0.001<br>$X^2$ = 75.858, df = 1, p<0.001<br>$X^2$ = 25.972, df = 1, p = 1 |
| **Jan 31-Feb 27, 2022** | 0.000146 | 0.000147 | 0.000144 | 0.000111 | All vaccinated vs. Unvaccinated<br>Third dose vs. Unvaccinated<br>Two doses vs. Unvaccinated<br>Third dose vs. Two doses | $X^2$ = 65.454, df = 1, p<0.001<br>$X^2$ = 58.17, df = 1, p<0.001<br>$X^2$ = 42.996, df = 1, p<0.001<br>$X^2$ = 0.28271, df = 1, p = 0.7025 |
| **Feb 14-Mar 13, 2022** | 0.000125 | 0.000107 | 0.000129 | 0.000084 | All vaccinated vs. Unvaccinated<br>Third dose vs. Unvaccinated<br>Two doses vs. Unvaccinated<br>Third dose vs. Two doses | $X^2$ = 104.5, df = 1, p<0.001<br>$X^2$ = 121.11, df = 1, p<0.001<br>$X^2$ = 21.615, df = 1, pp<0.001<br>$X^2$ = 25.946, df = 1, p<0.001 |
| **Feb 28-Mar 27, 2022** | 0.000199 | 0.000150 | 0.000211 | 0.000120 | All vaccinated vs. Unvaccinated<br>Third dose vs. Unvaccinated<br>Two doses vs. Unvaccinated<br>Third dose vs. Two doses | $X^2$ = 249.43, df = 1, p<0.001<br>$X^2$ = 309.05, df = 1, p<0.001<br>$X^2$ = 26.895, df = 1, p<0.001<br>$X^2$ = 117.1, df = 1, p<0.001 |

**Table S8b: Two-proportions test with continuity adjustment of SARS-CoV2 hospitalizations among NIMS vaccinated population of over 18 years of age since August 16, 2021 to March 27, 2022.** [‡]SARS-CoV2 cases tested positive with the specimen date ≥14 days post second dose **are** considered vaccinated with two doses. [ø] SARS-CoV2 cases tested positive with the specimen date ≥14 days post third dose **are** considered vaccinated with the third dose.

| Study period (rolling 4 weeks) | Proportion of population (deaths/population) | | | | Groups compared | Statistical analysis |
|---|---|---|---|---|---|---|
| | All vaccinated | 2 doses‡ | Third dose ø | Unvaccinated | | Two-Proportions test with continuity adjustment (X2=; df=; p-value) |
| **Aug 16-Sept 12, 2021** | 0.000056 | 0.000057 | | 0.000068 | All vaccinated vs. Unvaccinated<br>Two doses vs. Unvaccinated | $X^2$ = 20.926, df = 1, p = 1<br>$X^2$ = 15.286, df = 1, p = 1 |
| **Aug 23-Sept 19, 2021** | 0.000060 | 0.000062 | | 0.000069 | All vaccinated vs. Unvaccinated<br>Two doses vs. Unvaccinated | $X^2$ = 9.0561, df = 1, p=0.9987<br>$X^2$ = 5.8408, df = 1, p = 0.9922 |
| **Aug 30-Sept 26, 2021** | 0.000062 | 0.000064 | | 0.000065 | All vaccinated vs. Unvaccinated<br>Two doses vs. Unvaccinated | $X^2$ = 1.5266, df = 1, p = 0.8917<br>$X^2$ = 0.26222, df = 1, p = 0.6957 |
| **Sept 13-Oct 10, 2021** | 0.000056 | 0.000061 | | 0.000053 | All vaccinated vs. Unvaccinated<br>Two doses vs. Unvaccinated | $X^2$ = 0.84631, df = 1, p= 0.1788<br>$X^2$ = 7.6872, df = 1, p = 0.002781 |
| **Oct 11-Nov 7, 2021** | 0.000070 | 0.000094 | | 0.000057 | All vaccinated vs. Unvaccinated<br>Two doses vs. Unvaccinated | $X^2$ = 20.339, df = 1, p<0.001<br>$X^2$ = 116.92, df = 1, p<0.001 |
| **Nov 8-Dec 5, 2021** | 0.000064 | 0.000117 | | 0.000071 | All vaccinated vs. Unvaccinated<br>Two doses vs. Unvaccinated | $X^2$= 6.4766, df = 1, p= 0.9945<br>$X^2$ = 136.46, df = 1, p<0.001 |
| **Nov 22-Dec 19, 2021** | 0.000053 | 0.000143 | | 0.000080 | All vaccinated vs. Unvaccinated<br>Two doses vs. Unvaccinated | $X^2$ = 100.78, df = 1, p = 1<br>$X^2$ = 195.77, df = 1, p<0.001 |
| **Dec 6, 2021-Jan 2, 2022** | 0.000051 | 0.000189 | | 0.000084 | All vaccinated vs. Unvaccinated<br>Two doses vs. Unvaccinated | $X^2$ = 154.01, df = 1, p = 1<br>$X^2$ = 390.22, df = 1, p<0.001 |
| **Dec 20, 2021-Jan 16, 2022** | 0.000070 | 0.000163 | 0.000041 | 0.000107 | All vaccinated vs. Unvaccinated<br>Third dose vs. Unvaccinated<br>Two doses vs. Unvaccinated<br>Third dose vs. Two doses | $X^2$ = 137.28, df = 1, p<0.001<br>$X^2$ = 542.63, df = 1, p = 1<br>$X^2$ = 104.52, df = 1, p<0.001<br>$X^2$ = 1455.7, df = 1, p = 1 |
| **Dec 27, 2021-Jan 23, 2022** | 0.000088 | 0.000185 | 0.000060 | 0.000107 | All vaccinated vs. Unvaccinated<br>Third dose vs. Unvaccinated<br>Two doses vs. Unvaccinated<br>Third dose vs. Two doses | $X^2$ = 30.094, df = 1, p = 1<br>$X^2$ = 223.02, df = 1, p = 1<br>$X^2$ = 185.8, df = 1, p<0.001<br>$X^2$ = 1186, df = 1, p = 1 |

| | | | | | | |
|---|---|---|---|---|---|---|
| **Jan 10-Feb 6, 2022** | 0.000123 | 0.000201 | 0.000102 | 0.000097 | All vaccinated vs. Unvaccinated<br>Third dose vs. Unvaccinated<br>Two doses vs. Unvaccinated<br>Third dose vs. Two doses | $X^2$ = 41.017, df = 1, p<0.001<br>$X^2$ = 1.7226, df = 1, p= 0.09468<br>$X^2$ = 316.69, df = 1, p<0.001<br>$X^2$ = 506.48, df = 1, p = 1 |
| **Jan 17-Feb 13, 2022** | 0.000118 | 0.000169 | 0.000106 | 0.000078 | All vaccinated vs. Unvaccinated<br>Third dose vs. Unvaccinated<br>Two doses vs. Unvaccinated<br>Third dose vs. Two doses | $X^2$ = 107.58, df = 1, p<0.001<br>$X^2$ = 53.465, df = 1, p<0.001<br>$X^2$ = 291.24, df = 1, p<0.001<br>$X^2$ = 213.81, df = 1, p = 1 |
| **Jan 24-Feb 20, 2022** | 0.000105 | 0.000131 | 0.000100 | 0.000060 | All vaccinated vs. Unvaccinated<br>Third dose vs. Unvaccinated<br>Two doses vs. Unvaccinated<br>Third dose vs. Two doses | $X^2$ = 156.88, df = 1, p<0.001<br>$X^2$ = 122.84, df = 1, p<0.001<br>$X^2$ = 229.37, df = 1, p<0.001<br>$X^2$ = 57.692, df = 1, p = 1 |
| **Jan 31-Feb 27, 2022** | 0.000086 | 0.000093 | 0.000086 | 0.000043 | All vaccinated vs. Unvaccinated<br>Third dose vs. Unvaccinated<br>Two doses vs. Unvaccinated<br>Third dose vs. Two doses | $X^2$ = 181.54, df = 1, p<0.001<br>$X^2$ = 171.55, df = 1, p<0.001<br>$X^2$ = 159.44, df = 1, p<0.001<br>$X^2$ = 3.5199, df = 1, p = 0.9697 |
| **Feb 14-Mar 13, 2022** | 0.000053 | 0.000049 | 0.000055 | 0.000026 | All vaccinated vs. Unvaccinated<br>Third dose vs. Unvaccinated<br>Two doses vs. Unvaccinated<br>Third dose vs. Two doses | $X^2$ = 115.23, df = 1, p<0.001<br>$X^2$ = 122.59, df = 1, p<0.001<br>$X^2$ = 64.207, df = 1, p<0.001<br>V = 2.8687, df = 1, p = 0.04516 |
| **Feb 28-Mar 27, 2022** | 0.000043 | 0.000028 | 0.000049 | 0.000023 | All vaccinated vs. Unvaccinated<br>Third dose vs. Unvaccinated<br>Two doses vs. Unvaccinated<br>Third dose vs. Two doses | $X^2$ = 74.306, df = 1, p<0.001<br>$X^2$= 106.86, df = 1, p<0.001<br>$X^2$ = 3.5483, df = 1, p = 0.0298<br>$X^2$ = 58.664, df = 1, p<0.001 |

**Table S8c: Two-proportions test with continuity adjustment of SARS-CoV2 deaths among NIMS vaccinated population of over 18 years of age from August 16, 2021 to March 27, 2022.** [ǂ]SARS-CoV2 cases tested positive with the specimen date ≥14 days post second dose **are** considered vaccinated with two doses. [ø] SARS-CoV2 cases tested positive with the specimen date ≥14 days post third dose **are** considered vaccinated with the third dose.

| Study period (rolling 4 weeks*) | Proportion of population (cases/population) | | | | Compared groups | Statistical analysis Two-Proportions test with continuity adjustment (X2=; df=; p-value) |
|---|---|---|---|---|---|---|
| | All vaccinated | 2 doses‡ | Third dose ø | Unvaccinated | | |
| **until June 20, 2021** (Delta Variant) | 0.000486989 | 0.000269881 | | 0.000575473 | All vaccinated vs. Unvaccinated<br>Two doses vs. Unvaccinated | $X^2$ = 31.212, df = 1, p = 1<br>$X^2$ = 612.53, df = 1, p = 1 |
| **June 21-July 18, 2021** (Delta Variant) | 0.000441024 | 0.000413819 | | 0.000504709 | All vaccinated vs. Unvaccinated<br>Two doses vs. Unvaccinated | $X^2$ = 17.288, df = 1, p = 1<br>$X^2$ = 37.234, df = 1, p = 1 |
| **July 19-Aug 1, 2021** (Delta Variant) | 0.000413326 | 0.000404604 | | 0.000526581 | All vaccinated vs. Unvaccinated<br>Two doses vs. Unvaccinated | $X^2$ = 57.242, df = 1, p = 1<br>$X^2$ = 67.473, df = 1, p = 1 |
| **Aug 2-Aug 15, 2021** (Delta Variant) | 0.000578857 | 0.000569373 | | 0.000699815 | All vaccinated vs. Unvaccinated<br>Two doses vs. Unvaccinated | $X^2$ = 46.5, df = 1, p = 1<br>$X^2$ = 54.749, df = 1, p = 1 |
| **Aug 30-Sept 12, 2021** (Delta Variant) | 0.000938593 | 0.00092997 | | 0.000891319 | All vaccinated vs. Unvaccinated<br>Two doses vs. Unvaccinated | $X^2$ = 4.4235, df = 1, p = 0.018<br>$X^2$ = 2.9683, df = 1, p = 0.042 |
| **Aug 16-Sept 12, 2021*** | 0.00714124 | 0.00711672 | | 0.00551307 | All vaccinated vs. Unvaccinated<br>Two doses vs. Unvaccinated | $X^2$ = 708.94, df = 1, p<0.001<br>$X^2$ = 688.86, df = 1, p<0.001 |
| **Aug 23-Sept 19, 2021*** | 0.00675283 | 0.00674656 | | 0.00503149 | All vaccinated vs. Unvaccinated<br>Two doses vs. Unvaccinated | $X^2$ = 837.18, df = 1, p<0.001<br>$X^2$ = 830.53, df = 1, p<0.001 |
| **Aug 30-Sept 26, 2021*** | 0.00635938 | 0.00646325 | | 0.00449816 | All vaccinated vs. Unvaccinated<br>Two doses vs. Unvaccinated | $X^2$ = 1039.4, df = 1, p<0.001<br>$X^2$ = 1138.5, df = 1, p<0.001 |
| **Sept 13-Oct 10, 2021*** | 0.00635639 | 0.00712503 | | 0.00381063 | All vaccinated vs. Unvaccinated<br>Two doses vs. Unvaccinated | $X^2$ = 1954.1, df = p<0.001<br>v = 2953.9, df = 1, p<0.001 |
| **Oct 11-Nov 7, 2021*** | 0.01033954 | 0.01672072 | | 0.00557227 | All vaccinated vs. Unvaccinated<br>Two doses vs. Unvaccinated | $X^2$ = 4197.6, df = 1, p<0.001<br>$X^2$ = 14260, df = 1, p<0.001 |
| **Nov 8-Dec 5, 2021*** | 0.00883519 | 0.03083521 | | 0.00651932 | All vaccinated vs. Unvaccinated<br>Two doses vs. Unvaccinated | $X^2$ = 1119, df = 1, p<0.001<br>$X^2$ = 35890, df = 1, p<0.001 |
| **Dec 6, 2021-Jan 2, 2022*** | 0.02429517 | 0.23991204 | | 0.01421828 | All vaccinated vs. Unvaccinated<br>Two doses vs. Unvaccinated | $X^2$ = 7742.2, df = 1, p<0.001<br>$X^2$ = 441724, df = 1, p<0.001 |

| | | | | | | |
|---|---|---|---|---|---|---|
| **Dec 20, 2021-Jan 16, 2022*** | 0.03417810 | 0.08823135 | 0.02910401 | 0.01783886 | All vaccinated vs. Unvaccinated<br>Third dose vs. Unvaccinated<br>Two doses vs. Unvaccinated<br>Third dose vs. Two doses | $X^2$ = 14577, df = 1, p<0.001<br>$X^2$ = 7987, df = 1, p<0.001<br>$X^2$ = 91810, df = 1, p<0.001<br>$X^2$ = 168485, df = 1, p = 1 |
| **Dec 27, 2021-Jan 23, 2022*** | 0.03200767 | 0.06802698 | 0.02876190 | 0.01625914 | All vaccinated vs. Unvaccinated<br>Third dose vs. Unvaccinated<br>Two doses vs. Unvaccinated<br>Third dose vs. Two doses | $X^2$ = 14404, df = 1, p<0.001<br>$X^2$ = 9969.2, df = 1, p<0.001<br>$X^2$ = 61097, df = 1, p<0.001<br>$X^2$ = 76888, df = 1, p = 1 |
| **Jan 3-Jan 30, 2022*** | 0.02597837 | 0.04536145 | 0.02429350 | 0.01378302 | All vaccinated vs. Unvaccinated<br>Third dose vs. Unvaccinated<br>Two doses vs. Unvaccinated<br>Third dose vs. Two doses | $X^2$ = 10531, df = 1, p<0.001<br>$X^2$ = 8281.8, df = 1, p<0.001<br>$X^2$ = 31550, df = 1, p<0.001<br>$X^2$ = 26421, df = 1, p = 1 |
| **Jan 17-Feb 13, 2022*** | 0.01770613 | 0.02044630 | 0.01751527 | 0.00797034 | All vaccinated vs. Unvaccinated<br>Third dose vs. Unvaccinated<br>Two doses vs. Unvaccinated<br>Third dose vs. Two doses | $X^2$ = 9808.3, df = 1, p<0.001<br>$X^2$ = 9486.1, df = 1, p<0.001<br>$X^2$ = 9912.7, df = 1, p<0.001<br>$X^2$ = 722.09, df = 1, p = 1 |
| **Jan 31-Feb 27, 2022*** | 0.01292623 | 0.01245956 | 0.01418682 | 0.00524788 | All vaccinated vs. Unvaccinated<br>Third dose vs. Unvaccinated<br>Two doses vs. Unvaccinated<br>Third dose vs. Two doses | $X^2$ = 8334.2, df = 1, p<0.001<br>$X^2$ = 10303, df = 1, p<0.001<br>$X^2$ = 5228.9, df = 1, p<0.001<br>$X^2$ = 309.3, df = 1, p<0.001 |
| **Feb 14-Mar 13, 2022*** | 0.01538218 | 0.01072085 | 0.01585947 | 0.00466257 | All vaccinated vs. Unvaccinated<br>Third dose vs. Unvaccinated<br>Two doses vs. Unvaccinated<br>Third dose vs. Two doses | $X^2$ = 13786, df = 1, p<0.001<br>$X^2$ = 14569, df = 1, p<0.001<br>$X^2$ = 4210.6, df = 1, p<0.001<br>$X^2$ = 2455.8, df = 1, p<0.001 |
| **Feb 28-Mar 27, 2022*** | 0.02480554 | 0.01596269 | 0.02718747 | 0.00689690 | All vaccinated vs. Unvaccinated<br>Third dose vs. Unvaccinated<br>Two doses vs. Unvaccinated<br>Third dose vs. Two doses | $X^2$ = 24098, df = 1, p<0.001<br>$X^2$ = 28284, df = 1, p<0.001<br>$X^2$ = 6334.1, df = 1, p<0.001<br>$X^2$ = 6868.5, df = 1, p<0.001 |

**Table S8d: Two-proportions test with continuity adjustment of SARS-CoV2 cases among NIMS vaccinated population of over 50 years of age from August 16, 2021 to March 27, 2022 and the Delta variant cases until September 12, 2021.** ‡SARS-CoV2 cases tested positive with the specimen date ≥14 days post second dose **are** considered vaccinated with two doses. [ø] SARS-CoV2 cases tested positive with the specimen date ≥14 days post third dose **are** considered vaccinated with the third dose.

| Study period (rolling 4 weeks*) | Proportion of population (hospitalizations/population) | | | | Compared groups | Statistical analysis Two-Proportions test with continuity adjustment (X2=; df=; p-value) |
|---|---|---|---|---|---|---|
| | All vaccinated | 2 doses‡ | Third dose ø | Unvaccinated | | |
| **until June 20, 2021** (Delta Variant) | 0.00002049 | 0.00001366 | | 0.00008857 | All vaccinated vs. Unvaccinated | $X^2$= 336.46, df = 1, p = 1 |
| | | | | | Two doses vs. Unvaccinated | $X^2$= 517.31, df = 1, p = 1 |
| **June 21-July 18, 2021** (Delta Variant) | 0.00002434 | 0.00002212 | | 0.00011556 | All vaccinated vs. Unvaccinated | $X^2$ = 482.5, df = 1, p = 1 |
| | | | | | Two doses vs. Unvaccinated | $X^2$ = 533.6, df = 1, p = 1 |
| **July 19-Aug 1, 2021** (Delta Variant) | 0.00002313 | 0.00002153 | | 0.00010980 | All vaccinated vs. Unvaccinated | $X^2$ = 454.62, df = 1, p = 1 |
| | | | | | Two doses vs. Unvaccinated | $X^2$ = 490.35, df = 1, p = 1 |
| **Aug 2-Aug 15, 2021** (Delta Variant) | 0.00003779 | 0.00003545 | | 0.00015385 | All vaccinated vs. Unvaccinated | $X^2$ = 520.52, df = 1, p = 1 |
| | | | | | Two doses vs. Unvaccinated | $X^2$ = 562.56, df = 1, p = 1 |
| **Aug 16-Aug 29, 2021*** | 0.00004241 | 0.00004067 | | 0.00016193 | All vaccinated vs. Unvaccinated | $X^2$ = 498.22, df = 1, p = 1 |
| | | | | | Two doses vs. Unvaccinated | $X^2$ = 525.21, df = 1, p = 1 |
| **Aug 16-Sept 12, 2021*** | 0.00019764 | 0.00019316 | | 0.00066460 | All vaccinated vs. Unvaccinated | $X^2$ = 1686, df = 1, p = 1 |
| | | | | | Two doses vs. Unvaccinated | $X^2$ = 1738.6, df = 1, p = 1 |
| **Aug 23-Sept 19, 2021*** | 0.00020657 | 0.00020296 | | 0.00065748 | All vaccinated vs. Unvaccinated | $X^2$ = 1520.9, df = 1, p = 1 |
| | | | | | Two doses vs. Unvaccinated | $X^2$ = 1557.6, df = 1, p = 1 |
| **Aug 30-Sept 26, 2021*** | 0.00019658 | 0.00019615 | | 0.00058376 | All vaccinated vs. Unvaccinated | $X^2$ = 1194.4, df = 1, p = 1 |
| | | | | | Two doses vs. Unvaccinated | $X^2$ = 1188.7, df = 1, p = 1 |
| **Sept 13-Oct 10, 2021*** | 0.00013129 | 0.00014412 | | 0.00032709 | All vaccinated vs. Unvaccinated | $X^2$ = 471.89, df = 1, p = 1 |
| | | | | | Two doses vs. Unvaccinated | $X^2$ = 372.25, df = 1, p = 1 |
| **Oct 11-Nov 7, 2021*** | 0.00028050 | 0.00044881 | | 0.00069854 | All vaccinated vs. Unvaccinated | $X^2$ = 997.52, df = 1, p = 1 |
| | | | | | Two doses vs. Unvaccinated | $X^2$ = 221.12, df = 1, p = 1 |
| **Nov 8-Dec 5, 2021*** | 0.00018716 | 0.00063849 | | 0.00074169 | All vaccinated vs. Unvaccinated | $X^2$ = 2331.7, df = 1, p = 1 |
| | | | | | Two doses vs. Unvaccinated | $X^2$ = 23.277, df = 1, p = 1 |
| **Dec 6, 2021-Jan 2, 2022*** | 0.00020196 | 0.00191293 | | 0.00092076 | All vaccinated vs. Unvaccinated | $X^2$ = 3428.6, df = 1, p = 1 |
| | | | | | Two doses vs. Unvaccinated | $X^2$ = 678.34, df = 1, p<0.001 |
| **Dec 20, 2021-Jan 16, 2022*** | 0.00035854 | 0.00140191 | 0.00024814 | 0.00099393 | All vaccinated vs. Unvaccinated | $X^2$ = 1697, df = 1, p = 1 |

| | | | | | | |
|---|---|---|---|---|---|---|
| | | | | | Third dose vs. Unvaccinated<br>Two doses vs. Unvaccinated<br>Third dose vs. Two doses | $X^2$ = 3010.4, df = 1, p = 1<br>$X^2$ = 126.61, df = 1, p<0.001<br>$X^2$ = 6101.6, df = 1, p = 1 |
| **Dec 27, 2021-Jan 23, 2022*** | 0.00041049 | 0.00142582 | 0.00030687 | 0.00092962 | All vaccinated vs. Unvaccinated<br>Third dose vs. Unvaccinated<br>Two doses vs. Unvaccinated<br>Third dose vs. Two doses | $X^2$ = 1024.9, df = 1, p = 1<br>$X^2$ = 1823.1, df = 1, p = 1<br>$X^2$ = 188.07, df = 1, p<0.001<br>$X^2$ = 4841.4, df = 1, p = 1 |
| **Jan 17-Feb 13, 2022*** | 0.00025935 | 0.00062660 | 0.00022149 | 0.00042185 | All vaccinated vs. Unvaccinated<br>Third dose vs. Unvaccinated<br>Two doses vs. Unvaccinated<br>Third dose vs. Two doses | $X^2$ = 165.77, df = 1, p = 1<br>$X^2$ = 284.7, df = 1, p = 1<br>$X^2$ = 69.562, df = 1, p<0.001<br>$X^2$ = 948.59, df = 1, p = 1 |
| **Jan 31-Feb 27, 2022*** | 0.00022910 | 0.00044469 | 0.00020744 | 0.00029924 | All vaccinated vs. Unvaccinated<br>Third dose vs. Unvaccinated<br>Two doses vs. Unvaccinated<br>Third dose vs. Two doses | $X^2$ = 35.636, df = 1, p = 1<br>$X^2$ = 66.058, df = 1, p = 1<br>$X^2$ = 48.828, df = 1, p<0.001<br>$X^2$ = 357.61, df = 1, p = 1 |
| **Feb 14-Mar 13, 2022*** | 0.00020319 | 0.00033807 | 0.00019016 | 0.00021887 | All vaccinated vs. Unvaccinated<br>Third dose vs. Unvaccinated<br>Two doses vs. Unvaccinated<br>Third dose vs. Two doses | $X^2$ = 1.9844, df = 1, p-value = 0.92<br>$X^2$ = 7.1195, df = 1, p-value = 0.996<br>$X^2$ = 43.462, df = 1, p<0.001<br>$X^2$ = 153.28, df = 1, p = 1 |
| **Feb 28-Mar 27, 2022*** | 0.00032888 | 0.00045991 | 0.00031621 | 0.00028400 | All vaccinated vs. Unvaccinated<br>Third dose vs. Unvaccinated<br>Two doses vs. Unvaccinated<br>Third dose vs. Two doses | $X^2$ = 10.468, df = 1, p<0.001<br>$X^2$ = 5.523, df = 1, p = 0.0093<br>$X^2$ = 70.784, df = 1, p<0.001<br>$X^2$ = 87.958, df = 1, p = 1 |

**Table S8e: Two-proportions test with continuity adjustment of SARS-CoV2 hospitalizations among NIMS vaccinated population of over 50 years of age from August 16, 2021 to March 27, 2022 and the Delta variant cases until September 12, 2021.** ‡SARS-CoV2 cases tested positive with the specimen date ≥14 days post second dose **are** considered vaccinated with two doses. ø SARS-CoV2 cases tested positive with the specimen date ≥14 days post third dose **are** considered vaccinated with the third dose.

| Study period (rolling 4 weeks*) | Proportion of population (deaths/population) | | | | Compared groups | Statistical analysis Two-Proportions test with continuity adjustment (X2=; df=; p-value) |
|---|---|---|---|---|---|---|
| | All vaccinated | 2 doses‡ | Third dose ø | Unvaccinated | | |
| **until June 20, 2021** (Delta Variant) | 0.00000778 | 0.00000598 | | 0.00003225 | All vaccinated vs. Unvaccinated<br>Two doses vs. Unvaccinated | $X^2$ = 114.41, df = 1, p = 1<br>$X^2$ = 154.52, df = 1, p = 1 |
| **June 21-July 18, 2021** (Delta Variant) | 0.00000594 | 0.00000525 | | 0.00002830 | All vaccinated vs. Unvaccinated<br>Two doses vs. Unvaccinated | $X^2$ = 116.63, df = 1, p = 1<br>$X^2$ = 132.9, df = 1, p = 1 |
| **July 19-Aug 1, 2021** (Delta Variant) | 0.00000887 | 0.00000850 | | 0.00003533 | All vaccinated vs. Unvaccinated<br>Two doses vs. Unvaccinated | $X^2$ = 115.01, df = 1, p = 1<br>$X^2$ = 120.95, df = 1, p = 1 |
| **Aug 2-Aug 15, 2021** (Delta Variant) | 0.00001395 | 0.00001319 | | 0.00005450 | All vaccinated vs. Unvaccinated<br>Two doses vs. Unvaccinated | $X^2$ = 172.67, df = 1, p = 1<br>$X^2$ = 185.31, df = 1, p = 1 |
| **Aug 30-Sept 12, 2021** (Delta Variant) | 0.00002138 | 0.00002011 | | 0.00005787 | All vaccinated vs. Unvaccinated<br>Two doses vs. Unvaccinated | $X^2$ = 99.209, df = 1, p = 1<br>$X^2$ = 110.91, df = 1, p = 1 |
| **Aug 16-Sept 12, 2021*** | 0.000105248 | 0.00010247 | | 0.00029315 | All vaccinated vs. Unvaccinated<br>Two doses vs. Unvaccinated | $X^2$ = 534.8, df = 1, p = 1<br>$X^2$ = 559.64, df = 1, p = 1 |
| **Aug 23-Sept 19, 2021*** | 0.00011445 | 0.00011185 | | 0.00030198 | All vaccinated vs. Unvaccinated<br>Two doses vs. Unvaccinated | $X^2$ = 494.27, df = 1, p = 1<br>$X^2$ = 515, df = 1, p-value = 1 |
| **Aug 30-Sept 26, 2021*** | 0.00011705 | 0.00011634 | | 0.00028868 | All vaccinated vs. Unvaccinated<br>Two doses vs. Unvaccinated | $X^2$ = 409.22, df = 1, p = 1<br>$X^2$ = 411.49, df = 1, p = 1 |
| **Sept 13-Oct 10, 2021*** | 0.00010668 | 0.00011701 | | 0.00024135 | All vaccinated vs. Unvaccinated<br>Two doses vs. Unvaccinated | $X^2$ = 279.44, df = 1, p = 1<br>$X^2$ = 215.47, df = 1, p = 1 |
| **Oct 11-Nov 7, 2021*** | 0.00013501 | 0.00021503 | | 0.00025680 | All vaccinated vs. Unvaccinated<br>Two doses vs. Unvaccinated | $X^2$ = 183.77, df = 1, p = 1<br>$X^2$ = 13.381, df = 1, p = 0.9999 |
| **Nov 8-Dec 5, 2021*** | 0.00012198 | 0.00041925 | | 0.00031641 | All vaccinated vs. Unvaccinated<br>Two doses vs. Unvaccinated | $X^2$ = 484.81, df = 1, p = 1<br>$X^2$ = 39.061, df = 1, p<0.001 |
| **Dec 6, 2021-Jan 2, 2022*** | 0.00009639 | 0.00091918 | | 0.00036390 | All vaccinated vs. Unvaccinated<br>Two doses vs. Unvaccinated | $X^2$ = 1046.9, df = 1, p = 1<br>$X^2$ = 467.11, df = 1, p<0.001 |
| **Dec 20, 2021-Jan 16, 2022*** | 0.00013310 | 0.00077283 | 0.00006678 | 0.00047476 | All vaccinated vs. Unvaccinated | $X^2$ = 1247.8, df = 1, p = 1 |

| | | | | | | |
|---|---|---|---|---|---|---|
| | | | | | Third dose vs. Unvaccinated<br>Two doses vs. Unvaccinated<br>Third dose vs. Two doses | $X^2$ = 2731.4, df = 1, p = 1<br>$X^2$ = 129.96, df = 1, p<0.001<br>$X^2$ = 6227.2, df = 1, p = 1 |
| **Dec 27, 2021-Jan 23, 2022*** | 0.00016911 | 0.00088350 | 0.00009779 | 0.00047887 | All vaccinated vs. Unvaccinated<br>Third dose vs. Unvaccinated<br>Two doses vs. Unvaccinated<br>Third dose vs. Two doses | $X^2$ = 848.38, df = 1, p = 1<br>$X^2$ = 1870.2, df = 1, p = 1<br>$X^2$ = 217.07, df = 1, p<0.001<br>$X^2$ = 5852.6, df = 1, p = 1 |
| **Jan 3-Jan 30, 2022*** | 0.00021155 | 0.00099077 | 0.00013624 | 0.00048501 | All vaccinated vs. Unvaccinated<br>Third dose vs. Unvaccinated<br>Two doses vs. Unvaccinated<br>Third dose vs. Two doses | $X^2$ = 548.74, df = 1, p = 1<br>$X^2$ = 1234.4, df = 1, p = 1<br>$X^2$ = 310.4, df = 1, p<0.001<br>$X^2$ = 5361.9, df = 1, p = 1 |
| **Jan 17-Feb 13, 2022*** | 0.00022885 | 0.00081722 | 0.00017383 | 0.00035145 | All vaccinated vs. Unvaccinated<br>Third dose vs. Unvaccinated<br>Two doses vs. Unvaccinated<br>Third dose vs. Two doses | $X^2$ = 107.58, df = 1, p = 1<br>$X^2$ = 282.23, df = 1, p = 1<br>$X^2$ = 329.59, df = 1, p<0.001<br>$X^2$ = 2704.6, df = 1, p = 1 |
| **Jan 31-Feb 27, 2022*** | 0.00016817 | 0.00044276 | 0.00014243 | 0.00019183 | All vaccinated vs. Unvaccinated<br>Third dose vs. Unvaccinated<br>Two doses vs. Unvaccinated<br>Third dose vs. Two doses | $X^2$ = 5.5114, df = 1, p-value = 0.9906<br>$X^2$ = 27.948, df = 1, p = 1<br>$X^2$ = 174.2, df = 1, p<0.001<br>$X^2$ = 782.09, df = 1, p = 1 |
| **Feb 14-Mar 13, 2022*** | 0.00010368 | 0.00023821 | 0.00009147 | 0.00011559 | All vaccinated vs. Unvaccinated<br>Third dose vs. Unvaccinated<br>Two doses vs. Unvaccinated<br>Third dose vs. Two doses | $X^2$ = 2.2098, df = 1, p-value = 0.9314<br>$X^2$ = 10.304, df = 1, p-value = 0.9993<br>$X^2$ = 73.402, df = 1, p<0.001<br>$X^2$ = 296.23, df = 1, p = 1 |
| **Feb 28-Mar 27, 2022*** | 0.00008413 | 0.00013235 | 0.00008154 | 0.00010663 | All vaccinated vs. Unvaccinated<br>Third dose vs. Unvaccinated<br>Two doses vs. Unvaccinated<br>Third dose vs. Two doses | $X^2$ = 9.8147, df = 1, p-value = 0.9991<br>$X^2$ = 12.465, df = 1, p-value = 0.9998<br>$X^2$ = 4.4595, df = 1, p-value = 0.0173<br>$X^2$ = 41.747, df = 1, p = 1 |

**Table S8f: Two-proportions test with continuity adjustment of SARS-CoV2 deaths among NIMS vaccinated population of over 50 years of age from August 16, 2021 to March 27, 2022 and the Delta variant cases until September 12, 2021.** ‡SARS-CoV2 cases tested positive with the specimen date ≥14 days post second dose **are** considered vaccinated with two doses. ø SARS-CoV2 cases tested positive with the specimen date ≥14 days post third dose **are** considered vaccinated with the third dose.

8. NHS England and NHS Improvement. COVID-19 Daily Deaths. https://www.england.nhs.uk/statistics/statistical-work-areas/covid-19-daily-deaths/
